# Genome-Wide Meta-Analysis Identifies 47 Novel Loci and Links Essential Tremor to Ventral Diencephalon and Cerebellum Morphometry

**DOI:** 10.1101/2025.11.10.25339873

**Authors:** Natalia S. Ogonowski, Fangyuan Cao, Victor Flores-Ocampo, Sofia Salazar-Magaña, Mathias Seviiri, Liyang Song, Sam Nayler, Jason Kugelman, Gabriel Cuellar-Partida, Stuart MacGregor, Hae Kyung Im, Ian H. Harding, Puya Gharahkhani, Nicholas G. Martin, Kishore R. Kumar, Jian Yang, Santiago Diaz-Torres, Miguel E. Rentería

## Abstract

Essential tremor (ET) is the most common movement disorder, yet its genetic basis remains poorly understood. We performed a genome-wide association meta-analysis including 20,268 ET cases and 723,761 neurologically healthy controls of European ancestry from the Million Veteran Program, 23andMe Research Institute and All of Us. We identified 50 independent genome-wide significant loci, including 47 novel loci. We estimated the SNP-based heritability to be 24%. Genetic correlation analyses revealed considerable overlap between ET and Parkinson’s disease, myoclonus, and systemic traits such as cardiovascular and metabolic conditions. Inverse correlations were observed with cerebellar and diencephalic volumes. Integrative analyses prioritised candidate genes through transcriptome-wide association studies across 13 brain tissues, and spatial transcriptomics highlighted enrichment of ET heritability in hippocampal and cortical excitatory neurons as well as astrocytes and microglia. Polygenic risk scores significantly predicted ET risk and age at diagnosis across European and non-European cohorts, with the strongest transferability to admixed American ancestry. These findings substantially expand the catalogue of ET risk loci, implicate excitatory neurons and cerebellar–hippocampal circuit mechanisms, and provide a foundation for biomarker discovery and therapeutic development.

## INTRODUCTION

Essential tremor (ET) is one of the most common neurological movement disorders, with a global prevalence of ∼1–5%, predominantly among individuals older than 65 years^1–3^. ET is clinically characterised by rhythmic and involuntary oscillations, affecting primarily the hands and arms; however, tremors may also extend to the head, vocal cords, or lower limbs^2^.

The distribution and severity of tremors can vary by sex, with head and vocal tremors being more prevalent among females. In contrast, postural hand tremor tends to be more frequent and severe among males^3–5^. Despite extensive research, the precise neurobiological mechanisms underlying ET remain unclear. The heterogeneous clinical presentation of ET further complicates the identification of clear etiological pathways and often leads to diagnostic overlap with other disorders, particularly Parkinson’s disease (PD)^6,7^. Consequently, therapeutic options remain limited, with current treatments primarily targeting symptomatic relief rather than disease intervention.

Several hypotheses have been proposed to explain the neurobiological basis of ET. Most implicated disruptions in cerebellar-thalamo-cortical circuitry, abnormal oscillatory neuronal activity, and functional dysregulation within subcortical regions, including the basal ganglia and ventral diencephalon^6–8^. Imaging studies, including structural and functional magnetic resonance imaging (MRI), have highlighted abnormalities within cerebellar regions in patients with ET, suggesting cerebellar dysfunction as a core feature of its pathophysiology^7,9–11^. In addition, abnormalities within subcortical structures, particularly the subthalamic nucleus and thalamic regions, have been implicated in the generation or propagation of tremor symptoms^12,13^. However, the biological mechanisms underlying the relationships between ET severity and morphometric or structural changes within these brain regions remain poorly understood.

Genetic epidemiological evidence supports ET as a heritable condition, with familial aggregation observed in approximately 50% of cases^14,15^. Heritability estimates based on twin and family studies range between 45% and 90%, emphasising a substantial genetic contribution^16,17^. Neuroimaging genetics studies have begun identifying genetic variants associated with cerebellar and subcortical structural variations that may predispose individuals to ET^18^. However, due to the limited availability of large-scale datasets and detailed clinical characterisation, these associations have often been preliminary, calling for further exploration.

Here, we present a GWAS meta-analysis including 20,268 ET cases and 723,761 controls. Our study identifies 50 independent genomic loci, 47 of which have not been reported previously. These loci implicate tissue-relevant genes and uncover significant genetic overlap between ET and structural MRI-derived volumes, particularly the ventral diencephalon and cerebellar brain regions. Altogether, the present study advances our understanding of the genetic basis of ET and provides valuable insights into the distinct neurobiological mechanisms that differentiate it from other tremor-related conditions.

## RESULTS

### ET GWAS meta-analysis

In the largest meta-analysis of ET to date, we analysed GWAS sumstats from three independent cohorts comprising 20,268 individuals with ET and 723,761 neurologically healthy controls of European ancestry. We tested ∼22 million single-nucleotide polymorphisms (SNPs) with MAF ≥ 0.01 for association with ET under an additive model. Across the meta-analysis, we identified 1,398 genome-wide significant associations (p < 5 × 10□□), of which 50 were classified as independent loci after clumping (r² < 0.1 within a 1 Mb window). Of the identified loci, 41 represent novel associations not previously reported in ET, six have been implicated in prior GWAS of unrelated traits, and three have been previously associated with ET (**Figure 1 and Supplementary Figures 1 - 3)**. After conditioning on 12 previously reported ET-associated variants, 8 out of the 12 unreported loci retained a statistically significant association with ET. Of importance, four loci (rs17432566, rs13128363, rs17650401 and rs7042473) detected as genome-wide significant in the GWAS meta-analysis were not present in the COJO output. This is likely due to a lack of representation of the variants in the LD reference panel (**Table 1, Supplementary Table 1)**

**Figure 1.**
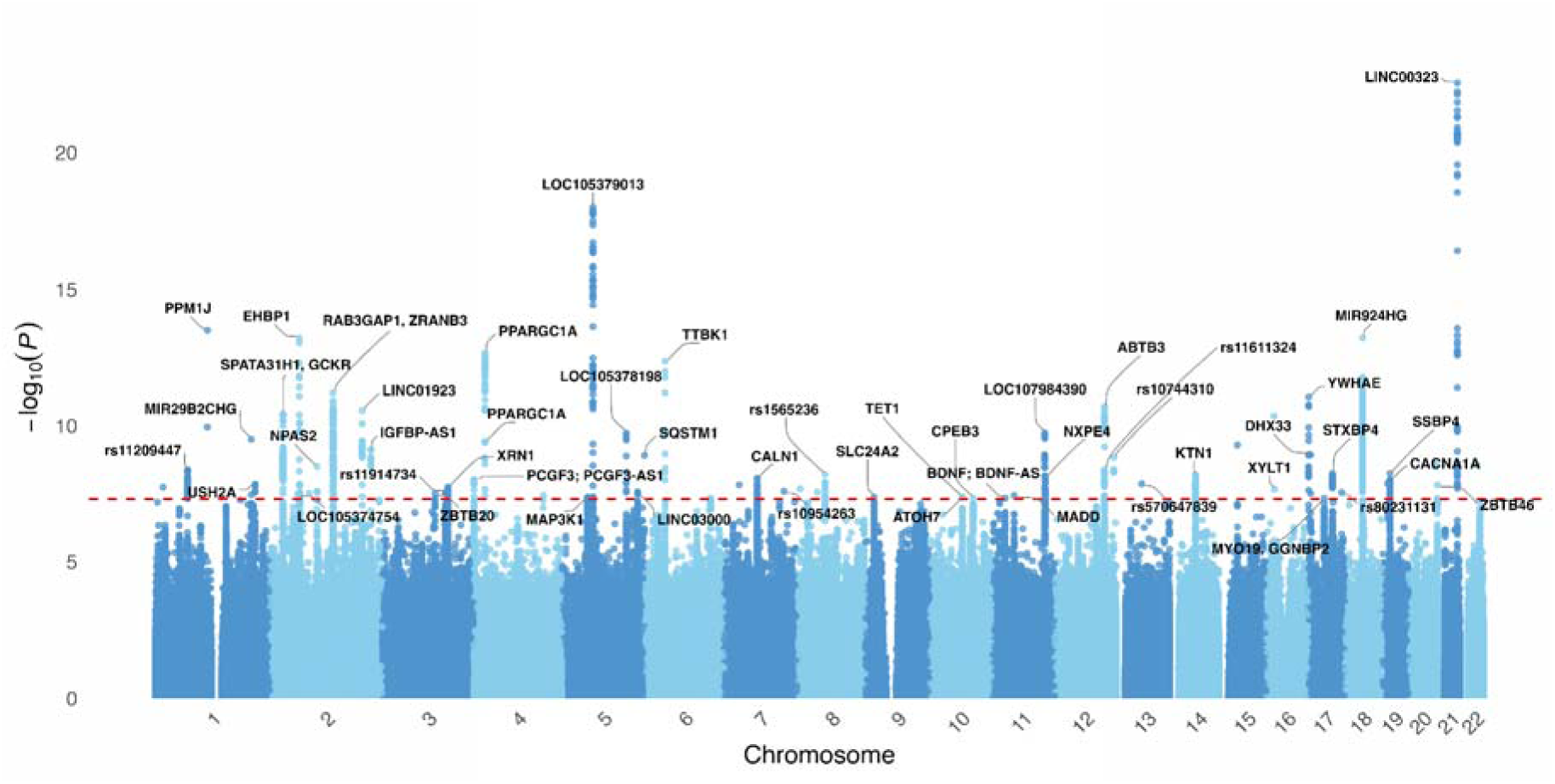
Manhattan plot of the ET gene-level association study. Each point represents a gene, positioned along the x-axis according to its genomic location across the 22 autosome chromosomes. The y-axis shows the –log□□-transformed p-values of association. The red dashed line indicates the genome-wide significance threshold p < 5e-8. Genes surpassing the genome-wide threshold are labelled, with top loci of interest annotated by gene name.

**Table 1.**
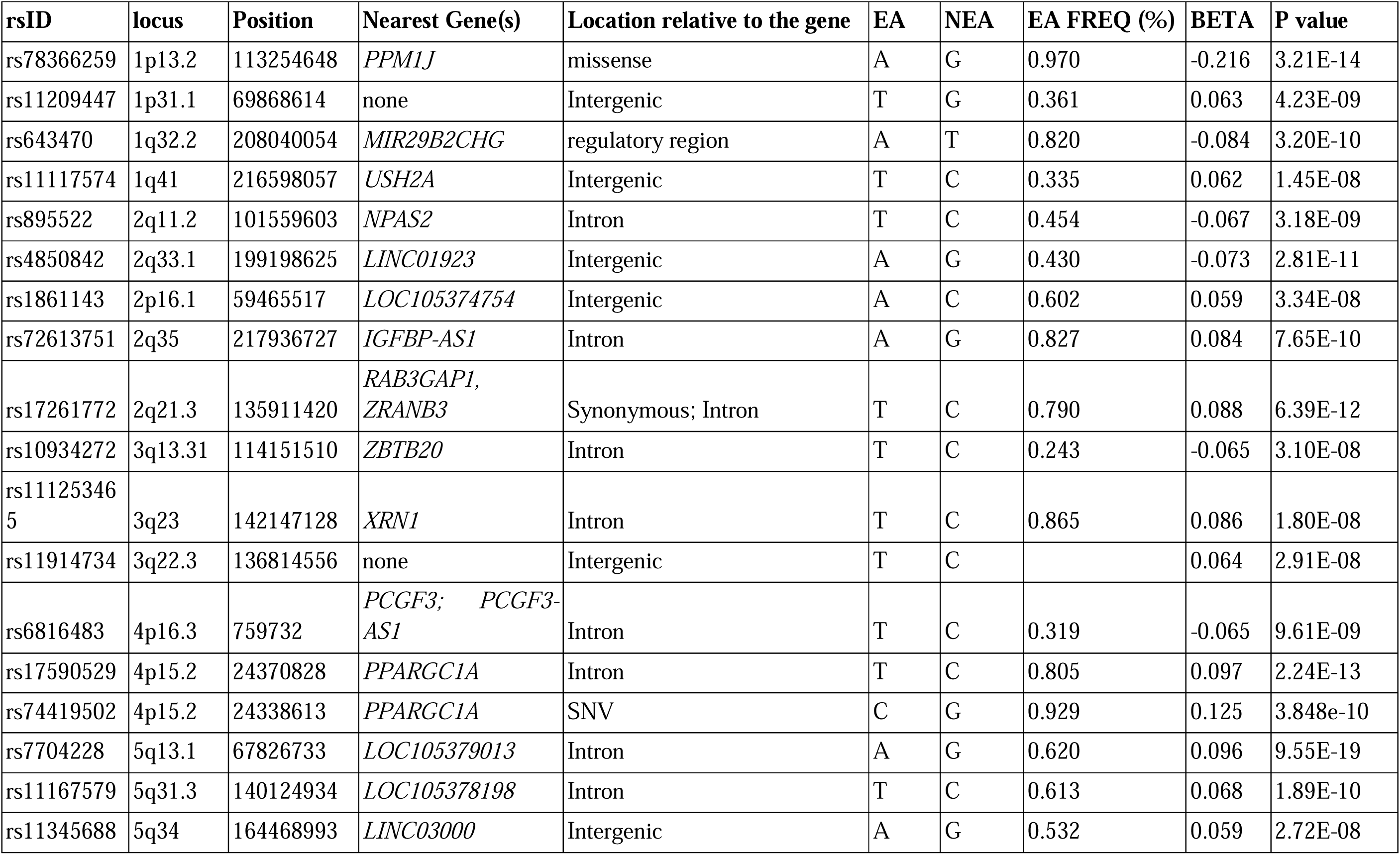
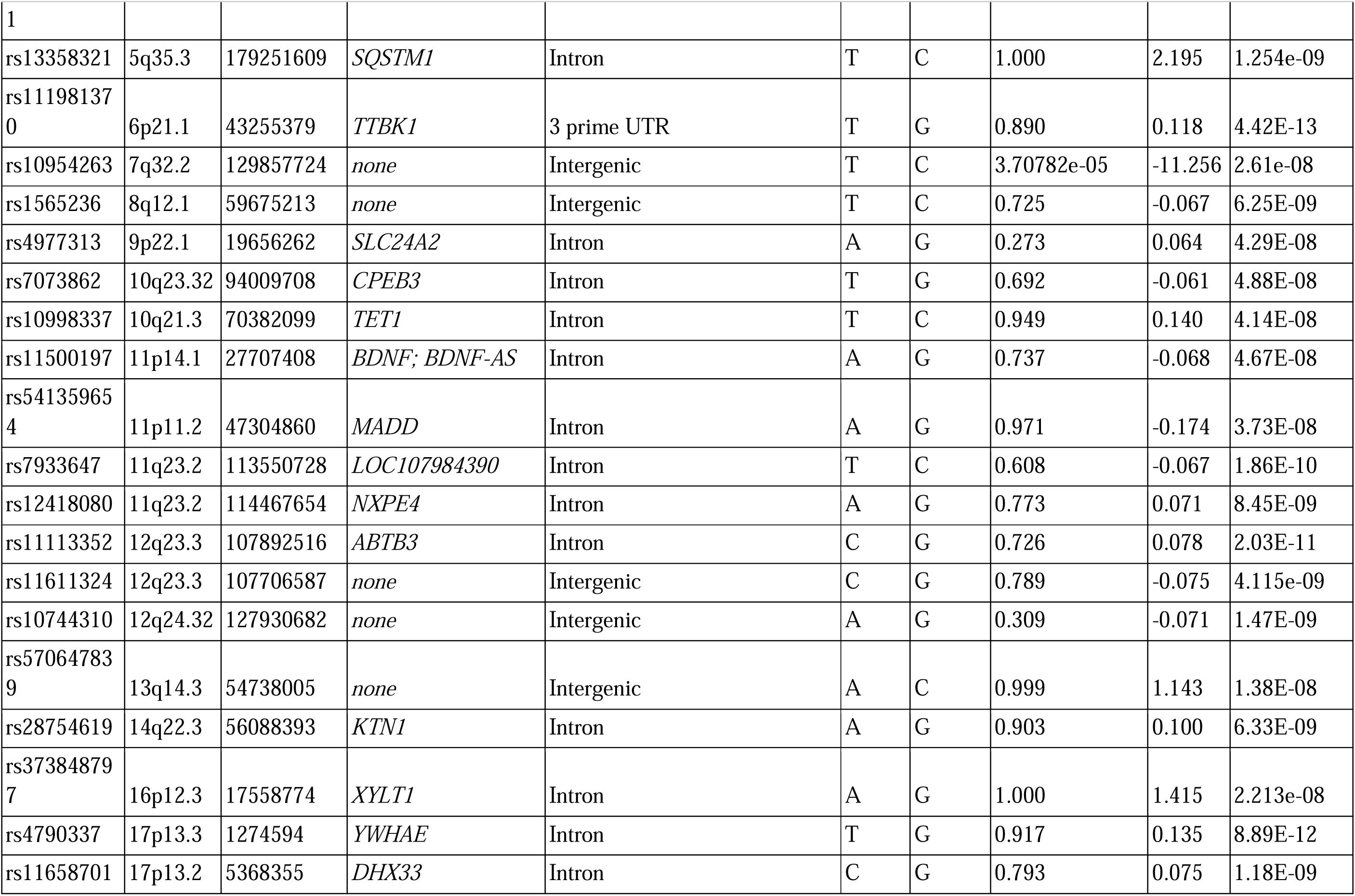
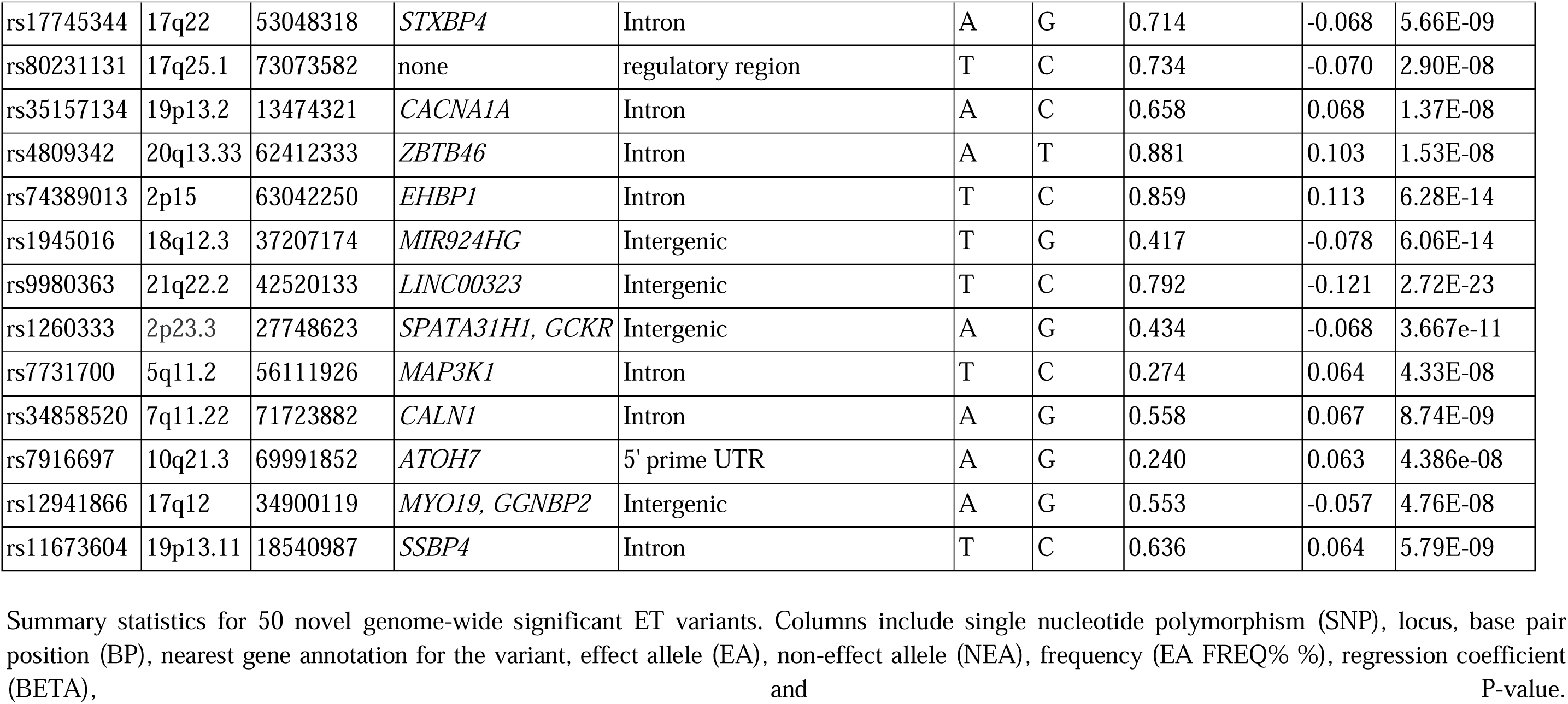
50 loci associated with essential tremor.

### SNP-based heritability

SNP-based heritability estimates indicated that common genetic variants explain a modest but significant proportion of the phenotypic variance in ET, with an observed-scale heritability of approximately 24% (h² = 0.24, SE = 0.02), corresponding to a liability-scale estimate of 18.5% assuming a population prevalence of 5%. The LD score regression intercept was 1.09 (SE = 0.01), indicating some inflation in the test statistics that could reflect contributions from polygenicity and potential confounding. However, the attenuation ratio of 0.19 suggests that the observed inflation is consistent mainly with polygenicity rather than population stratification. The genomic inflation factor (λGC) was determined to be 1.37 (mean χ² = 1.44). The attenuation ratio, which assesses the degree of inflation due to polygenicity rather than confounding, was 0.19, indicating that most inflation was attributable to polygenicity rather than population stratification. Quantile–quantile (QQ) and Manhattan plots for the meta-analysis are shown in **Supplementary Figures 4 to 6.**

### Genetic correlations

To assess the consistency of genetic effects across cohorts, we estimated pairwise genetic correlations using LDSC among the 23andMe, MVP, and AoU EUR cohorts (**Supplementary Table 2**). We observed a significant and positive genetic correlation between the MVP and 23andMe EUR cohorts (r*_G_* = 1.00, SE = 0.08; p = 5.88 × 10□^37^), indicating near-complete concordance in genetic architecture. The AoU cohort also showed strong genetic correlations with both 23andMe (r*_G_* = 0.81, SE = 0.16; p = 2.18 × 10□□) and MVP (r*_G_* = 1.01, SE = 0.11; p = 5.81 × 10□²²).

We assessed genetic correlations (r*_G_*) between ET and 2,871 complex traits using GWAS summary statistics sourced from CTG-VL, based on a common set of HapMap3 variants. Statistical significance was determined using a Benjamini–Hochberg false discovery rate (FDR) threshold of 5% to account for multiple testing. After correction, we identified 128 traits with significant genetic correlations with ET, including neurological, cardiovascular, musculoskeletal, pain-related, respiratory, gastrointestinal, pharmacological, and other complex traits, as well as brain volumes (**Supplementary Table 3, Figures 7-15**).

We observed strong positive genetic correlations between ET and several neurological traits, including PD (r*_G_* = 0.49; 95% CI: [0.41, 0.57]; p = 1.5 × 10^-7^), myoclonus (r*_G_* = 0.32; 95% CI: [0.22, 0.42]; p = 0.02), voice disturbance (r*_G_* = 0.30; 95% CI: [0.23, 0.37]; p = 0.002) and 17 other neurological conditions (**Supplementary Figure 7**).

We observed additional associations with 20 cardiovascular traits, such as stroke (r*_G_* = 0.28; 95% CI: [0.18, 0.38]; p = 0.04) and orthostatic hypotension (r*_G_* = 0.26; 95% CI: [0.20, 0.32]; p = 4 □×□10□^4^) (**Supplementary Figure 8**). Notably, ET showed significant positive correlations with multisite chronic pain (r_G_□=□0.14; 95% CI: [0.12, 0.16]; p□=□4.9□×□10□□), 13 musculoskeletal disorders (r_G_□=□0.11–0.32), 13 respiratory traits (r_G_□=□0.10–0.25), 10 metabolic traits (r_G□_=□0.10–0.22), nine pharmacological treatments and 24 other systemic conditions (r_G_□=□0.11–0.31), underscoring the broad pleiotropic architecture of ET (**Supplementary Figures 9 and 14**).

We also assessed r_G_ between ET and multiple neuroimaging measures derived from T1-weighted MRI, including subcortical volumes, cerebellar lobules, and cortical measures of surface area and thickness, using publicly available GWAS summary statistics pre-adjusted for intracranial volume by the original studies. We identified significant inverse genetic correlations with the ventral diencephalon (rG = −0.15; 95% CI: [-0.18,-0.11]; p = 0.003), a region involved in relaying motor and sensory signals, as well as integrating autonomic and limbic functions. Additionally, three cerebellar areas also showed significant inverse genetic correlations with ET: the inferior posterior lobe, including the Crus II–IX hemispheric lobules (r_G_ = −0.13; 95% CI: [−0.16, −0.09]; p = 0.009), which supports motor coordination and higher cognitive processing; the flocculonodular lobe, including the X vermal lobules (r_G_ = −0.12; 95% CI: [−0.16, −0.08]; p = 0.03), which may contribute to balance and oculomotor control; and total cerebellar volume, excluding the Crus I vermis (r_G_ = −0.12; 95% CI: [−0.15, −0.08]; p = 0.03), reflecting overall cerebellar integrity. We detected no significant genetic correlations between ET and cortical surface area and thickness measures after Benjamini-Hochberg FDR correction (**Supplementary Figure 15**).

### Transcriptome-Wide Associations

A TWAS was conducted using S-PrediXcan, integrating meta-analysed ET GWAS summary statistics with tissue-specific gene expression prediction models from GTEx v8 across 13 brain tissues, including cortical and subcortical regions, cerebellum, spinal cord, and brainstem. The TWAS identified multiple genes whose genetically predicted expression levels were significantly associated with ET after Bonferroni correction for multiple comparisons using the Bonferroni method. We then conducted a multi-tissue TWAS analysis with S-MultiXcan to integrate single-tissue associations through a multivariate approach. The strongest multi-tissue associations with ET were *BACE2* (p = 1.9 ×□10^-18^), followed by *RP11-344P13.6* (p = 2.8 ×□10^-12^) and *ZKSCAN8* (p = 5.9 ×□10^-9^) (**Figure 2**).

**Figure 2.**
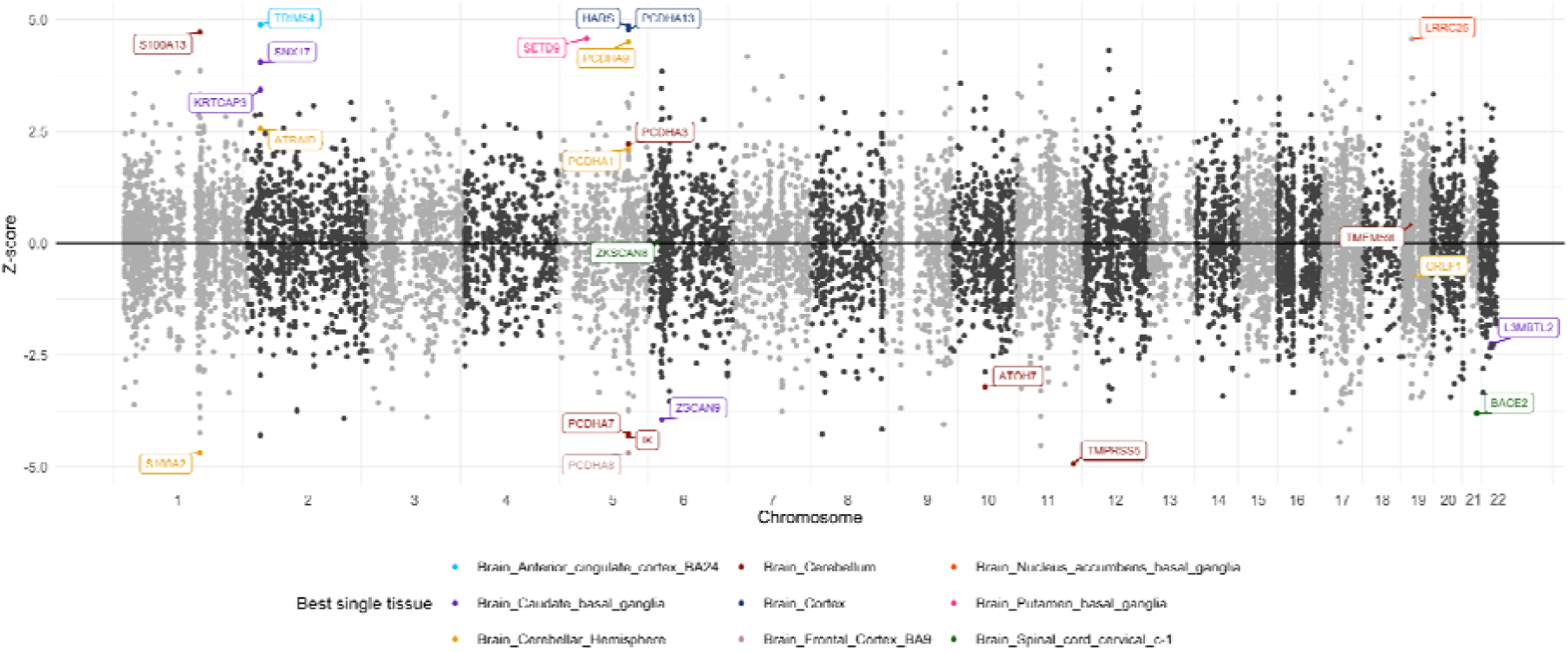
Manhattan Plot of Transcriptome-Wide Associations for Essential Tremor across brain and spinal cord tissues. Manhattan plot showing the SMultiXcan-derived z-scores for each gene, computed using shared genetic effects on gene expression across 13 GTEx v8 tissues. Each dot represents a gene whose predicted genetic expression, given by the effects of eQTLs across multiple tissues, was tested for association with ET. Gene names are annotated for those whose p-value reached a significance of alpha = 0.05 after Bonferroni correction. The x-axis shows the chromosomal positions of each gene’s canonical transcription start, while the y-axis represents the significance of the association. Colours indicate the tissue with the strongest association in the single-tissue TWAS analysis, as defined by the t_i_best field.

The top associations per tissue included *BACE2* (spinal cord cervical C-1, p = 1.4 ×□10^-20^ and substantia nigra, p = 2.6 ×□10^-17^), *PCDHA7* (cerebellum, p = 1.4 ×□10^-10^; anterior cingulate cortex, p = 3.5 ×□10^-7^), *HARS* (cortex, p = 3.4 ×□10^-10^; hypothalamus, p = 4.2 ×□10^-10^), *ATRAID* (frontal cortex, p = 9.9 ×□10^-9^; cerebellar hemisphere, p = 1.8 ×□10^-9^), *SNX17* (caudate basal ganglia, p = 1.4 ×□10^-8^; putamen basal ganglia, p = 3.4 ×□10^-8^), *RP11-344P13.6* (nucleus accumbens basal ganglia, p = 1.1 ×□10^-9^), *TMPRSS5* (hippocampus, p = 2.2 ×□10^-7^) and *S100A13* (amygdala, p = 5.9 ×□10^-7^) (**Figure 3**).

**Figure 3.**
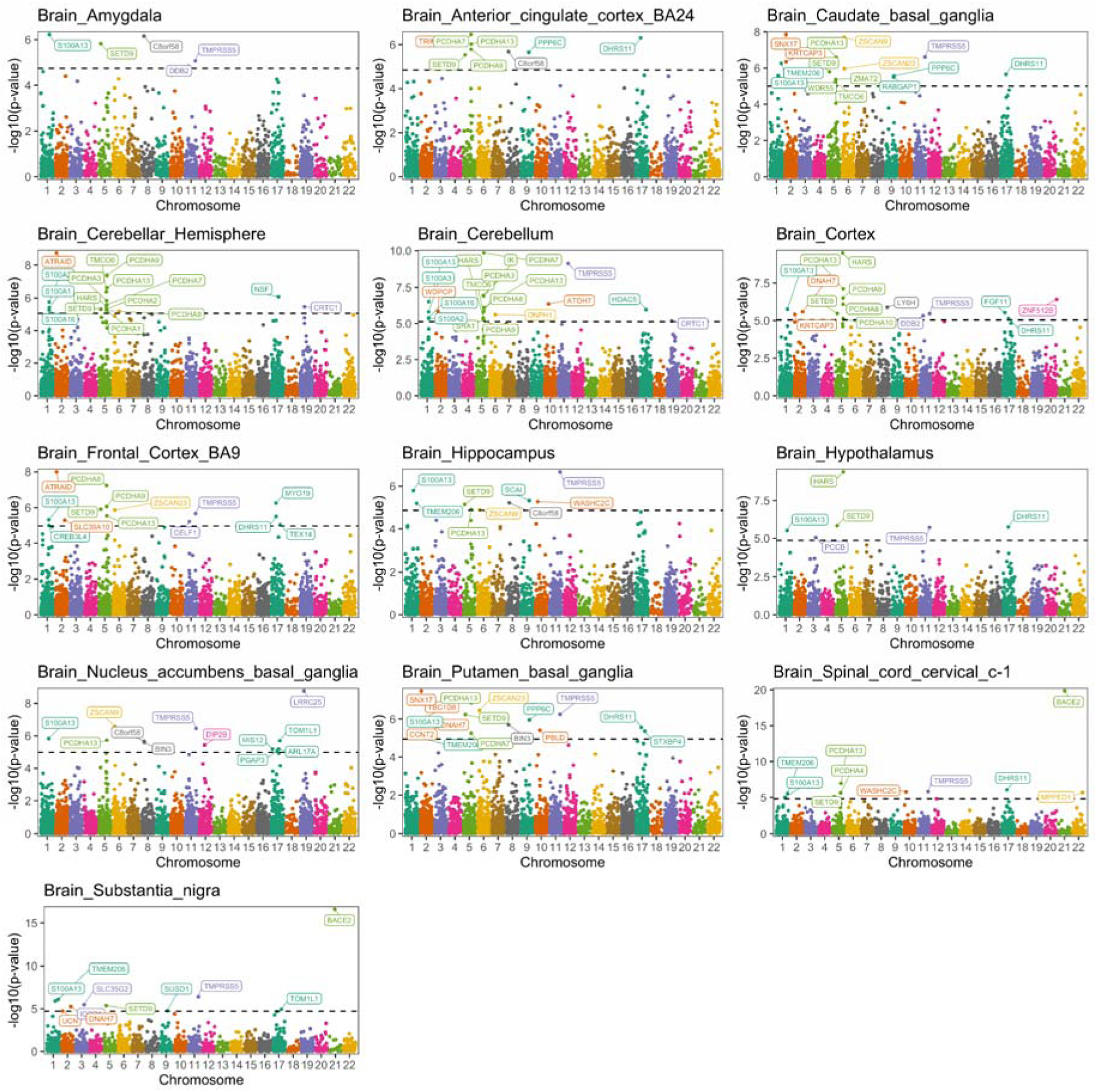
Tissue-specific Transcriptome-wide associations for ET results across multiple brain regions. Manhattan plots display the –log□□(P) values of gene-trait associations identified by S-PrediXcan across 13 brain tissues, including cortical and subcortical areas, cerebellum, spinal cord, and brainstem. Each point represents a gene whose predicted expression in a given tissue was tested for association with ET using meta-analysed GWAS summary statistics. Chromosomal positions of their canonical transcription start sites are plotted along the x-axis, and significance is plotted on the y-axis. Genes whose p-value reached a significance of alpha = 0.05 after Bonferroni correction are annotated.

### Fine-mapping with FOCUS

Because TWAS associations may arise from noncausal genes in LD with causal genes, we performed fine-mapping using FOCUS to prioritise those with a high probability of causality across all autosomes and brain tissues from GTEx v8. **Figure 4A** represents the maximum posterior inclusion probability (PIP) for a gene at a given locus, aggregated across tissues. Several loci surpassed the high-confidence threshold (PIP ≥ 0.8), including those on chromosomes 1, 2, 3, 5, 7, 10, and 16, with some signals reaching PIP = 1.0.

**Figure 4.**
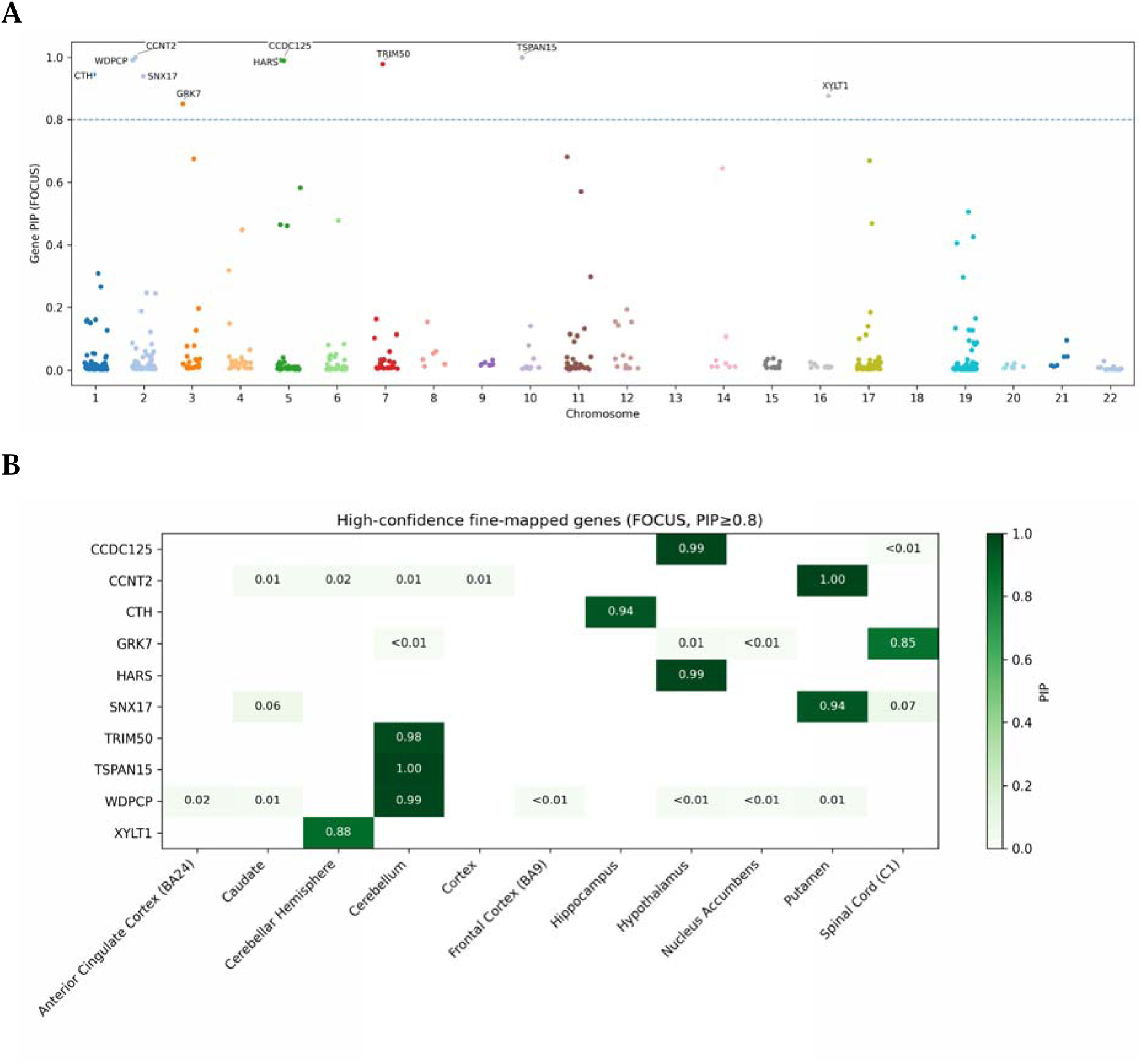
Genome-wide fine-mapping of TWAS loci using FOCUS. **A.** Manhattan-style plot showing the maximum posterior inclusion probability (PIP) for each gene locus across chromosomes 1-22, aggregated across 13 brain tissues from GTEx. Each point represents a unique gene–locus combination. The high-confidence threshold (PIP ≥ 0.8) is plotted on the y-axis. Genes above this threshold are considered strong candidates for causal involvement in ET risk. **B.** Brain-region specificity of high-confidence fine-mapped genes with PIP ≥ 0.8. Colours indicate the maximum PIP for each gene–tissue pair, with darker green representing higher probabilities. Only the top 10 genes by maximum PIP are displayed. Gene–tissue pairs without a signal are shown in pale green.

To explore tissue specificity of these high-confidence signals, we examined the distribution of genes with PIP ≥ 0.8 across brain regions (**Figure 4B**). This analysis identified ten genes — *CCDC125, CCNT2, CTH, GRK7, HARS, SNX17, TRIM50, TSPAN15, WDPCP,* and *XYLT1* —that were fine-mapped to high confidence in at least one tissue. Notably, *TRIM50, TSPAN15, XYLT1* and *WDPCP* were prioritised in the cerebellum, *HARS* and *CCDC125* in the hypothalamus, *CTH* in the hippocampus, *SNX17* and *CCNT2* in the putamen basal ganglia, and *GRK7* in the spinal cord cervical C-1.

### Colocalisation with COLOC

Additionally, we conducted a colocalisation analysis to evaluate whether the same genetic variants located near the TWAS-identified genes were responsible for both the ET risk and gene expression changes in brain tissues. The variants situated in 1MB of the significantly associated genes identified in the S-MultiXcan analysis were colocalised to expression Quantitative Trait Loci (eQTLs) derived from the GTEx v8 brain tissues. This analysis identified four candidate causal genes: *S100A13, BACE2, PCDH9*, and *HARS*. The *S100A13* gene displayed strong colocalisation across nearly all regions, except the cerebellar hemisphere, with PP.H4 values (posterior probability that there is colocalisation) close to 1. Meanwhile, *BACE2* colocalised primarily with the spinal cord cervical C-1 region (PP.H4 ∼0.7–0.8). *PCDH9* exhibited strong colocalisation in the cerebellum (PP.H4 ∼1), whereas *HARS* colocalised with the cortex and cerebellum, both with PP.H4 values of approximately 0.7-0.8 (**Figure 5**).

**Figure 5.**
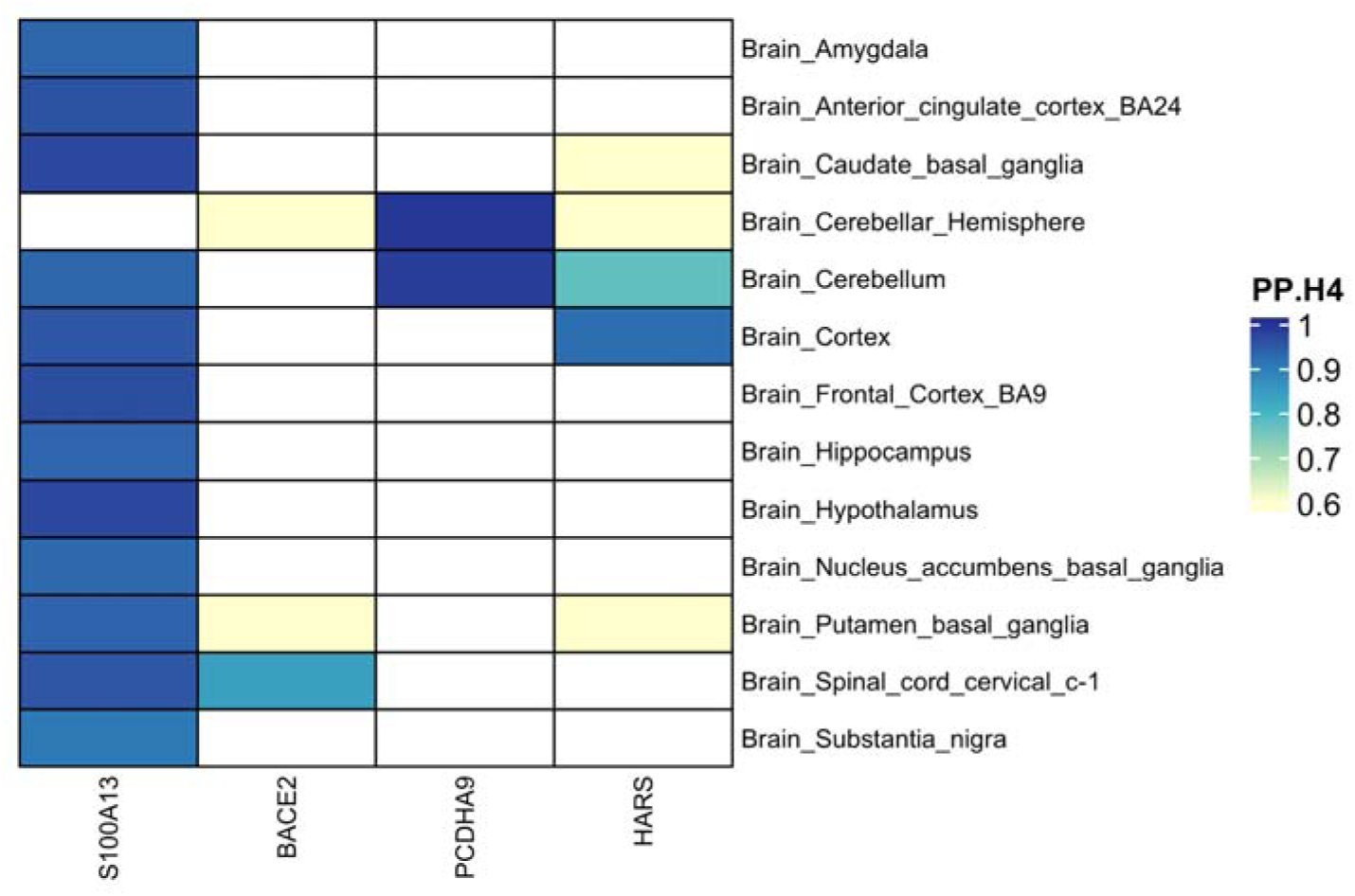
Genes with ET risk signals colocalising with brain-specific eQTLs. Genes (columns) identified from the multi-tissue TWAS using SMultiXcan that have evidence of colocalisation with eQTL signals from at least one brain tissue from GTExV8 (rows). Colocalisation is assumed if COLOC posterior probability PP.H4 > 0.8 (shown by the colour).

### Integration of GWAS with Spatial Transcriptomics using gsMap

To characterise the spatial and cellular architecture of ET, we integrated GWAS meta-analysis results with adult mouse brain spatial transcriptomics using the gsMap framework. This approach mapped ET-associated loci across transcriptionally defined single-cell populations and anatomically resolved brain regions. The strongest enrichment signals are localised to hippocampal areas, including CA1 and subiculum, as well as cortical regions, highlighting the relevance of limbic-cortical circuits in ET pathogenesis. Among cell types, excitatory neurons, particularly those in the EX L2/3, EX L5/6, EX Mb and EX CA populations, displayed the most substantial enrichment. We also observed moderate enrichment within non-neuronal populations, including astrocytes (Astr4, Astr2), microglia, and oligodendrocytes, suggesting involvement of glial cells in ET-related circuit regulation. In contrast, inhibitory neuronal subtypes, such as IN Pvalb+, IN Vip+ and IN Sst+ demonstrated lower or spatially restricted enrichment patterns (**Figure 6**).

**Figure 6.**
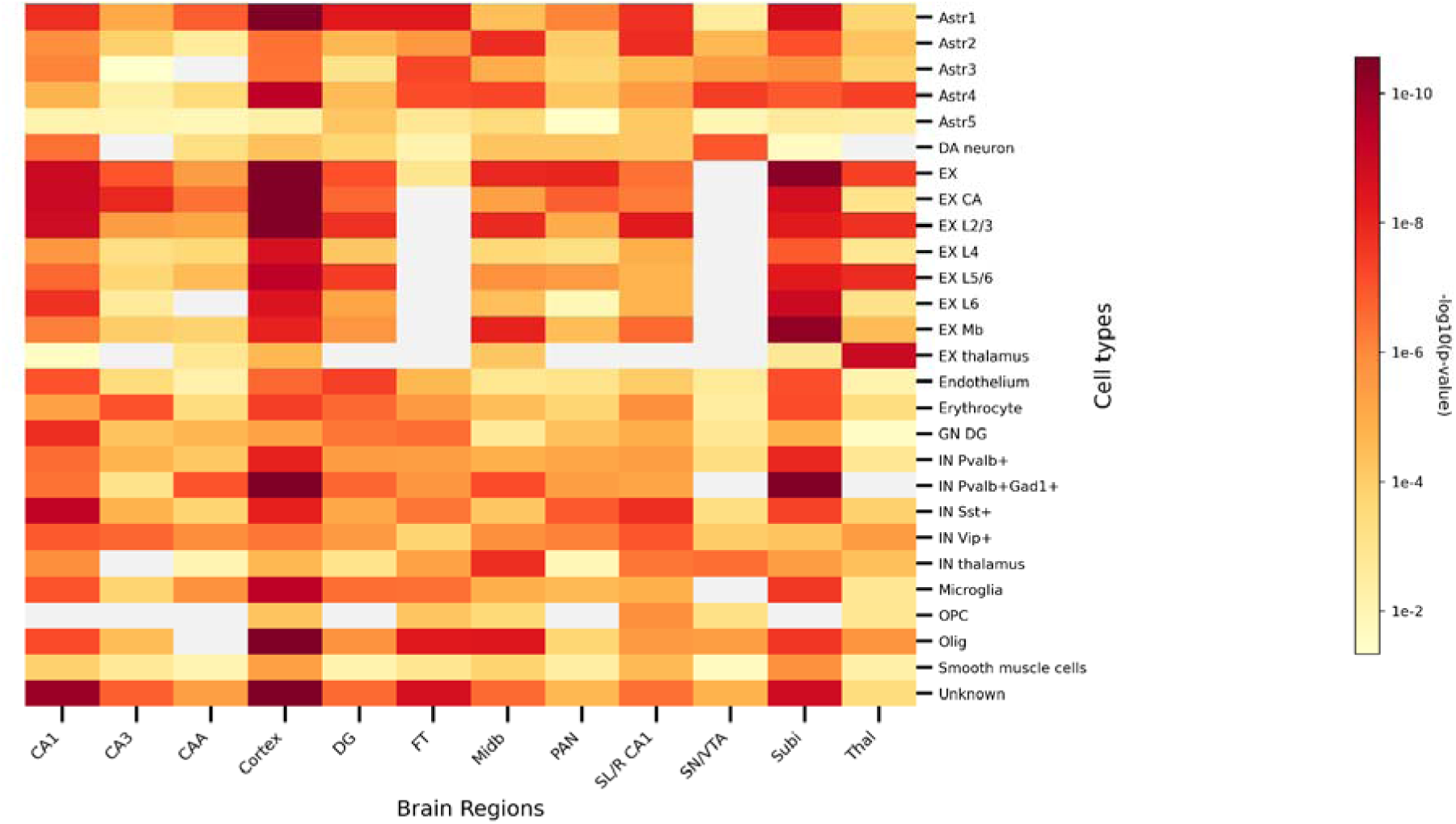
Spatial and cell-type–specific enrichment of ET-associated genetic loci across the adult mouse brain. Heatmap illustrating −log10-transformed p-values from the gsMap analysis, representing the spatial association between ET GWAS loci and transcriptomic signatures across anatomically defined brain regions and transcriptionally characterised cell types. Darker colours indicate stronger genetic enrichment (lower p-values), while white areas represent nonsignificant associations (p > 0.05). Based on the anatomical localisation, brain regions were grouped into: Cortex; Hippocampus CA fields (CA1, CA3, and CAA); DG, Dentate Gyrus; Subi, Subiculum; Thal, Thalamus; Midb, Midbrain; SN/VTA, Substantia Nigra/Ventral Tegmental Area; FT, Frontal Telencephalon; PAN, Pons; and SL/R CA1, Septal Nucleus/Layer CA1. Cell types were annotated based on canonical marker expression and include excitatory neurons (EX), inhibitory neurons (IN), astrocytes (Astr), dopaminergic neurons (DA), microglia, oligodendrocytes (Olig), oligodendrocyte progenitor cells (OPC), endothelial cells, erythrocytes, smooth muscle cells, and unclassified cells (“Unknown”).

To explore gene-level spatial correspondence, we next examined patterns of gene expression alignment with ET-associated enrichment using the gsMap-derived PCC. The genes with the strongest spatial correspondence included *PRKCB* and *SYT1* in excitatory neuronal populations, particularly the EX L5/6 subtype, mirroring the enrichment observed in hippocampal and cortical regions. Other genes showing similar spatial alignment included *SNAP25, ATP2A2* and *SLC24A2*, which are involved in synaptic signalling and neuronal excitability (**Figure 7**). Within fast-spiking inhibitory interneurons, *SCN1B* showed the strongest spatial correspondence, while *MDH1* was most correlated in Pvalb+ interneurons. In oligodendrocyte precursor cells (OPCs), *OPCML* demonstrated the highest PCC values, suggesting co-localisation with ET-relevant regions. Consistent with the gsMap framework, PCC highlights co-distribution of gene expression with ET-associated spatial signals and should not be interpreted as evidence of direct functional involvement (**Figure 7, Supplementary Figures 16-31**).

**Figure 7.**
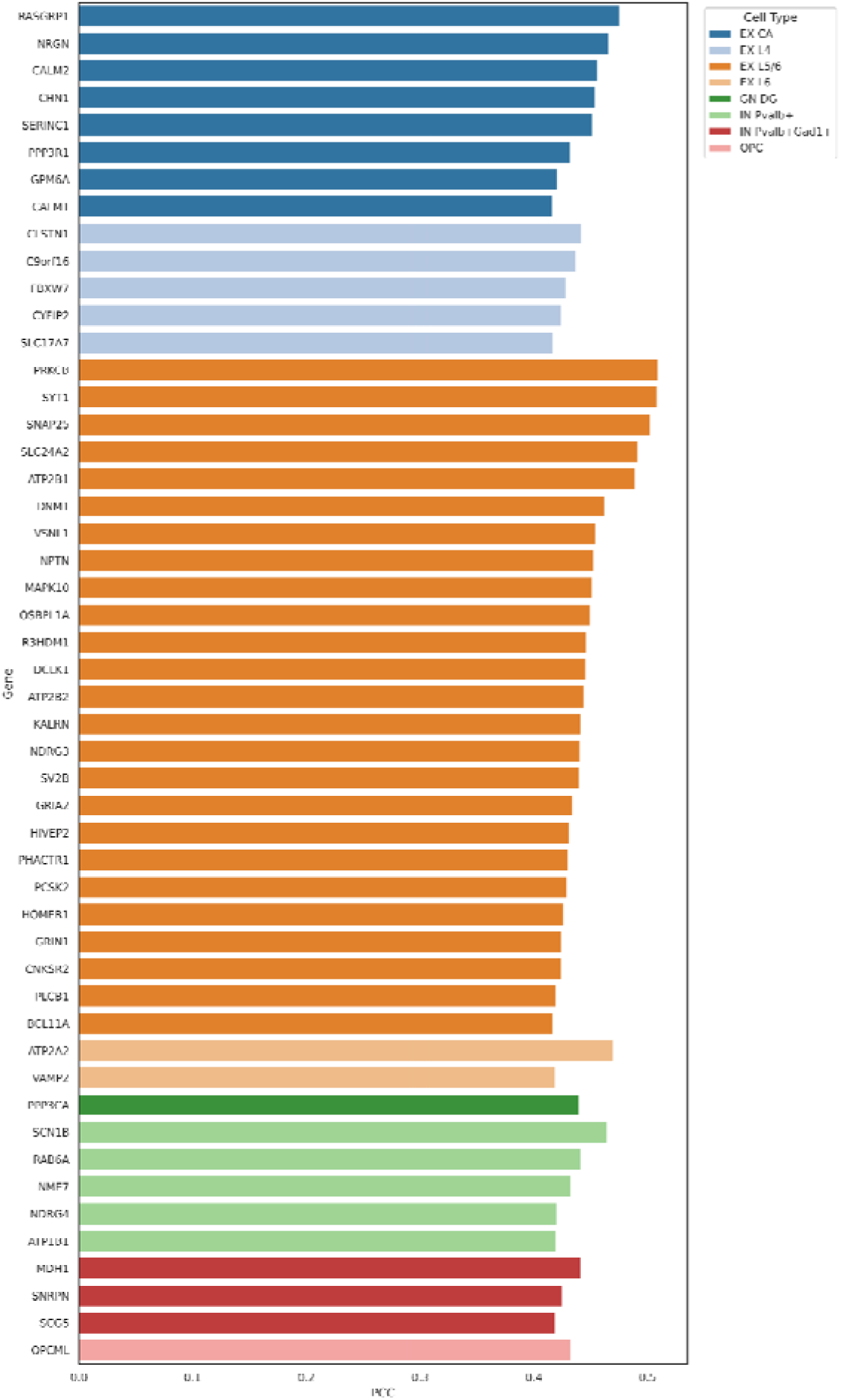
Top genes with the highest PCC by brain cell type. For each cell type, the gene with the strongest association in gene expression with ET was selected based on its PCC value. Bars represent the highest PCC per cell type, and the gene names are displayed at the end of each bar. Colours represent a continuous gradient based on PCC magnitude, with darker tones indicating stronger correlation. Gene expression data were derived from adult mouse brain cell types, and PCC values were calculated based on gene–trait similarity scores. Cell types: EX, excitatory; IN, Interneurons; GN DG, Granule neurons of the Dentate Gyrus; OPC, Oligodendrocyte Progenitor Cells; Astr, Astrocytes; DA, Dopaminergic neuron; Olig, Oligodendrocytes.

### Polygenic risk score (PRS)

Among UKB participants of EUR ancestry included in this study, 872 individuals (50.1%) were male, and the mean age at recruitment was 61 years. In the AoU cohort, 166 (32.9%) of participants with AFR ancestry and 151 (40.1%) with AMR ancestry were male. The mean age at recruitment was 66.1 years and 64.9 years, respectively.

In the UKB EUR cohort, PRS were computed using 7,356,519 variants shared between the SBayesRC weight-corrected summary statistics and the target dataset. After fitting a general linear model, the results showed that the PRS were a significant predictor of ET (OR per SD = 2.15, SE = 0.49, p < 2 × 10^-15^). The analysis of the PRS deciles in this group indicated an increase in ET prevalence in higher PRS deciles (**Figure 8**). Moreover, the PRS displayed modest discriminatory performance between cases and controls (AUC = 0.69). Further analysis of the association between the PRS and age at diagnosis in EUR showed that the mean age at diagnosis was approximately one year earlier in cases in the top decile compared to those in the lowest decile, as shown in **Supplementary Figure 32.**

**Figure 8.**
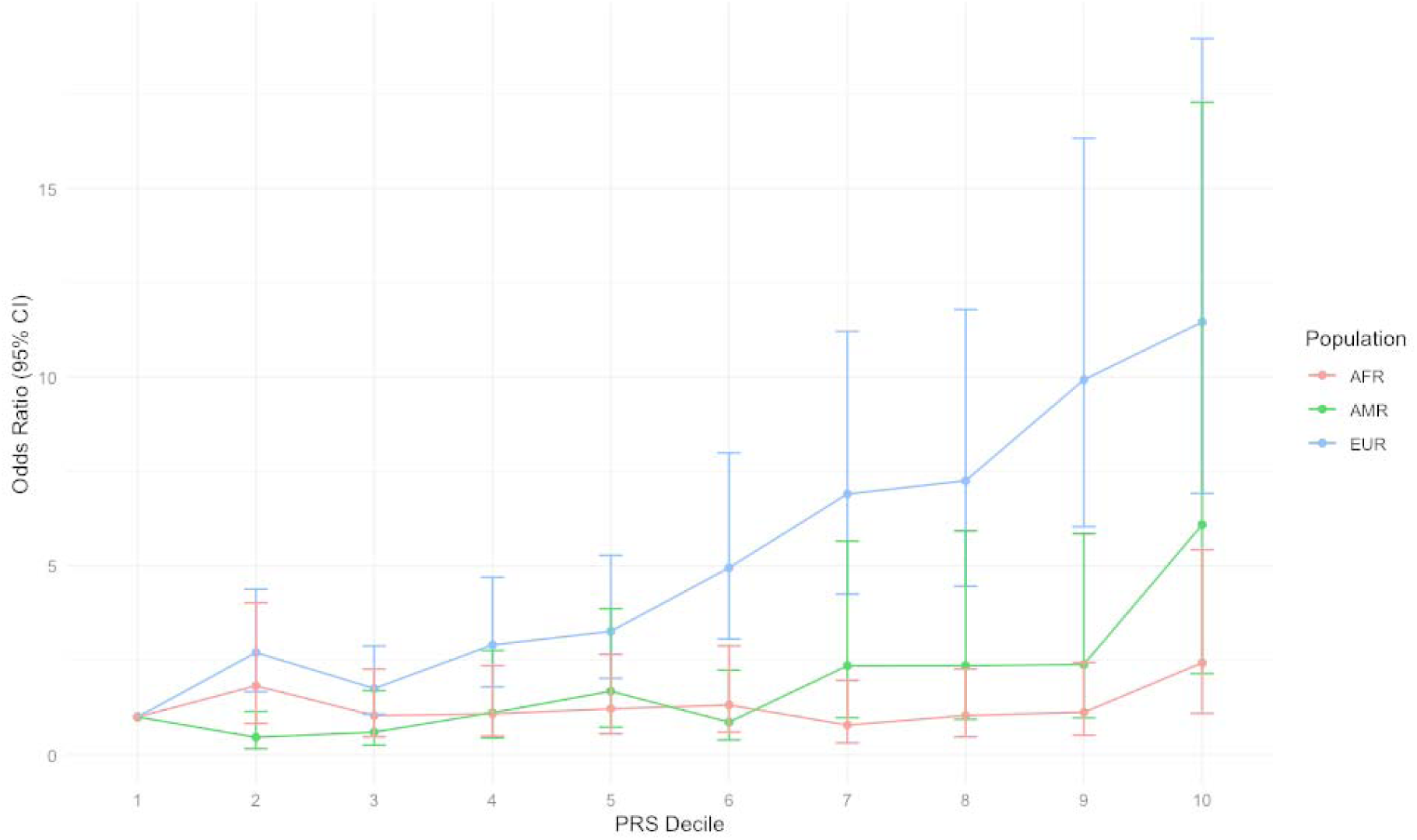
Polygenic Risk Score by population. In the EUR cohort, ET prevalence was 73.56% in the top PRS decile compared with 19.54% in the lowest, corresponding to an OR of 11.45 (CI 6.92 - 18.96). In the AFR cohort, ET prevalence was lower in the top decile (44.0%) than in the bottom decile (66.7%), resulting in an OR of 2.43 (CI 1.09-5.42). In the AMR cohort, prevalence increased from 40.5% in the lowest decile to 81.6% in the highest, with an associated OR of 6.08 (CI 2.14 - 17).

In the AFR and AMR cohorts from AoU, 3,647,795 and 4,097,301 variants, respectively, were included in PRS construction. Results from the general linear model indicated that the ET PRS was significantly associated with ET status in the AMR cohort (OR per SD = 1.99, SE = 0.12; p = 3.8 × 10CC). In the AFR cohort, the association showed a consistent direction of effect, although it did not reach statistical significance (OR per SD = 1.17, SE = 0.1; p = 0.09). A comparison of PRS deciles showed a stepwise increase in ET prevalence across both ancestry groups, as illustrated in **Figure 8**. AUC analysis indicated moderate predictive utility of the PRS in AMR (AUC = 0.67) and a low performance in AFR (AUC = 0.52).

## DISCUSSION

This genome-wide association meta-analysis represents the largest genetic study of ET to date, combining data from 20,268 individuals with ET and 723,761 neurologically healthy controls across three major European cohorts. We identified 50 independent genome-wide significant loci associated with ET, including 47 that have not been reported previously, substantially expanding the catalogue of ET risk loci. These findings provide a robust genetic framework for understanding ET and offer a valuable reference set for future research into disease mechanisms and potential interventions.

Our study revealed important insights into the genetic architecture of ET. We estimated SNP-based heritability at 24% on the observed scale (h² = 0.24, SE = 0.02) and 18.5% on the liability scale, assuming a population prevalence of 5%. These estimates are consistent with those reported for other complex disorders, including PD, restless legs syndrome, and bipolar disorder^19,20^, highlighting the substantial contribution of common variants to ET risk. The LD score regression intercept and attenuation ratio suggest that the observed genomic inflation is primarily driven by polygenicity rather than confounding factors such as population stratification or cryptic relatedness. This interpretation is further supported by the genomic inflation factor λGC of 1.37 and a mean chi-squared statistic of 1.44, both of which are indicative of a highly polygenic trait. We observed strong genetic correlations across cohorts, with near-complete concordance between MVP and 23andMe (r_G_ = 1) and robust correlations between these and the All of Us dataset. These findings underscore the consistency of ET’s genetic architecture across large, independent samples, supporting the validity of our meta-analytic approach.

Based on functional annotation, we identified several genes and biological pathways related to synaptic transmission, cerebellar development, and neuroinflammation with potential biological relevance in the pathogenesis of ET. Among 50 genome-wide significant loci, 47 were not previously associated with ET; the majority mapped to non-coding regions, including 29 intronic and 16 intergenic regions, as well as two regulatory regions and one variant in the 3’ untranslated region (UTR) and another in the 5’ UTR. Several genes surpassed the multiple testing threshold for genome-wide gene-level significance, further supporting the role of altered neuronal signalling and immune modulation in ET. One of the most significant novel loci identified in our study is a missense variant in *PPM1J* (rs78366259, P = 3.21 × 10□¹□). This gene has not been previously implicated in ET or other movement disorders. It encodes a serine/threonine phosphatase belonging to the PP2C family, which participates in cellular stress responses, regulates the cell cycle, and apoptosis^21^. Since ET may involve neurodegenerative or dysfunctional processes affecting Purkinje cells in the cerebellum, variants in phosphatases such as *PPM1J* might influence neuronal resilience to stress or signal transduction in motor pathways. We also replicated previously reported ET-associated loci^22,23^, including *EHBP1* rs74389013, *MIR924HG* rs1945016, and *LINC00323* rs9980363, providing further validation of these associations^22^. Of the four familial ET linkage loci previously reported, two (3q13^24^ and 5q35^25^) demonstrated concordance with genome-wide significant loci identified in this study, mapping in proximity to *ZBTB20* and *SQSTM1*^26^, respectively. This overlap reinforces the relevance of these chromosomal regions in ET susceptibility across both family-based and population-based analyses. The first locus linked to ET, referred to as *ETM1/FET1*, was initially reported in Icelandic families and mapped to the dopamine receptor D3 (*DRD3)* gene region on chromosome 3q13. Although early studies suggested that the *DRD3* rs6280 variant might increase the risk of ET and lower the age at onset, subsequent replication efforts yielded inconsistent results across populations, and other meta-analyses have not confirmed a significant association. These replications support both the robustness of our findings and the relevance of these loci across diverse populations.

On the other hand, six loci have been associated with a range of different complex traits. For instance, *SPATA3H1/GCKR* rs1260333 has been linked to metabolic traits such as triglyceride and cholesterol levels; *MAP3K1* rs7731700 to body weight; *ATOH7* rs7916697 to ocular and anthropometric phenotypes, including glaucoma and optic disc area; *MY019/GGNBP2* rs12941866 and *SSBP4* rs11673604 to neuropsychiatric and immune-related disorders; and *CALN1* rs34858520 to physical activity. This overlap suggests that ET shares aspects of its genetic architecture with metabolic regulation, anthropometric variation, and neurodevelopmental or neuropsychiatric processes. Such pleiotropy highlights potential biological connections between systemic metabolic pathways, brain development, and motor circuit vulnerability, which could be explored in future mechanistic studies.

Many novel loci overlap genes involved in neurodevelopmental and motor function pathways, including *PPARGC1A, CACNA1A*, and *YWHAE*, which are known to play roles in mitochondrial function, calcium signalling and synaptic plasticity, respectively. These findings expand the candidate gene catalogue, suggesting both neurodevelopmental and neurophysiological processes contribute to ET pathogenesis. Notably, *PPARGC1A* encodes PGC-1α, a transcriptional coactivator that regulates mitochondrial function and oxidative stress responses, processes critical for the survival and function of cerebellar neurons^27^. While previous studies identified rs17590046 as the lead SNP within this locus^28^, our GWAS meta-analysis detected strong associations with rs17590529 and rs74419502, which are located in the same genomic region. This convergence underscores the robustness of the association and suggests that common variation in *PPARGC1A* may contribute to ET pathogenesis. Similarly, previous reports have highlighted *STK32B* and *CTNNA3* as additional susceptibility loci, although these were not among the top signals in our dataset^28^. Moreover, *CACNA1A* encodes the pore-forming α1 subunit of the P/Q-type voltage-gated calcium channel, a critical mediator of neuronal calcium influx whose dysfunction is associated with a clinically heterogeneous spectrum of disorders, including episodic ataxia, cerebellar ataxia, and neurodevelopmental syndromes with epilepsy and intellectual disability, underscoring its established role in cerebellar excitability and motor control^29^. Consistent with our findings, transcriptomic analyses have shown that *CACNA1A* is downregulated in ET cerebellar tissue. Pharmacological studies report that calcium-channel blockers exacerbate tremor in ET patients, highlighting the functional importance of calcium homeostasis in tremor generation^30^. Finally, *YWHAE* encodes 14-3-3ε, a protein highly expressed in the brain and involved in intracellular signalling and synaptic integrity^31^. These associations suggest that dysregulated neuronal calcium homeostasis, mitochondrial dysfunction, and synaptic vulnerability contribute to the development of ET.

Beyond inter-cohort consistency, this study highlights the broad pleiotropic architecture of ET. Genetic correlation analyses with 2,871 complex traits identified 128 significant associations after multiple testing corrections. Notably, several neurological phenotypes, including PD, abnormal involuntary movement and myoclonus, displayed strong positive genetic correlations with ET, supporting prior epidemiological observations of shared clinical features and possible overlapping pathophysiology. In particular, the high genetic correlation with PD (r_G_ = 0.49) reinforces the hypothesis of partially convergent mechanisms involving motor circuits and neurodegenerative processes^22,23,32^. Moreover, significant correlations with cardiovascular, pain-related, musculoskeletal, and metabolic traits suggest that ET may share etiological pathways with systemic conditions, potentially implicating common inflammatory or vascular processes^33–36^. These findings indicate that ET is not only a neurological disease but may also be influenced by systemic biological processes, warranting further investigation into potential shared mechanisms and comorbidities^37,38^. In parallel, the genetic correlation analysis with neuroimaging phenotypes revealed significant inverse associations with volumes of the ventral diencephalon and multiple cerebellar subregions. The cerebellum has long been implicated in ET pathophysiology^39^, as evidenced by both post-mortem and neuroimaging studies. The observed negative correlations support the notion that genetic predisposition to ET may be linked to smaller regional volumes. The localisation of these effects to inferior posterior lobules and flocculonodular regions is consistent with recent work implicating cerebellar circuits in tremor generation and modulation^39^. The absence of significant correlations with cortical surface area and thickness measures suggests that ET-associated genetic variation may preferentially affect subcortical and cerebellar structures rather than cortical morphometry.

To better understand the biological pathways involved in ET, we performed TWAS, identifying *BACE2, RP11*L*344P13.6* and *ZKSCAN* genes, whose genetically predicted expression was significantly associated with ET pathophysiology. Among the tissues examined, the most significant transcriptional associations were observed in the cerebellum, basal ganglia (including the caudate, putamen, and nucleus accumbens), substantia nigra, and the cervical spinal cord C-1, which are core regions involved in motor coordination and tremor generation. *BACE2* showed compelling associations in spinal cord cervical C-1 and substantia nigra tissues, aligning with a novel cell-type specificity. A single-cell eQTL atlas of the human cerebellum demonstrated that ET risk variants at the *BACE2* locus are causally linked to its downregulation in immature oligodendrocytes, suggesting selective vulnerability and possible demyelination processes in ET pathology^40^.

Our fine-mapping outcomes further support the cerebellum as a central hub in ET pathophysiology^41,42^. The repeated prioritisation of cerebellar genes such as *TRIM50, TSPAN15, WDPCP,* and *XYLT1* is consistent with long-standing neuropathological and imaging evidence implicating cerebellar circuitry dysfunction in ET^42–44^. These genes are involved in diverse processes, including cytoskeletal organisation, synaptic architecture, and cell adhesion, that might influence Purkinje cell integrity and cerebellar network stability^45,46^. The identification of *HARS* and *CCDC125* in the hypothalamus and *CTH* in the hippocampus points to additional, potentially under-recognised neuroanatomical contributors to ET, such as metabolic regulation and limbic-motor integration^47^. Moreover, *HARS*, an aminoacyl-tRNA synthetase, showed colocalisation in both cortex and cerebellum, pointing to potential disruptions in protein translation and neuronal maintenance in ET. Mutations in *HARS* have been linked to neuropathies, suggesting a plausible mechanistic link to tremor phenotypes through axonal degeneration or synaptic dysfunction^48^. The presence of *SNX17* and *CCNT2* in the putamen and *GRK7* in the spinal cord cervical C-1 suggests that the genetic risk for ET extends beyond the cerebellum, potentially involving broader motor and sensorimotor networks^49^. Together, these findings reinforce the concept that ET is not exclusively a cerebellar disorder but rather the product of a distributed network pathology with region-specific molecular vulnerabilities.

Our colocalisation analysis identified *S100A13, BACE2, PCDH9*, and *HARS* as causal genes underlying ET risk. The near-complete colocalisation of *S100A13* across most brain regions suggests a ubiquitous role in ET biology. *S100A13* encodes a calcium-binding protein involved in angiogenesis, stress response, and neuroprotection, with altered expression previously reported in neurodegenerative contexts^50^. Its widespread colocalisation may indicate a core role in maintaining neuronal and vascular homeostasis across multiple brain networks vulnerable in ET. *PCDH9*, with strong cerebellar colocalisation, encodes a protocadherin involved in synaptic adhesion and dendritic spine regulation^51^. Its implication aligns with converging evidence that synaptic connectivity alterations within the cerebello-thalamo-cortical circuit are central to ET pathogenesis.

Our spatial transcriptomic integration provides mechanistic granularity to the growing view that ET arises from distributed circuit dysfunction rather than a singular cerebellar lesion model. The pronounced enrichment of genetic risk within excitatory pyramidal neurons of cortical and hippocampal layers, particularly EX L2/3 and EX L5/6 subtypes, is consistent with converging neuroimaging and electrophysiological evidence showing altered functional connectivity between the motor cortex, hippocampus, and cerebellum in patients with ET^52–54^. Recent evidence also highlights the existence of distinct excitatory neuron subtypes within hippocampal fields that support long-range information transfer and behavioural modulation^55^. These findings suggest that ET genetic risk may converge on transcriptional programmes active in cortico-hippocampal projection neurons involved in motor coordination, sensorimotor integration, and limbic-cortical communication.

The intrinsic vulnerability of these long-range projection neurons, characterised by high metabolic demand, elaborate dendritic arborisation, and sustained firing, has been implicated in other neurodegenerative disorders, suggesting that similar mechanisms such as impaired axonal transport, synaptic vesicle trafficking, and calcium homeostasis may contribute to tremor pathogenesis^56,57^. In our analysis, several genes that show strong spatial correspondence with ET-enriched regions, including *PRKCB, SYT1* and *SNAP25*, are known to regulate synaptic transmission and plasticity. However, consistent with the gsMap framework, these gene-level associations reflect spatial co-localisation rather than direct evidence of functional causality. Thus, they should be interpreted as supportive rather than definitive mechanistic candidates.

Moderate but consistent enrichment was also observed in glial populations, including astrocytes, microglia, and oligodendrocyte progenitor cells (OPCs), aligning with post-mortem and transcriptomic evidence of altered myelination, gliosis, and neuroimmune activity in ET^2,58,59^. The spatial correspondence of *OPCML* in OPCs may indicate impaired oligodendrocyte maturation and axonal insulation, potentially affecting the temporal precision of cerebello-thalamo-cortical transmission. Enrichment in microglia, particularly within subcortical and hippocampal regions, reinforces evidence for immune-mediated synaptic remodelling in tremor and related movement disorders^60–63^

In contrast, enrichment within inhibitory interneurons (Pvalb+, Sst+) was relatively restricted, suggesting that GABAergic tone may be partially preserved, consistent with electrophysiological observations^64^. This pattern supports a model in which ET risk influences excitatory network instability, with inhibitory systems exerting a compensatory or modulatory role rather than a primary pathogenic one^65^.

Importantly, gsMap enabled the anchoring of GWAS risk loci to anatomically and transcriptionally defined cell populations, bridging the gap between statistical association and functional neurobiology. When considered alongside our TWAS, fine-mapping, and colocalisation evidence, highlighting genes such as *HARS, BACE2*, and *S100A13*, these findings support a multi-cellular, regionally distributed model of ET involving excitatory neuronal susceptibility and glial circuit modulation. Such insights open avenues for targeted therapeutic strategies aimed at enhancing synaptic resilience, modulating glial support functions, and stabilising excitatory network dynamics in tremor circuits.

In addition to genome-wide and cross-trait correlation analyses, we tested the capacity of our findings to inform individual-level risk stratification through PRS. PRS derived from our meta-analytic GWAS demonstrated significant predictive utility for ET diagnosis in both the EUR and AMR cohorts. Predictive performance was strongest in the EUR cohort, consistent with its larger sample size and greater genetic similarity to the discovery population. In the AMR cohort, despite smaller numbers, PRS remained significantly associated with ET, likely reflecting shared European admixture that enhances portability^66^. In contrast, no significant association was observed in the AFR ancestry cohort, despite a larger sample size than the AMR cohort, underscoring the persistent challenge of cross-ancestry PRS transferability, which is attributable to divergent linkage disequilibrium patterns, allele frequencies, and the under-representation of non-European populations in discovery datasets. Nonetheless, a notable number of ET cases was observed in the top PRS decile relative to the bottom decile within the AFR group (OR = 2.54, p = 2.3 × 10□²), indicating a detectable genetic signal despite reduced statistical power. We also observed an association between higher PRS values and earlier age of onset in the EUR cohort. This suggests that individuals with a higher genetic liability to ET may be more likely to develop symptoms at an earlier age. This trend has been previously reported in other disorders such as PD^67^, supporting a potential role for PRS in informing early diagnosis through risk stratification, particularly in well-characterised EUR datasets.

Despite these promising findings, important limitations remain. While PRS showed significant predictive utility in the EUR cohort, the moderate effect sizes suggest that current models may be influenced by residual phenotype misclassification and the inherent complexity of differentiating ET from related movement disorders in population-scale datasets. A plausible explanation is that, given the clinical heterogeneity of ET and the potential overlap with related phenotypes, even strict diagnostic criteria may lack the specificity required for precise case definition. Such phenotypic imprecision could dilute genetic signals and contribute to reduced PRS accuracy. The modest SNP-based heritability of ET constrains current PRS models, and their predictive value will likely need to be enhanced through integration with additional clinical and environmental risk factors.

Furthermore, ancestry-related disparities in predictive performance highlight the need for more inclusive GWAS with greater representation of non-European populations. The development and application of ancestry-aware or multi-ancestry PRS models will be essential to ensure equitable utility of polygenic prediction. Addressing these classification, heterogeneity, and representation challenges, both at the diagnostic and discovery stages, will be critical to improving PRS performance and ensuring their equitable application across ancestries.

In summary, this meta-analysis substantially advances the genetic characterisation of ET, expanding the number of genome-wide significant loci to 50 and implicating biological pathways centred on cerebellar circuitry, excitatory neuronal function, and glial support. Integration with transcriptomic, fine-mapping, and spatial analyses converges on a distributed network model of ET pathophysiology, extending beyond a purely cerebellar paradigm. PRS demonstrates predictive potential, particularly within European populations, but also underscores the persistent challenges of cross-ancestry portability, phenotypic heterogeneity, and limited heritability capture. Addressing these challenges through larger, ancestrally diverse cohorts, refined phenotyping, and multi-omic integration will be crucial for translating genetic discoveries into reliable risk stratification, mechanistic insight, and, ultimately, targeted interventions for ET.

## METHODS

### Sample information and phenotype ascertainment

This study included genome-wide association study (GWAS) data from three large-scale cohorts: the Million Veteran Program (MVP), the All of Us (AoU) Research Program, and 23andMe Research Institute. The total sample size across all cohorts included 20,268 individuals with ET and 723,761 neurologically healthy controls. Case status was defined using self-reported diagnosis of ET or corresponding International Classification of Diseases (ICD) codes, where available. Controls were individuals with no known neurological disorders.

#### All of Us Research Program (AoU)

A case-control GWAS was conducted in the AoU Cohort using the Data Release 8 version (February 2025). The analysis included 2,960 cases and 221,377 controls with short-read whole-genome sequence data, all of European ancestry (with a predicted European ancestry of greater than 60%). In addition, participants who failed genetic QC and those with discordance between their self-reported sex at birth and genetic sex were excluded. Cases were defined using SNOMED code 609558009 for ET, while controls excluded participants with a history or diagnosis of any tremors. Regenie v3.2.6^68^, which accounts for sample relatedness, was used for the analysis. The logistic regression model included covariates for age, sex, and the first 10 ancestral principal components (PC). Variants with a MAC <20 and later on MAF <1% were excluded from the analysis.

#### Million Veteran Program (MVP)

Publicly available GWAS summary statistics for ET were obtained from Verma et al.^32^, comprising 13,900 ET cases and 614,227 controls, all of European ancestry. Participants were genotyped and imputed according to the protocols described in Verma et al^32^. Standard quality control (QC) procedures were applied, including exclusion of variants with imputation quality <0.3 and minor allele frequency (MAF) <1%.

#### 23andMe

GWAS summary statistics from the 23andMe cohort were obtained through an application to the 23andMe Publication Dataset Access Program (https://research.23andme.com/dataset-access/) and formalised through a Data Transfer Agreement. The dataset comprised 3,408 self-reported ET cases and 65,772 controls, all of whom were of European ancestry. Participants provided informed consent and were genotyped using custom arrays. Individuals under the age of 30 or those genotyped on the v1 platform were excluded. Genotyping, imputation, and quality control procedures were carried out as described in previous 23andMe publications^22^. Ethical and Independent Review Services approved the study protocol, which was reviewed by an institutional review board accredited by AAHRPP.

### ET GWAS meta-analysis

An inverse-variance-weighted fixed-effects meta-analysis was conducted across the three cohorts using METAL^69^. All GWAS results were harmonised to ensure allele alignment and strand orientation before the meta-analysis. Studies were weighted by effective sample size to account for differences in case-control ratios. Only variants present in at least two cohorts and passing cohort-specific QC thresholds were included. Final summary statistics were used for downstream analyses, including genetic correlation and pathway enrichment.

### Cortical, subcortical and cerebellar GWAS data

To investigate brain regions implicated in ET, we utilised publicly available GWAS summary statistics for the volume of nine subcortical nuclei and eight cerebellar regions, as well as surface area and cortical thickness of 34 cortical areas, and intracranial volume.

GWAS summary statistics for cortical regions were obtained from the Oxford Brain Imaging Genetic Server-BIG40^70^, which included hemisphere-independent (i.e., bilateral) MRI brain measurements from 33,224 individuals of European ancestry based on UK Biobank data. GWAS summary statistics for subcortical volumes were obtained from a large-scale meta-analysis that included participants from ENIGMA-CHARGE, UK Biobank and the ABCD cohort (n = 41,362), all of European ancestry^71^. GWAS data for the cerebellum were derived from T1-weighted MRI brain volume measurements in 33,265 individuals of European ancestry from the UK Biobank, as reported by Chambers et al^72^. Grey matter volumes were extracted using FAST segmentation. The following cerebellar regions were assessed bilaterally and at the vermis when available: (1) anterior lobe (lobules I–V), (2) superior posterior lobe (hemispheric lobules VI–Crus I), (3) superior posterior vermis (vermal lobules VI–Crus I, excluding Crus I vermis), (4) inferior posterior lobe (hemispheric lobules Crus II–IX), (5) inferior posterior vermis (vermal lobules Crus II–IX), (6) flocculonodular lobe (hemispheric lobule X), (7) flocculonodular vermis (vermal lobule X), and (8) total cerebellar volume (excluding Crus I vermis)^72^.

### Functional annotation via FUMA and MAGMA

Post-GWAS analyses were conducted using the FUMA online platform v1.3.7 to perform functional annotation across the entire genome. A gene-based association study was conducted using Multi-marker Analysis of Genomic Annotation (MAGMA)^73^. This method calculates gene-based P-values by evaluating the combined effects of all SNPs within a gene while taking into account linkage disequilibrium (LD) among them. SNPs were mapped to genes using the human genome assembly Build 37 (GRCh37/hg19), with a focus on 19,107 protein-coding genes. Gene boundaries were extended by ±10 kb to include regulatory regions, aligning with the default mapping parameters of FUMA. To address multiple tests, we applied corrections based on the number of protein-coding genes, setting the significance threshold at P = 0.05 / 19,107 = 2.62 × 10□□.

### Conditional-joint analysis to nominate variants of interest

To identify which of the genome-wide significant variants identified in this study are independent (i.e., not in LD) from variants found in the ET GWAS by Skuladottir et al.^23^, we performed a conditional analysis to account for the 12 previously reported variants. We used conditional and joint analysis (GCTA-COJO v 1.91.7, http://cnsgenomics.com/software/gcta/), an approach that estimates the independent effects of SNPs by conditioning on other associated variants in LD, thereby identifying signals that represent true independently associated loci^74^. Summary-level statistics from the European-ancestry meta-analysis of ET were conditioned using independent signals from the previous larger ET meta-analysis in Europeans^23^, and an LD reference panel based on five thousand healthy individuals from UKBB (European subset, GRCh37 with 40510321 number of variants). Variants were restricted to MAF ≥ 0.01 and high-quality SNPs present in both the GWAS summary data and the LD reference.

### SNP-based heritability and genetic correlation via LDSC

We performed univariate Linkage Disequilibrium Score Regression (LDSC) to estimate the SNP-based heritability (h^2^_SNP_) of the ET GWAS meta-analysis. SNP-based heritability represents the proportion of phenotypic variance in a trait that can be attributed to common genetic variants included in the analysis. Next, we used LDSC to calculate the genetic correlation (r_G_) between ET and 2,871 phenotypes (with available GWAS summary statistics) using CTG-VL^75^ based on a common set of HapMap3 variants. We used the Benjamini–Hochberg false discovery rate (FDR) at 5% to define statistical significance. To ensure robust and interpretable results in the r_G_ analyses, we applied a systematic trait filtering procedure. First, we retained only those phenotypes that showed a statistically significant r_G_ with ET after using a 5% FDR correction for multiple testing. Next, we removed duplicate traits and consolidated overlapping or redundant entries. We also excluded traits that were not directly relevant to the phenotype under investigation, such as those referring to family members’ diagnoses (e.g., traits related to parents or siblings) or overly broad or vague phenotypes. Additionally, traits with limited biological interpretability in the context of neurological or motor disorders (e.g., dietary preferences or general lifestyle factors) were removed to focus the analysis on meaningful and disease-relevant associations.

We further calculated r_G_ between ET and 68 neuroanatomical regions derived from neuroimaging GWAS summary statistics, encompassing cortical, subcortical, and cerebellar areas. Genetic correlation quantifies the proportion of gene variance shared between ET and each brain region, normalised by the square root of the product of their respective SNP-based heritability estimates. To account for multiple comparisons, we applied a 5% FDR correction independently within each anatomical group (cortical, subcortical, and cerebellar). This robust approach accounts for potential sample overlap and provides insights into the shared genetic architecture between ET and each regional brain category^76^.

### Transcriptome-wide association studies (TWAS)

TWAS was conducted to identify gene expression-mediated associations with ET across 13 brain tissues, including the cervical spinal cord. Associations between genetically predicted gene expression and ET risk were estimated using S-PrediXcan^77^, a summary-statistics–based method that integrates SNP-level effect sizes with tissue-specific gene expression prediction models while adjusting for local LD. We used elastic net-based prediction models pretrained on GTEx v8 and their corresponding covariance matrices, obtained from the PredictDB data repository (http://predictdb.org/). The input GWAS summary statistics were derived from a meta-analysis of European ancestry cohorts (20,268 cases and 723,761 controls), harmonised to the GRCh38 genome build. The S-PrediXcan analysis was conducted separately for each brain tissue, and tissue-specific gene–trait associations were computed. To integrate evidence across tissues, we subsequently applied SMultiXcan^78^, a multivariate extension of S-PrediXcan that aggregates single-tissue TWAS results by leveraging cross-tissue variation in genetically regulated expression. This approach increases power by capturing shared genetic effects across tissues and was implemented using our tissue-specific S-PrediXcan results. LD-aware filtering was applied to exclude components with high condition numbers (>30) to ensure numerical stability. Multiple testing was addressed using Bonferroni correction based on the number of genes tested across tissues, with significance defined as p < 0.05 / N. All analyses were conducted using the MetaXcan framework (v0.6.0; https://github.com/hakyimlab/MetaXcan). Input specifications included SNP ID, alleles, effect size (β), standard error (SE), and Z-score.

### Fine-mapping with FOCUS

We applied FOCUS (Fine-mapping Of CaUsal gene Sets) version 0.9 to perform probabilistic fine-mapping of TWAS associations^79^. FOCUS estimates the posterior inclusion probability (PIP) that each gene is causally linked to the trait, accounting for correlation between gene expression prediction models (i.e., eQTL weights) due to LD. We ran FOCUS in single-population mode using precomputed prediction weights derived from GTEx v8 brain tissues and the genotypes from the European subset of the 1000 Genomes Project Phase 3 reference panel to model LD. Input summary statistics were harmonised to GRCh38 coordinates. For each locus, FOCUS produced posterior inclusion probabilities, credible gene sets achieving 90% cumulative PIP, and locus-level model fits. Genes with PIP >0.9 were considered highly likely to be causal contributors to the observed association signals.

### Colocalisation with COLOC

We assessed the co-occurrence of ET risk and brain-tissue-specific eQTL signals using the COLOC package^80^ with default parameters in R. For each gene identified as significant by SMultiXcan, we extracted GWAS variants within 1 MB up- and downstream of its transcription start site. We colocalised these signals with harmonised eQTL summary statistics mapped to these genes, retrieved from the eQTL Catalogue^81^ for GTEx v8 brain and spinal cord tissues. We extracted the posterior probabilities for Hypothesis 4, which indicates a shared causal variant between the GWAS and eQTL signals. We considered a posterior probability > 0.8 as evidence of colocalisation.

### Integration of GWAS with Spatial Transcriptomics using gsMap

To investigate the spatial relevance of genetic associations in ET, we applied the gsMap framework to integrate GWAS summary statistics with spatial transcriptomic (ST) data from the adult mouse brain. gsMap quantifies the enrichment of heritability across spatially defined transcriptomic domains by combining graph neural networks (GNNs) and stratified linkage disequilibrium score regression (S-LDSC). First, a spatial graph of transcriptomic spots is constructed, and GNNs are used to learn low-dimensional embeddings that capture spatial proximity and transcriptional similarity. Homogeneous spot groups are identified using cosine similarity in the embedding space, and gene spot scores (GSSs) are calculated to reflect cell–type–specific gene expression. These scores are then mapped to SNPs based on genomic proximity, generating spot-level genetic annotations. Using S-LDSC, gsMap tests whether SNPs with higher GSSs are enriched for ET GWAS signals, thereby identifying spatially localised brain regions and transcriptionally defined cell types contributing disproportionately to ET heritability^82^.

In addition to spatial heritability enrichment, we evaluated spatial correspondence between ET-associated genomic architecture and gene expression using the Pearson correlation coefficient (PCC) as implemented in gsMap. For each gene, PCC was computed by correlating its expression pattern across Visium ST spots with the gsMap-derived enrichment scores representing ET genetic association across the same spatial coordinates. This analysis identifies genes whose spatial expression distributions align with ET-relevant anatomical domains. Consistent with the original gsMap framework, PCC was used as a descriptive measure of spatial co-localisation rather than as a functional gene prioritisation metric^82^.

### Polygenic Risk Score (PRS)

PRS for ET were constructed using the meta-analysed GWAS summary statistics generated in this study. Scores were calculated separately for three ancestry groups: Europeans (EUR) from UK Biobank (UKB), Africans (AFR) and Admixed Americans (AMR, predominantly Latino) from AoU.

#### UK Biobank

The EUR cohort was derived from the UKB, a population-based study that includes genetic and phenotypic data from approximately 500,000 participants aged 40 to 69 at recruitment (2006-2010)^83^. Genotyping was performed on over 480,000 participants and imputed using the Haplotype Reference Consortium (HRC)^84^ and UK10K^85^ panels. After quality control, 487,409 individuals were retained, of whom 438,870 were classified as of white British ancestry, following a PCA-based approach. SNPs with a missing rate greater than 5% were excluded, as were individuals with more than 5% missing genotypes, using PLINK. Further details on the sample quality control procedures have been published previously. ^86^. Variants with a MAF < 0.01 or an imputation quality score Rsq < 0.8 were excluded from the analysis. Diagnosis information for ET participants was based on ICD-10 criteria. Age at diagnosis was calculated using age at recruitment (f.21022), date of attendance at an assessment centre (f.53), and date of diagnosis (G20; f.131022). ET cases were selected based on the ICD-10 criteria (G25; f.41270). Participants with ICD-10 codes indicative of major neurological conditions were excluded to minimise confounding from comorbid or overlapping disorders. Specifically, we removed individuals with codes G10-G14 (hereditary ataxias and systemic atrophies), G20-G24 (Parkinson’s disease and other movement disorders), G30-G32 (degenerative diseases of the nervous system), G35-G37 (demyelinating diseases), and G80-G83 (cerebral palsy and other paralytic syndromes). The total number of ET cases after applying exclusion criteria was 870. The analysis included 870 age- and sex-matched controls. All models were adjusted for sex, age and population stratification (i.e., 10 PCs).

#### All of Us Research Platform

AFR and AMR were sourced from the AoU Research Program’s whole-genome sequencing dataset (short-read v8 “ACAF” callset)^87^. This callset includes variants with MAF > 1% or allele count > 100 per subpopulation. ET cases were defined using OMOP concept code 43531003; controls were matched by age and sex, excluding individuals with any neurological condition (ICD-10 chapter VI) reported. The final analysis included 258 unrelated cases and 258 controls (AFR) and 189 cases and 189 controls (AMR).

Quality control involved removing indels, multiallelic variants, and variants with QUALapprox < 60^88^, MAF < 1%, or > 5% missingness (--geno 0.05). Individuals with > 2% missing genotypes were also excluded (--mind 0.02). SNPs were updated to the rsID format using the dbSNP hg38 reference panel. We calculated the first 10 PCs of both target samples. Before computing PCs, we filtered out variants with MAF < 5% and performed LD pruning using Plink 1.9, with a 500 kb window and an R^2^ threshold of 0.2. Given the differences in LD structure and variant density, one round of pruning was performed for the AMR cohort, and two rounds for AFR.

### PRS construction

In UKB and AoU, PRS analyses were performed using SBayesRC^89^, which jointly estimates SNP effect sizes within LD blocks while integrating functional annotations (∼7 million SNPs). A EUR ancestry LD reference was used, as presented in the methods section (https://github.com/zhilizheng/SBayesRC).

In the UKB EUR cohort, SNP weights processed by SbayesRC were applied to individual genotypes using PLINK 2. The resulting scores were normalised using a rank-based inverse normal transformation. We then performed generalised linear regression analyses for ET PRS as the main predictor, adjusting for sex, age, the first 10 PCs, and genotyping batch as covariates. In an additional model, we assessed age at diagnosis as a dependent variable only for participants with an available age at diagnosis (based on ICD-10 ET cases) using ET PRS as the main predictor.

In the AoU AFR and AMR cohorts, the same SNP weights processed by SBayesRC were applied to each genotype set. We subsequently transformed the resulting PRS using the same rank-based inverse normal transformation. Moreover, for each ancestry group, we fit a generalised linear model using the PRS as the main predictor while adjusting for age, sex and the first 10 PCs.

## Acknowledgments

We would like to thank the research participants and employees of 23andMe Research Institute for making this work possible. We are also deeply grateful to all the participants of All of Us, whose invaluable contributions and commitment made this research possible. We also thank the National Institutes of Health’s All of Us Research Program for making available the cohort examined in this study as part of the ET GWAS meta-analysis and PRS construction section. Additionally, this work was conducted using the UK Biobank Resource (application number 25331). The UK Biobank was established by the Wellcome Trust, a medical charity; the Medical Research Council of the United Kingdom; the Department of Health of the United Kingdom; the Scottish Government; and the Northwest Regional Development Agency. It also had funding from the Welsh Assembly Government, British Heart Foundation, and Diabetes UK. MER thanks the support from the Rebecca L. Cooper Medical Research Foundation (F20231230). SM is supported by an NHMRC Investigator grant.

## AUTHOR’S CONTRIBUTIONS

NSO, SDT and MER were responsible for the original study idea. NSO, FC, VFO, SSM, MS, LS, and JK conducted the genetic data analysis. All authors contributed to data interpretation, critically reviewed each draft of the report, and approved the final submitted version.

## DATA AVAILABILITY

This study used data from the All of Us Research Program’s Controlled Tier Dataset version 8, available to authorised users on the Researcher Workbench. Downsampled GWAS summary statistics for this Essential Tremor GWAS (without 23andMe data) will be made available via the GWAS Catalog upon publication of this manuscript. Researchers interested in accessing summary statistics from the 23andMe cohort should follow the appropriate application procedure (https://research.23andme.com/dataset-access/).

## CODE AVAILABILITY

All source code is available at GitHub.

