## Supplementary Figures for "Genome-Wide Meta-Analysis Identifies 47 Novel Loci and Links Essential Tremor to Ventral Diencephalon and Cerebellum Morphometry"

**Contents**

This file contains the following supplementary information for the manuscript:

**Figure 1.** QQ plot of ET GWAS meta-analysis and ET gene-level association study

**Figure 2.** Gene enrichment across GTEx 53 samples

**Figure 3.** Manhattan plot of individual dataset

**Figure 4.** Regional plots of the loci associated with ET - Previously known associations with ET.

**Figure 5.** Regional plots of the loci associated with ET - Previously known associations.

**Figure 6.** Regional plots of the loci associated with ET - Novel associations.

**Figure 7.** Genetic correlation between ET and Neurological traits

**Figure 8.** Genetic correlation between ET and Cardiovascular traits

**Figure 9.** Genetic correlation between ET and Musculoskeletal traits

**Figure 10.** Genetic correlation between ET and Pain conditions

**Figure 11.** Genetic correlation between ET and Respiratory traits

**Figure 12.** Genetic correlation between ET and Gastrointestinal traits

**Figure 13.** Genetic correlation between ET and Pharmacological treatment

**Figure 14.** Genetic correlation between ET and other complex traits

**Figure 15.** Genetic correlation between ET and brain volumes

**Figure 16.** Spatial transcriptomic maps of *RASGRP1, NRGN, CALM2, CHN1, SEINC1, PPP3R1, GPM6A,* and *CALM1* genes involved in excitatory neuronal signaling within the hippocampal CA region.

**Figure 17.** Spatial transcriptomic maps of *CLSTN1, C9orf16, FBXW7, CYFIP2* and *SLC17A7* genes involved in excitatory neuronal signaling within the LA region.

**Figure 18.** Spatial transcriptomic maps of *PRKCB, SYT1, SNAP25, SLC24A2, ATP2B1, DNM1, VSNL1, NPTN, MAPK10, OSBPL1A, R3HDM1, DCLK1, ATP2B2, KALRN, NDRG3, SV2B, GRIA2, HIVEP2, PHACTR1, PCSK2, HOMER1, GRIN1, CNKSR2, PLCB1,* and *BCL11A* genes involved in excitatory neuronal signaling within the L5/6 region.

**Figure 19.** Spatial transcriptomic maps of *ATP2A2* and *VAMP2* genes involved in excitatory neuronal signaling within the L6 region.

**Figure 20.** Spatial transcriptomic maps of *PPP3CA* genes involved in excitatory neuronal signaling within the GN DG region.

**Figure 21.** Spatial transcriptomic maps of *SCN1B, RAB6A, NME7, NDRG4* and *ATP1B1* genes involved in excitatory neuronal signaling within the IN Pvalb+ region.

**Figure 22.** Spatial transcriptomic maps of *MDH1, SNRPN* and *SCG5* genes involved in interneurons Pvalb+Gad1+.

**Figure 23.** Spatial transcriptomic maps of *OPCML* gene involved in OPC region.

**Figure 24.** Gene Spatial Specificity (GSS) scores of *RASGRP1, NRGN, CALM2, CHN1, SEINC1, PPP3R1, GPM6A,* and *CALM1* in excitatory neuronal signaling within the hippocampal CA region.

**Figure 25.** Gene Spatial Specificity (GSS) scores of *CLSTN1, C9orf16, FBXW7, CYFIP2* and *SLC17A7* in excitatory neuronal signaling within LA region.

**Figure 26.** Gene Spatial Specificity (GSS) scores of *PRKCB, SYT1, SNAP25, SLC24A2, ATP2B1, DNM1, VSNL1, NPTN, MAPK10, OSBPL1A, R3HDM1, DCLK1, ATP2B2, KALRN, NDRG3, SV2B, GRIA2, HIVEP2, PHACTR1, PCSK2, HOMER1, GRIN1, CNKSR2, PLCB1,* and *BCL11A* in excitatory neuronal signaling within L5/6 region.

**Figure 27.** Gene Spatial Specificity (GSS) scores of *ATP2A2* and *VAMP2* in excitatory neuronal signaling within L6 region.

**Figure 28.** Gene Spatial Specificity (GSS) scores of *PPP3CA* in excitatory neuronal signaling within GN DG region.

**Figure 29.** Gene Spatial Specificity (GSS) scores of *PSCN1B, RAB6A, NME7, NDRG4 and ATP1B1* in excitatory neuronal signaling within IN Pvalb+ region.

**Figure 30.** Gene Spatial Specificity (GSS) scores of *MDH1, SNRPN* and *SCG5* in interneurons Pvalb+Gad1+.

**Figure 31.** Gene Spatial Specificity (GSS) scores of the *OPCML* gene in OPC region.

**Figure 32.** Difference in mean age at diagnosis for essential tremor across PGS deciles

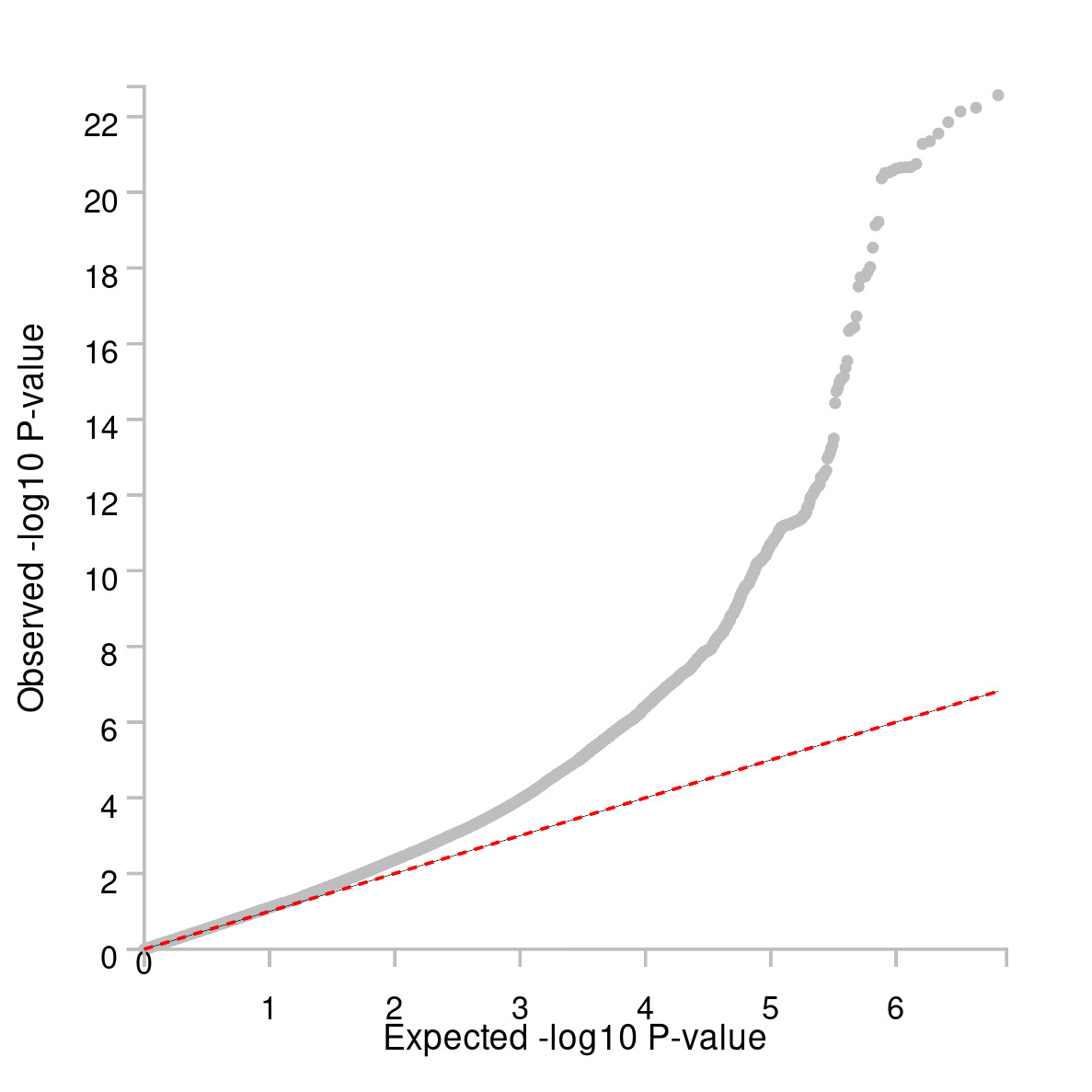

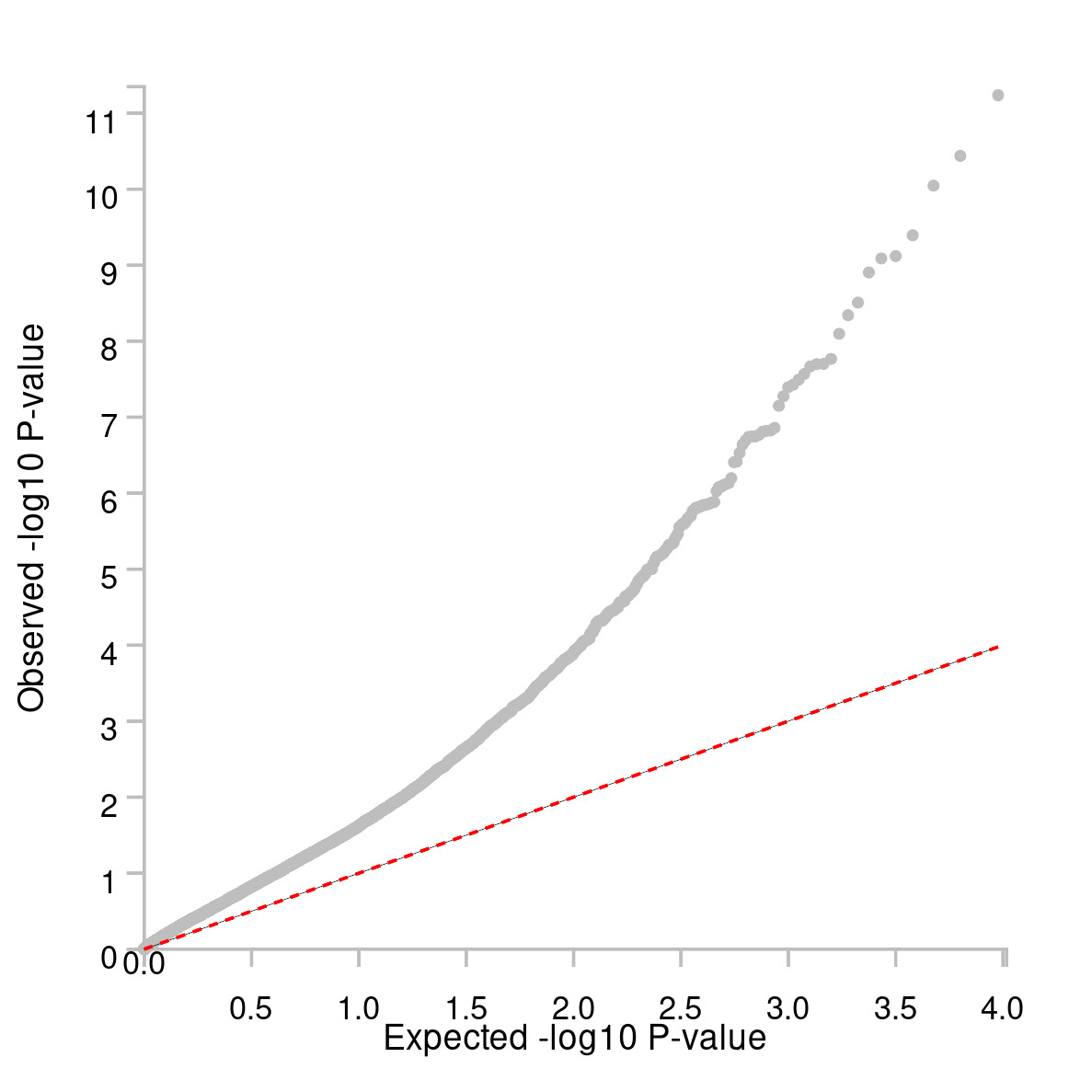

**Supplementary Figure 1. QQ plot of GWAS meta-analysis (a) and ET gene-level association study (b)**

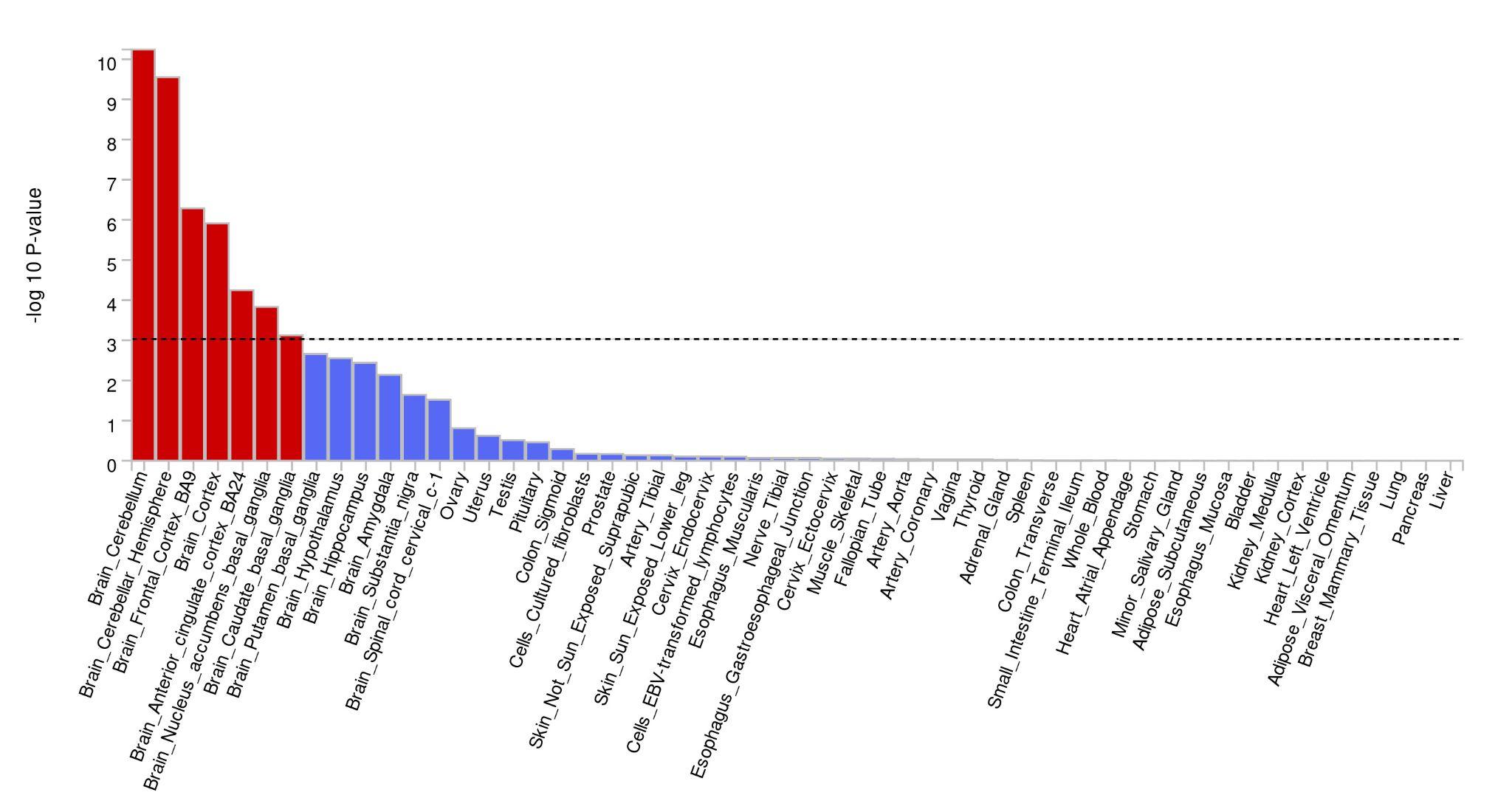

**Supplementary Figure 2. Gene enrichment across GTEx 53 samples**

**a**

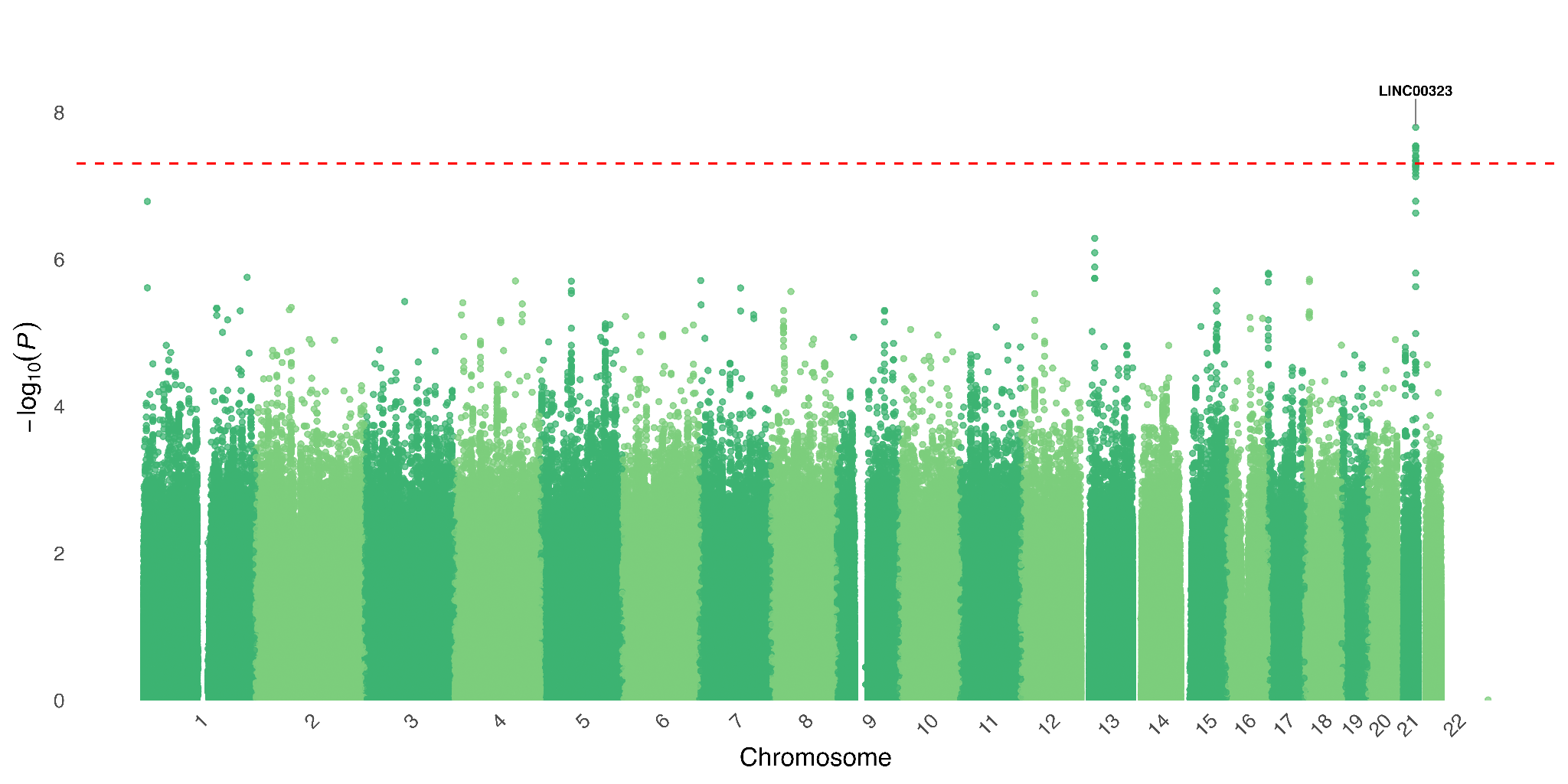

**b**

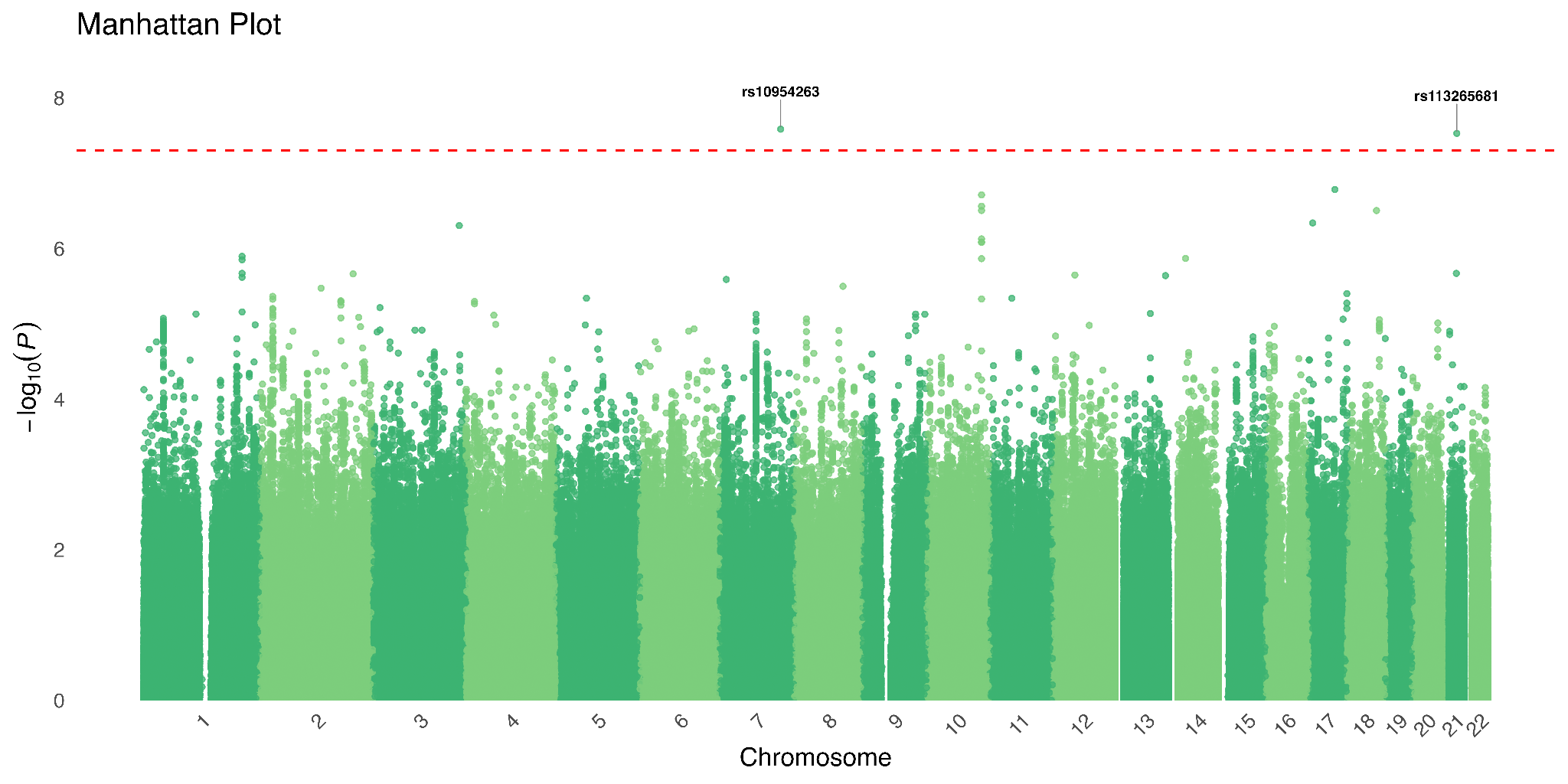

**c**

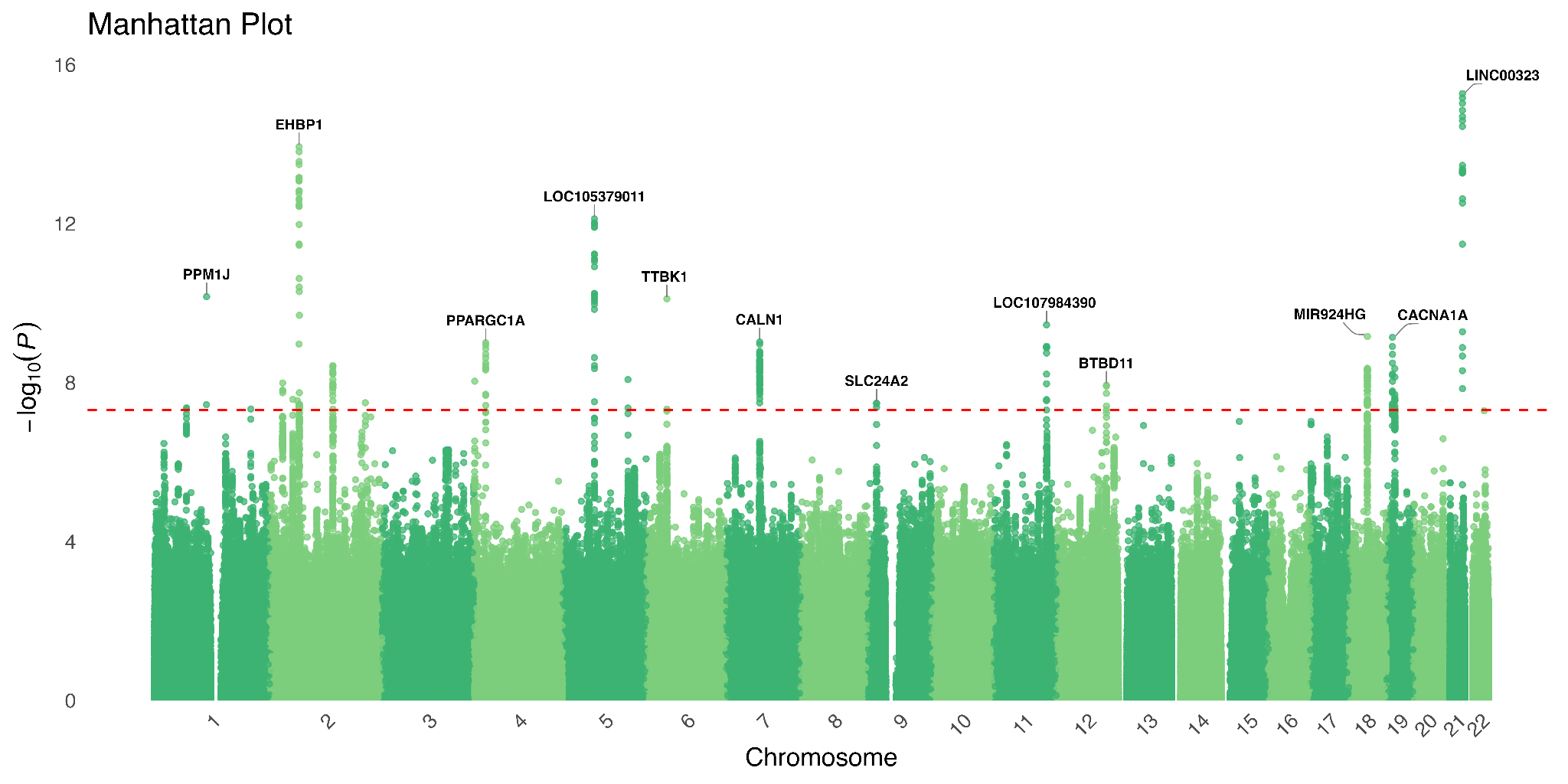

**Supplementary Figure 3. Manhattan plot of individual dataset.**

Manhattan plots showing GWAS results under an additive model for a) 23andMe, b) All of Us and c) MVP. The -log_10_*P*-values (y-axis) are plotted for each variant against their chromosomal position (x-axis). *P*-values are two-sided and derived from a likelihood-ratio test.

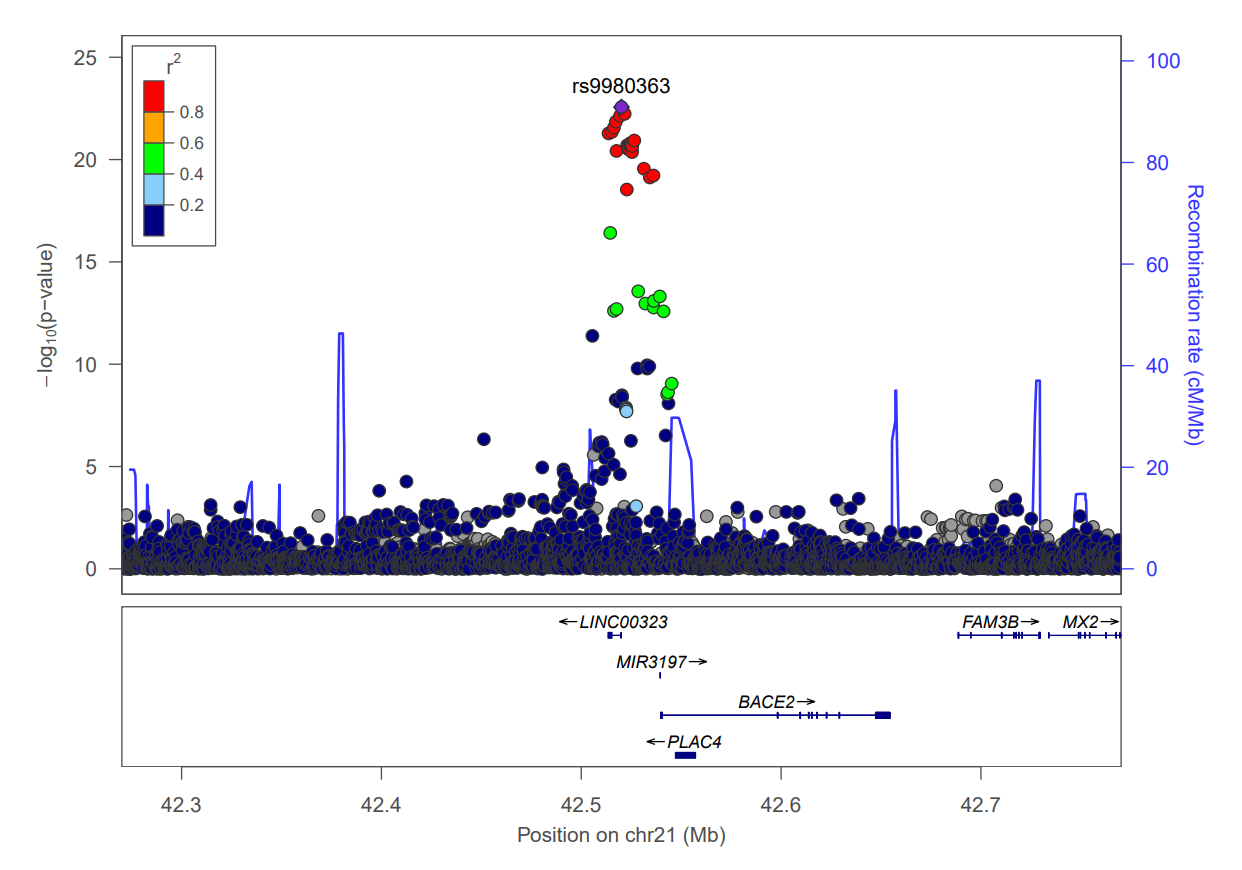

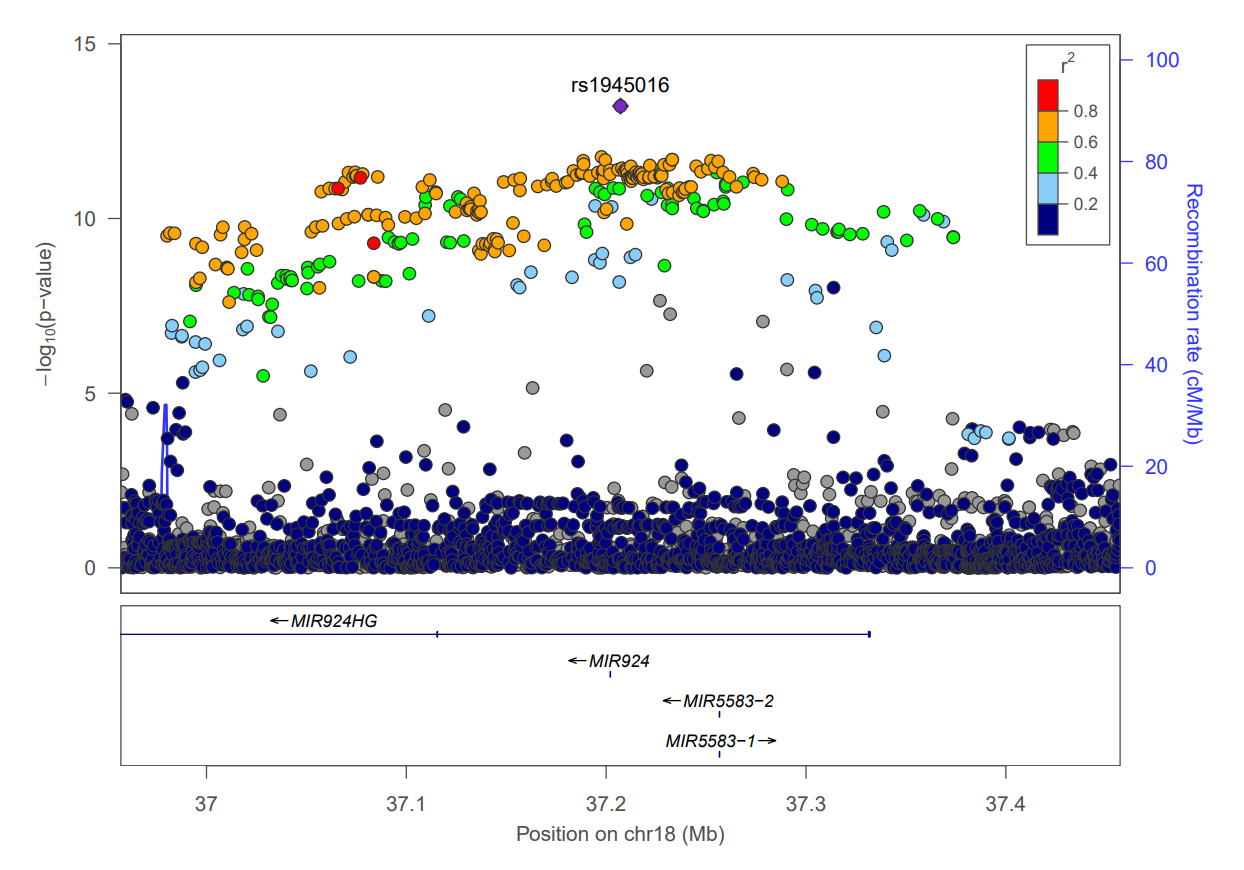

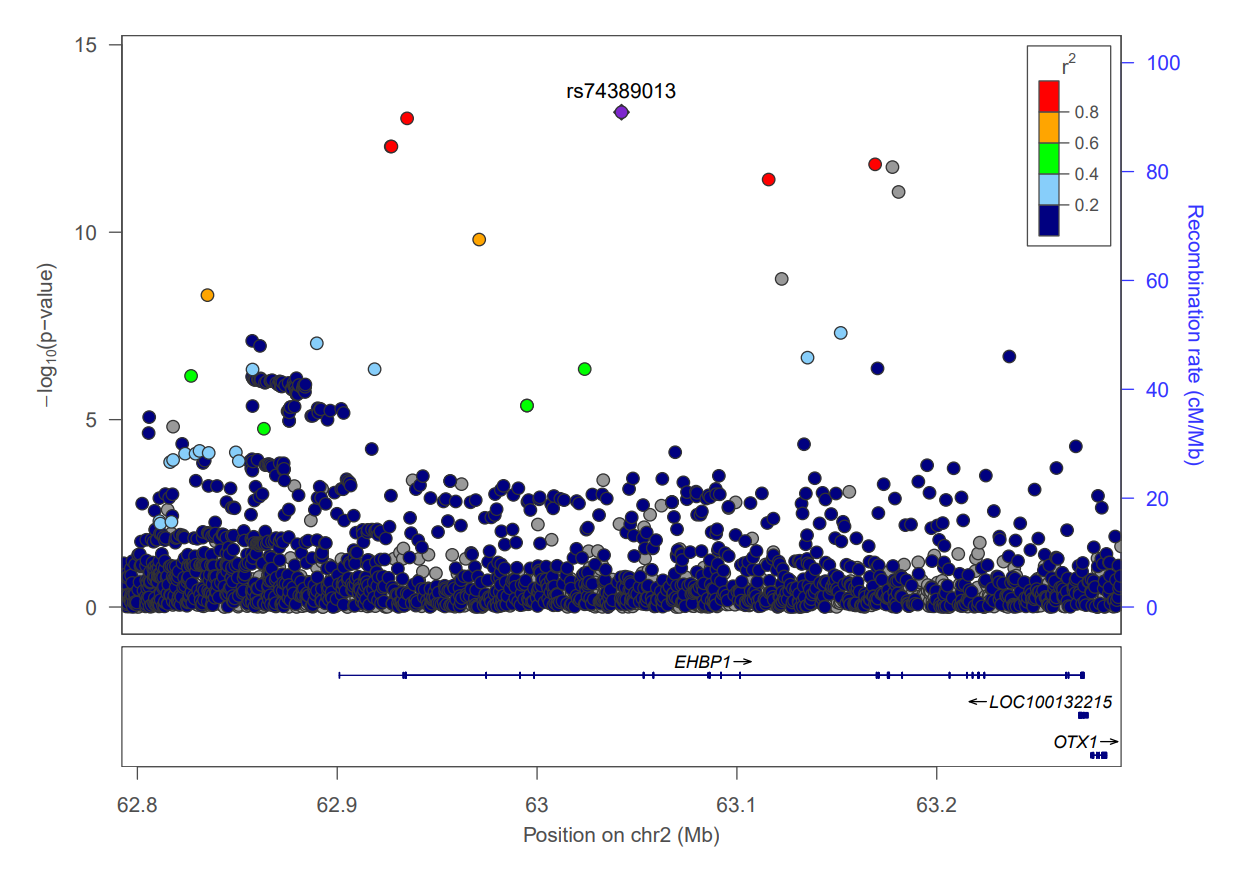

**Supplementary Figure 4. Regional association plots of loci associated with ET - Previously known associations with ET.** Each point represents a variant plotted by its chromosomal position (x-axis) and –log₁₀(P-value) (left y-axis), derived from a two-sided logistic regression likelihood-ratio test. Variants are colored by the degree of linkage disequilibrium (r²) with the lead variant (highlighted in red). Variants with predicted functional impact are indicated by their shape: squares represent moderate-impact variants, and diamonds represent high-impact variants. The recombination rate estimated from Icelandic data is shown as a solid blue line (right y-axis).

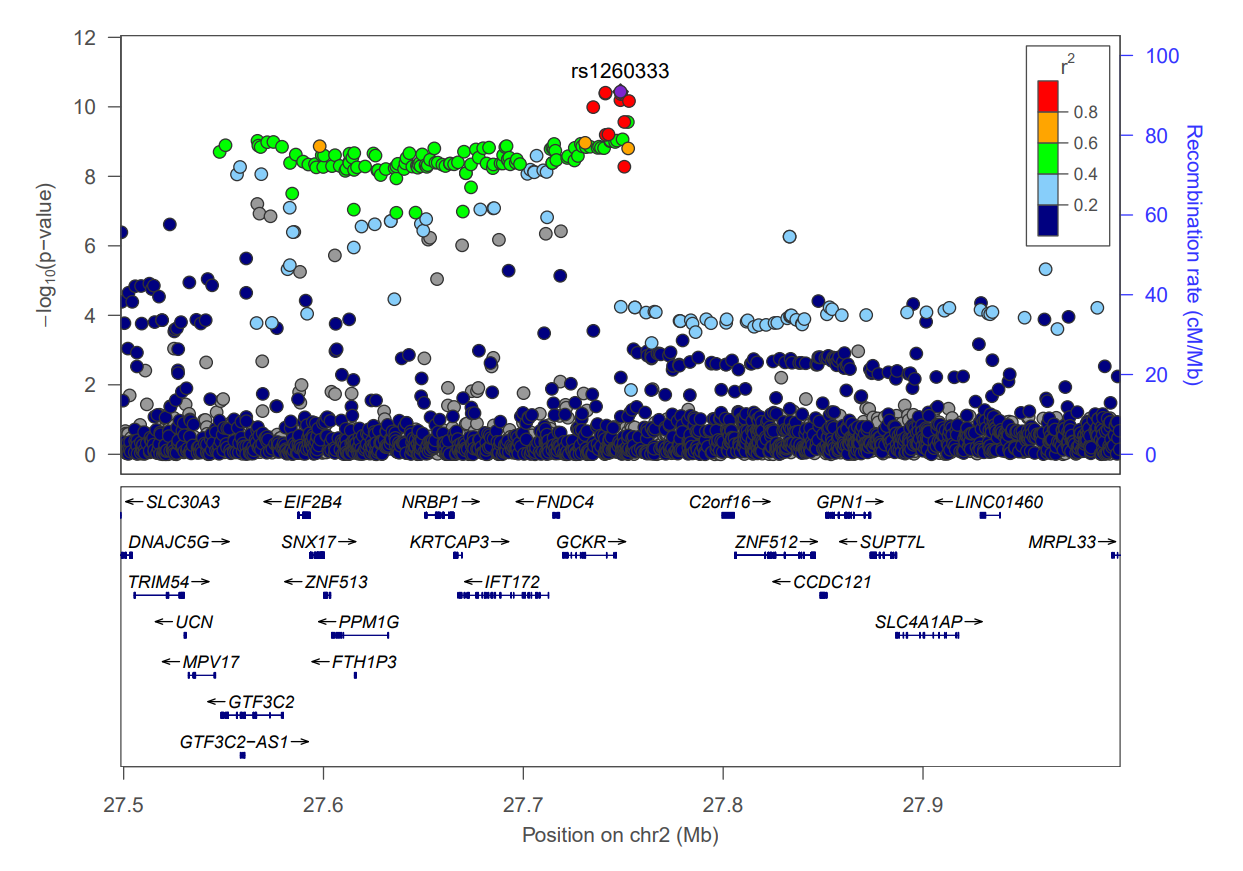

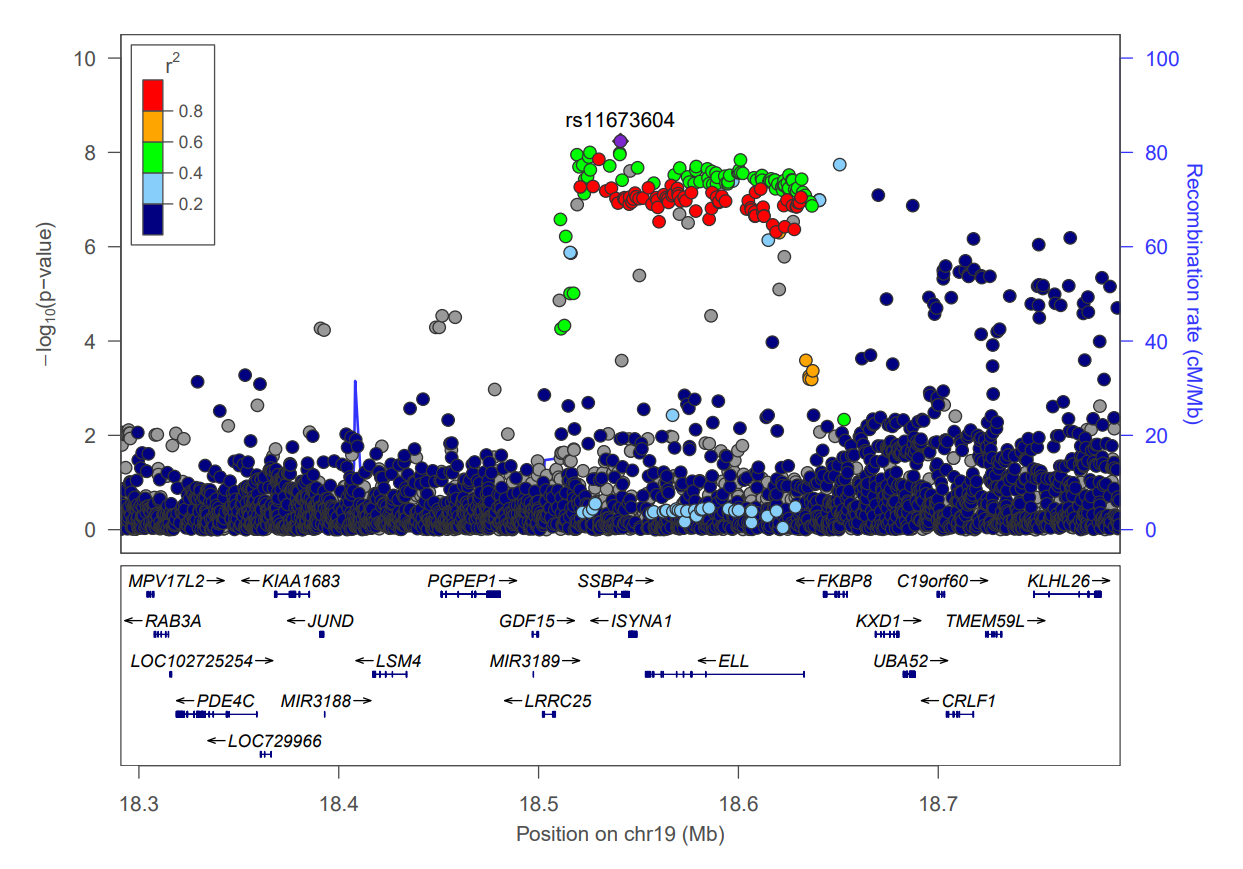

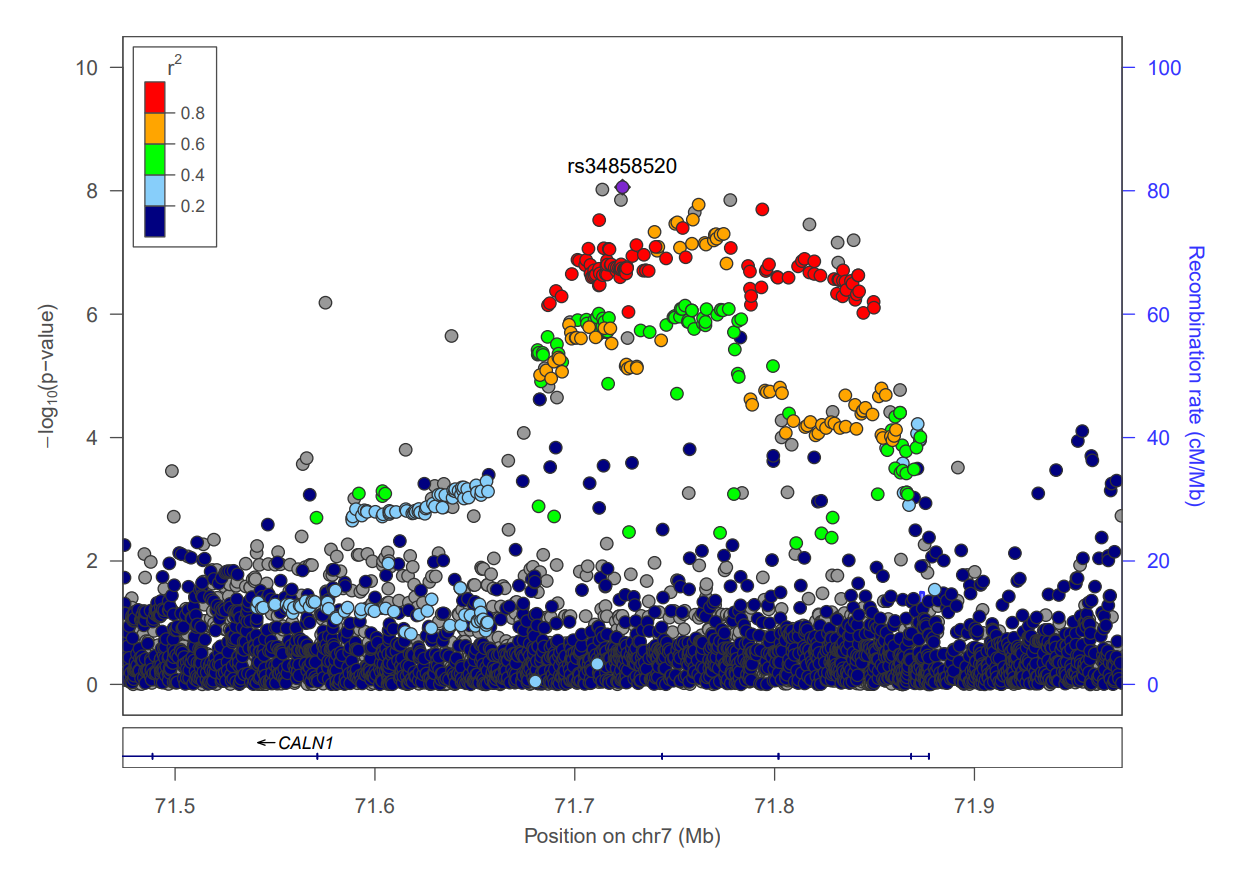

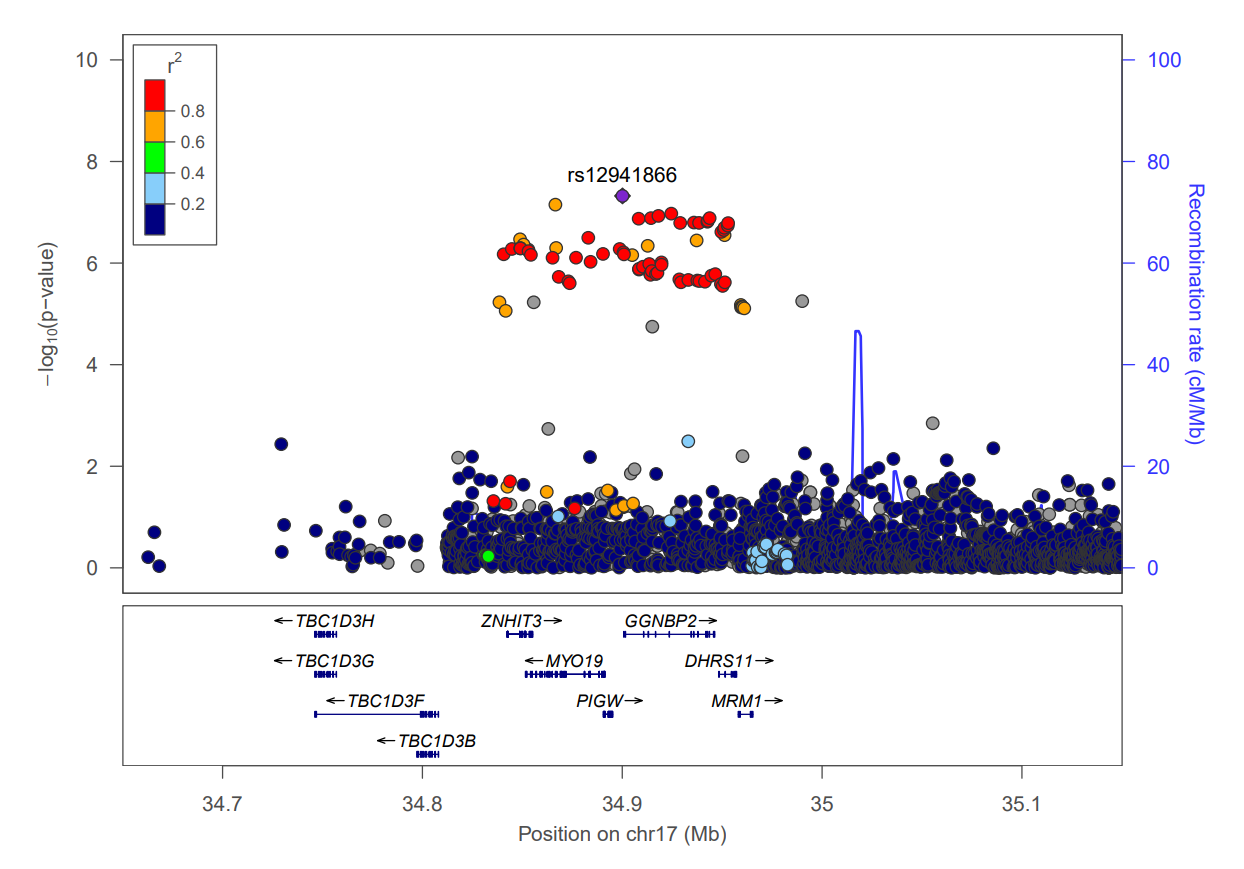

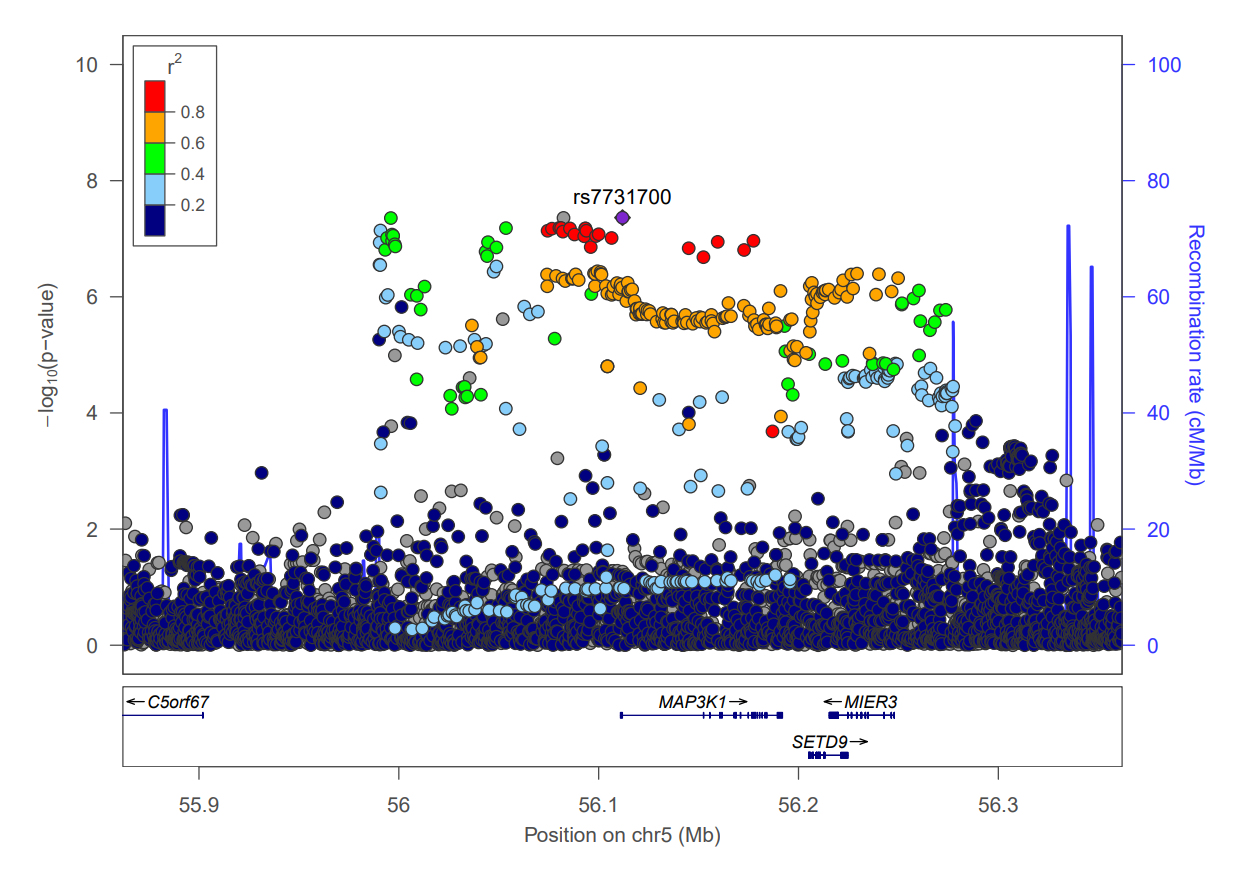

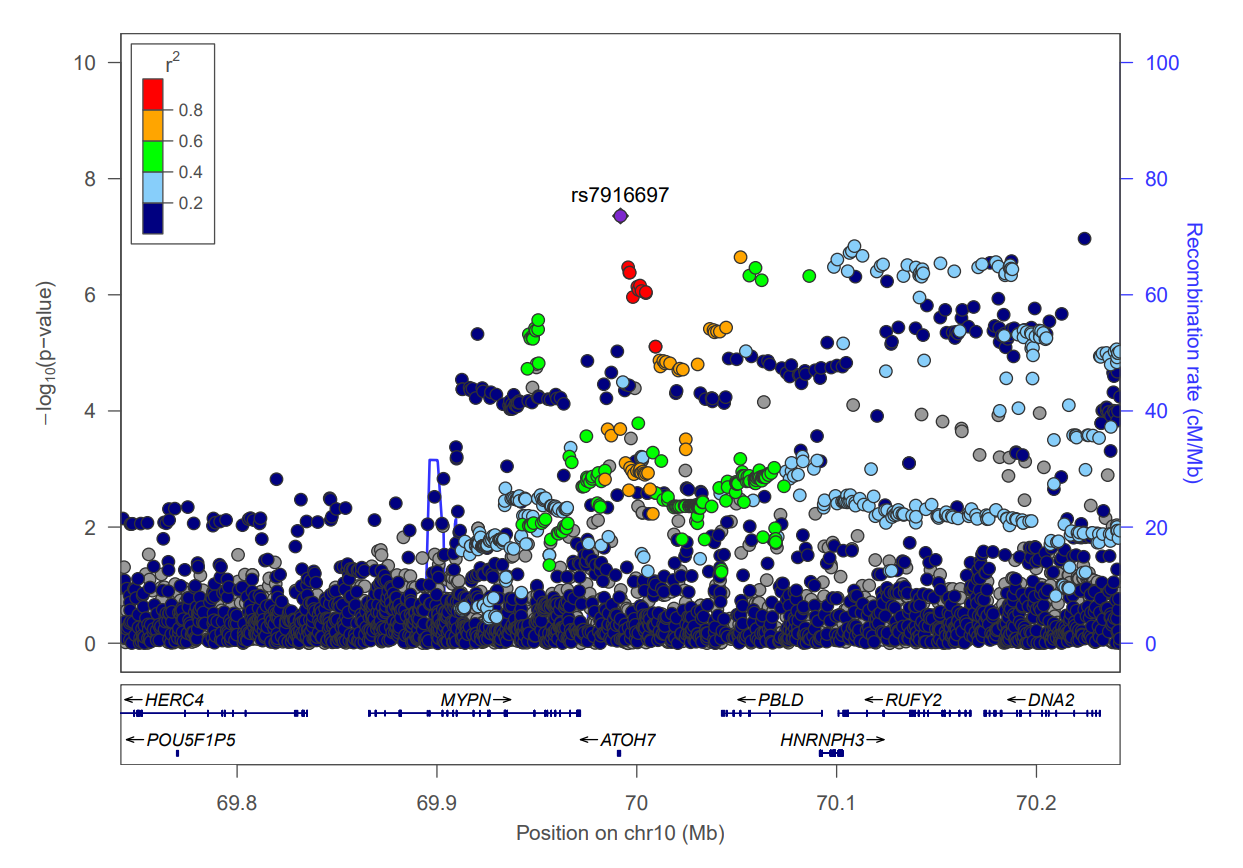

**Supplementary Figure 5. Regional plots of the loci associated with ET - Previously known associations.** Each point represents a variant plotted by its chromosomal position (x-axis) and –log₁₀(P-value) (left y-axis), derived from a two-sided logistic regression likelihood-ratio test. Variants are colored by the degree of linkage disequilibrium (r²) with the lead variant (highlighted in red). Variants with predicted functional impact are indicated by their shape: squares represent moderate-impact variants, and diamonds represent high-impact variants. The recombination rate estimated from Icelandic data is shown as a solid blue line (right y-axis).

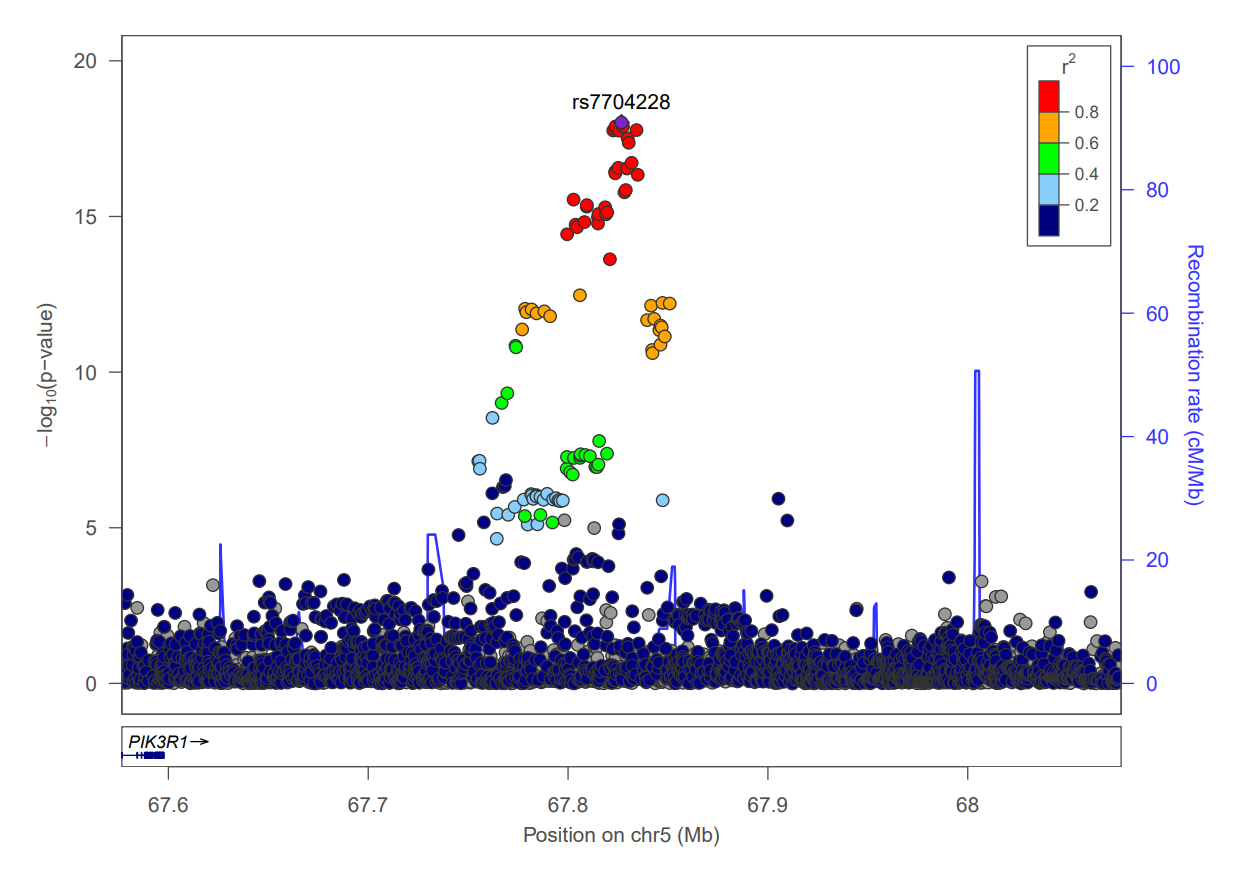

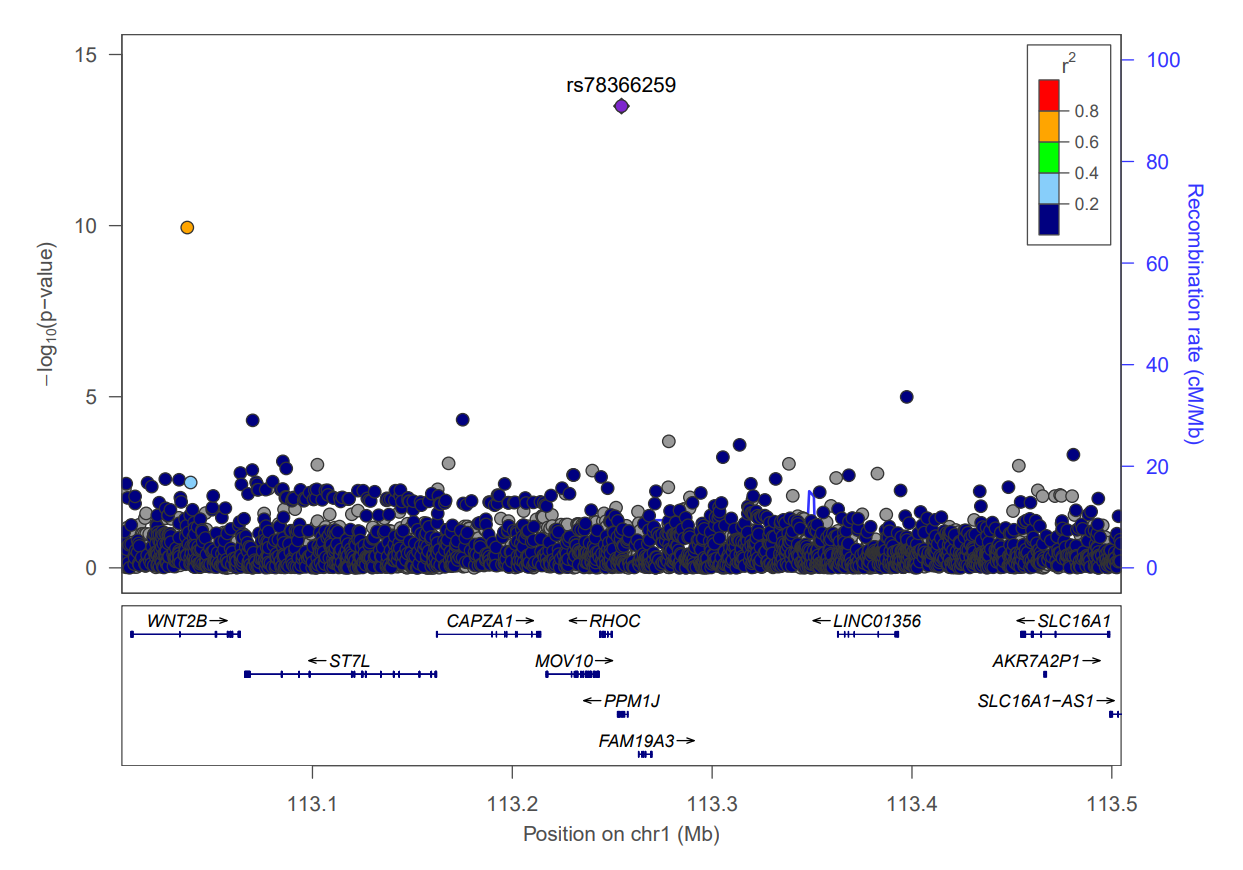

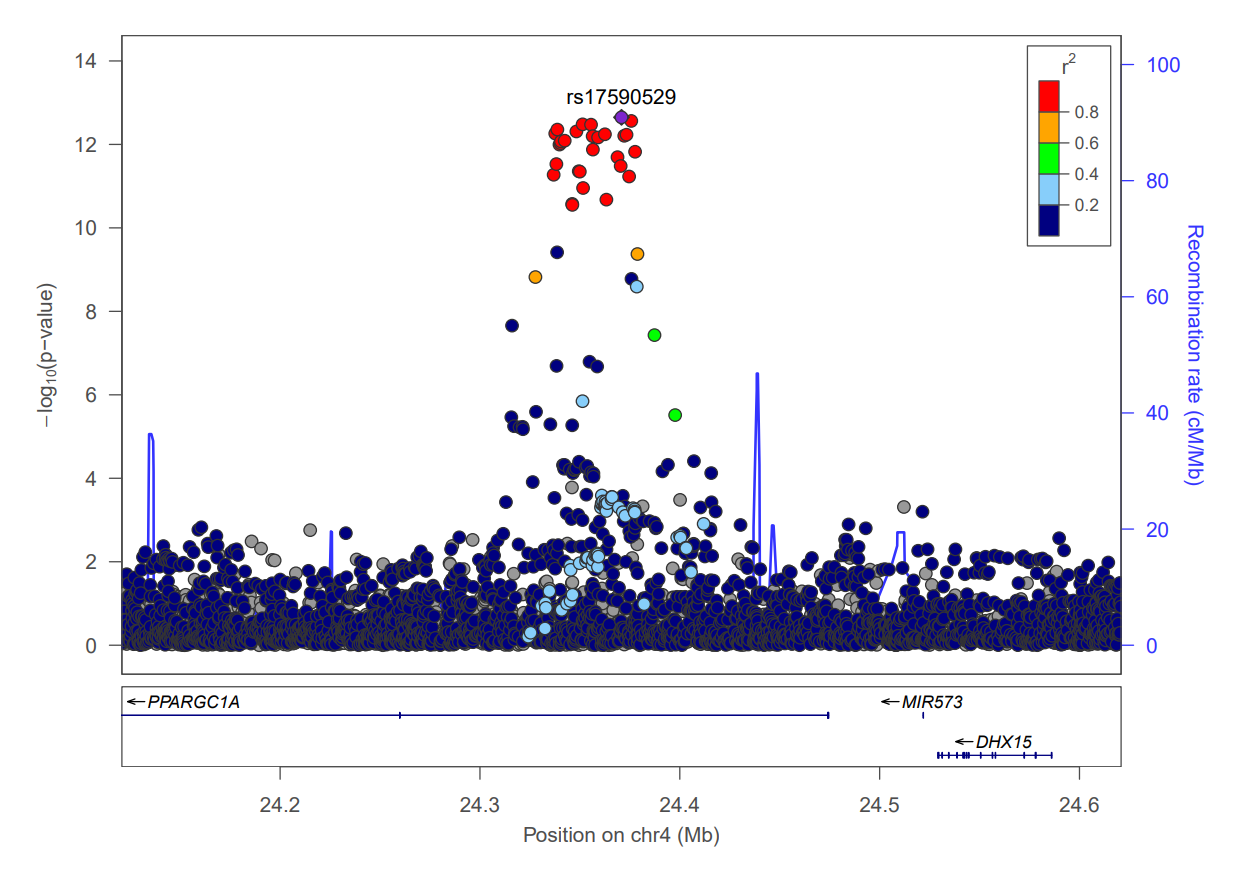

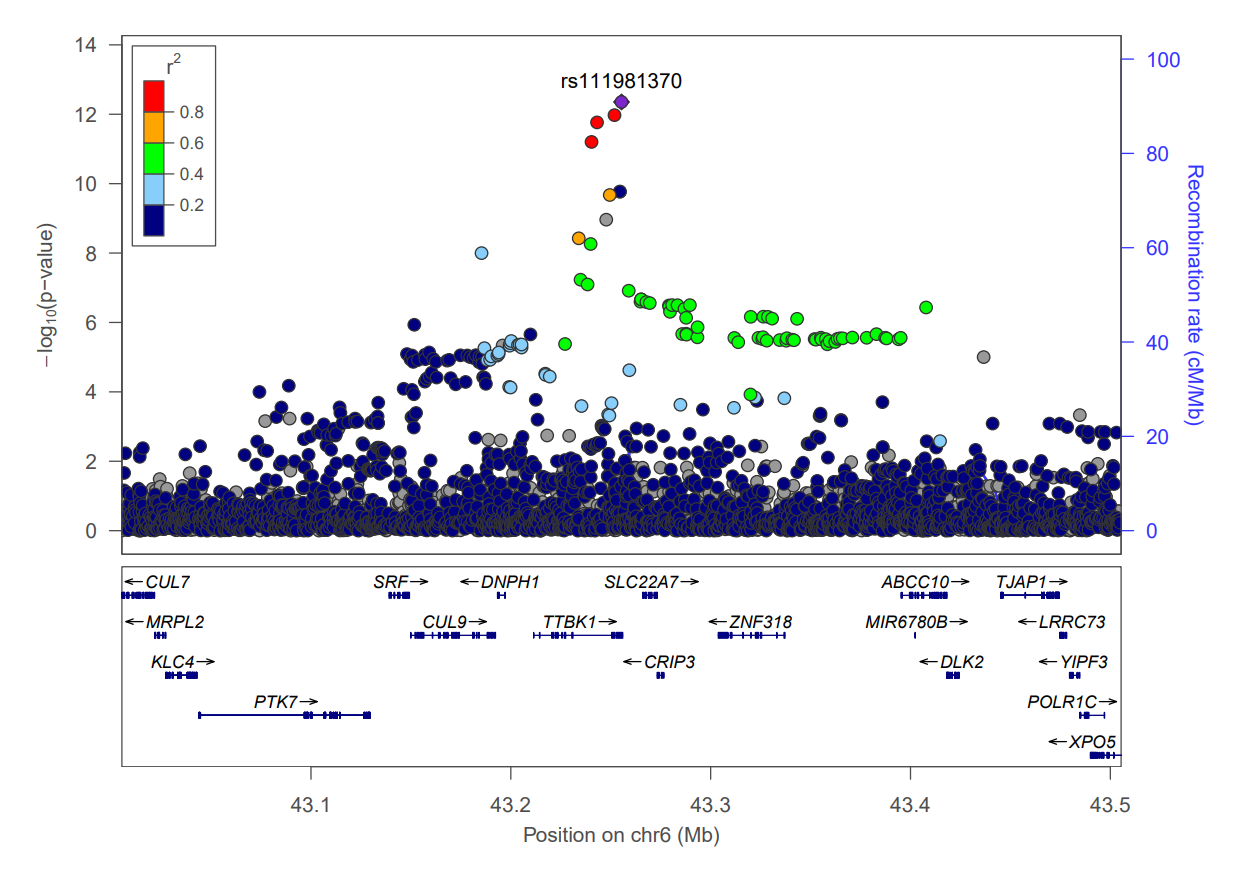

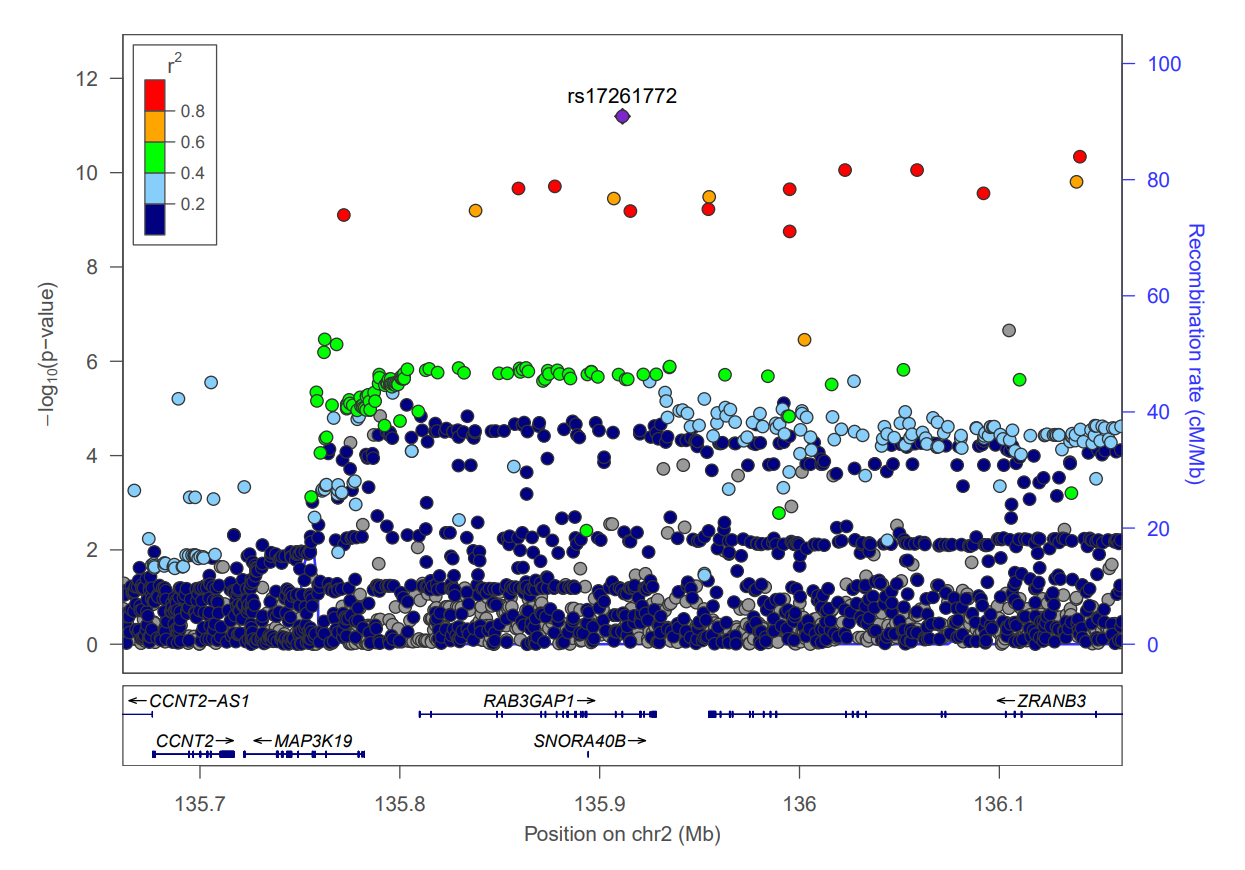

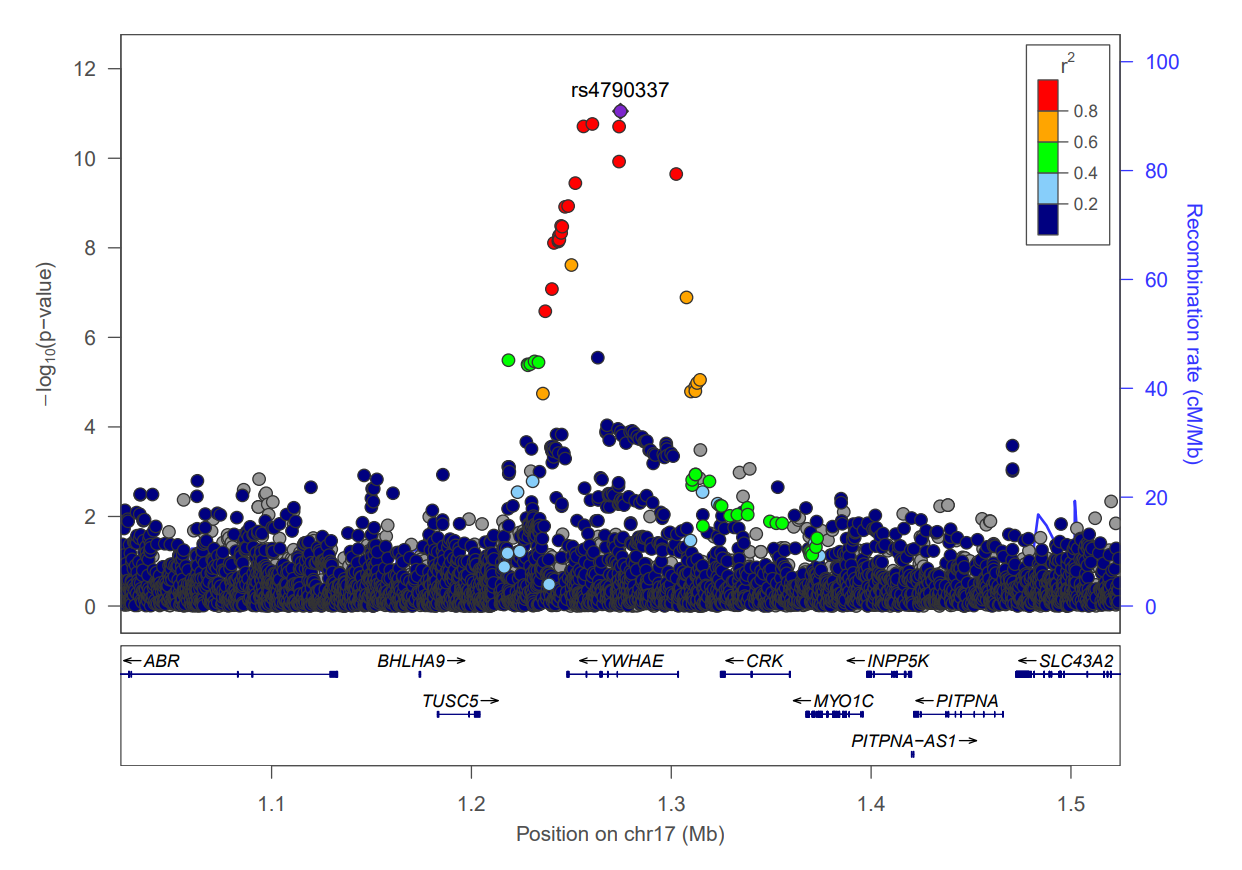

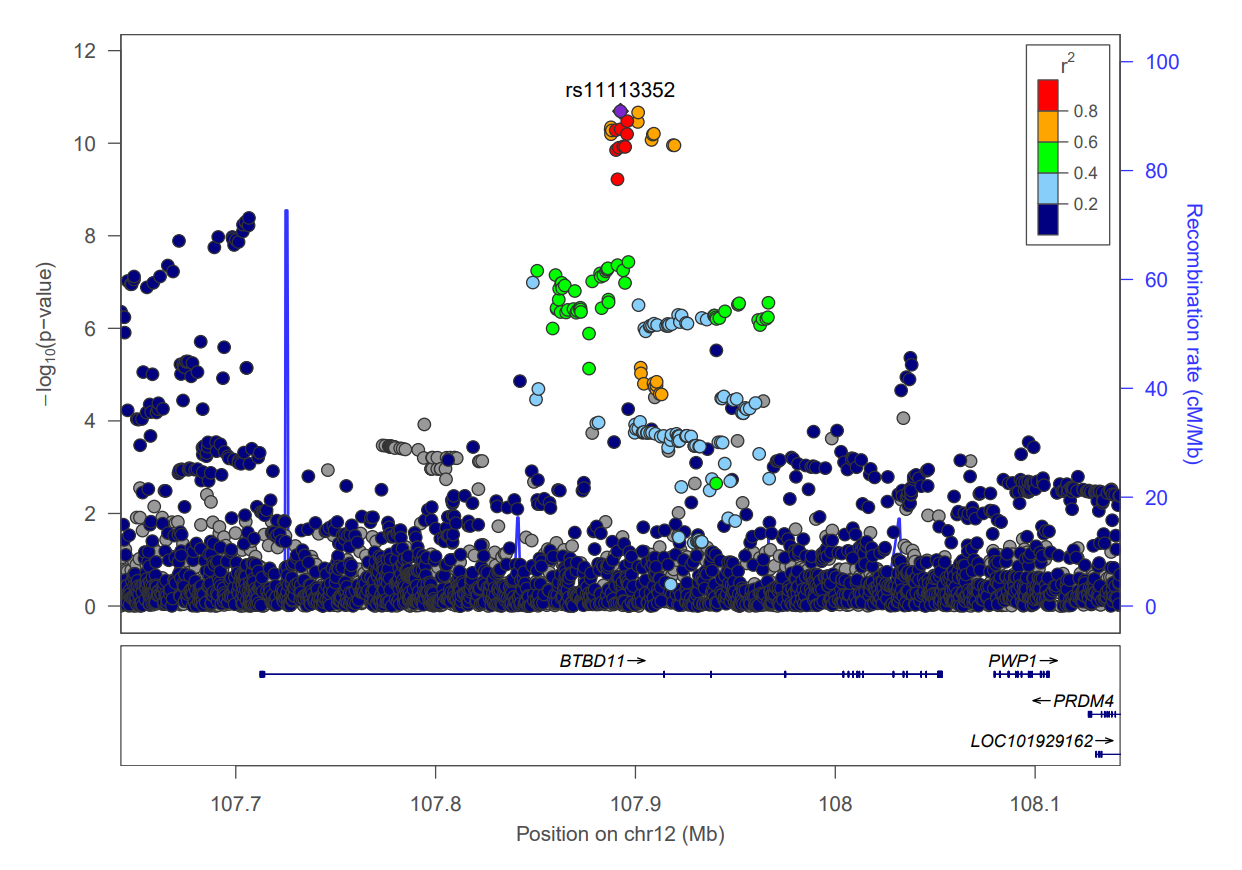

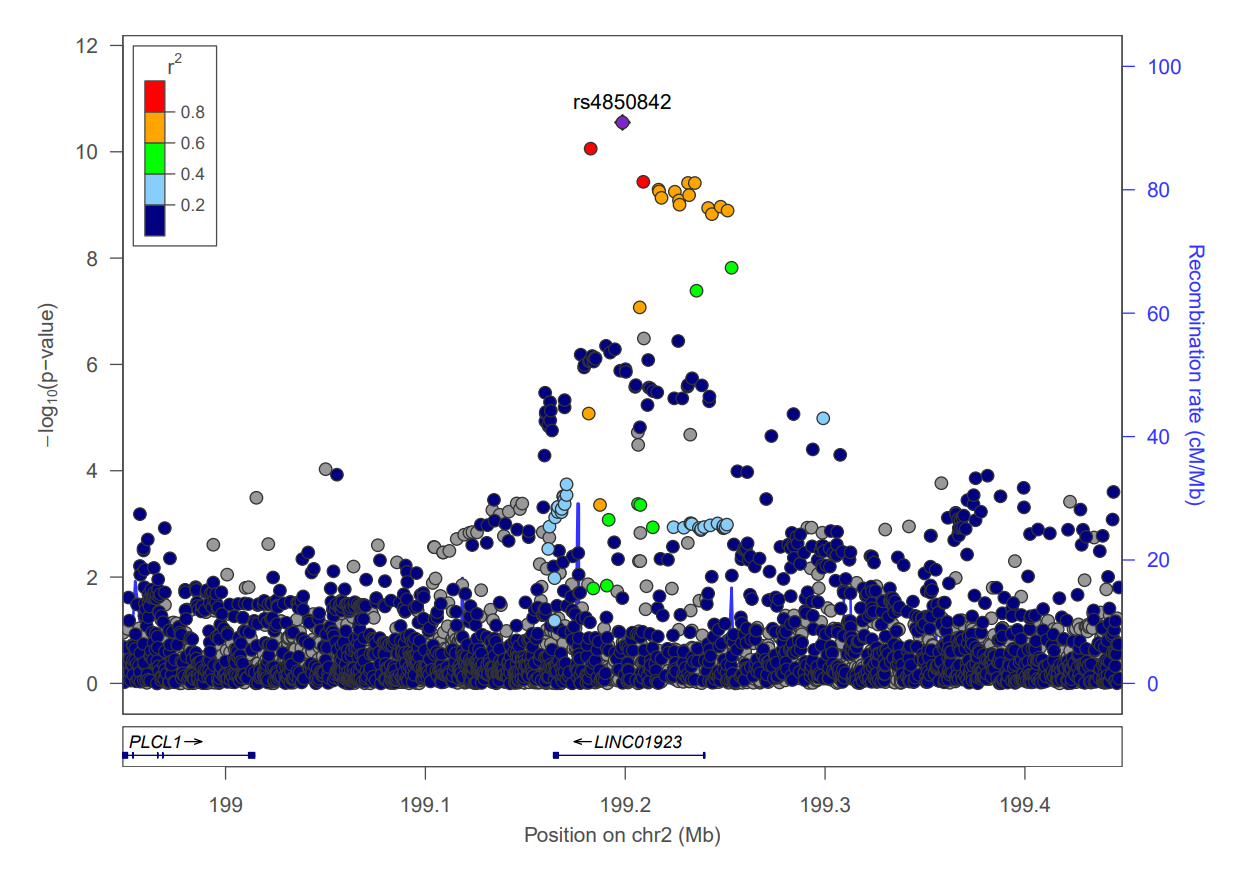

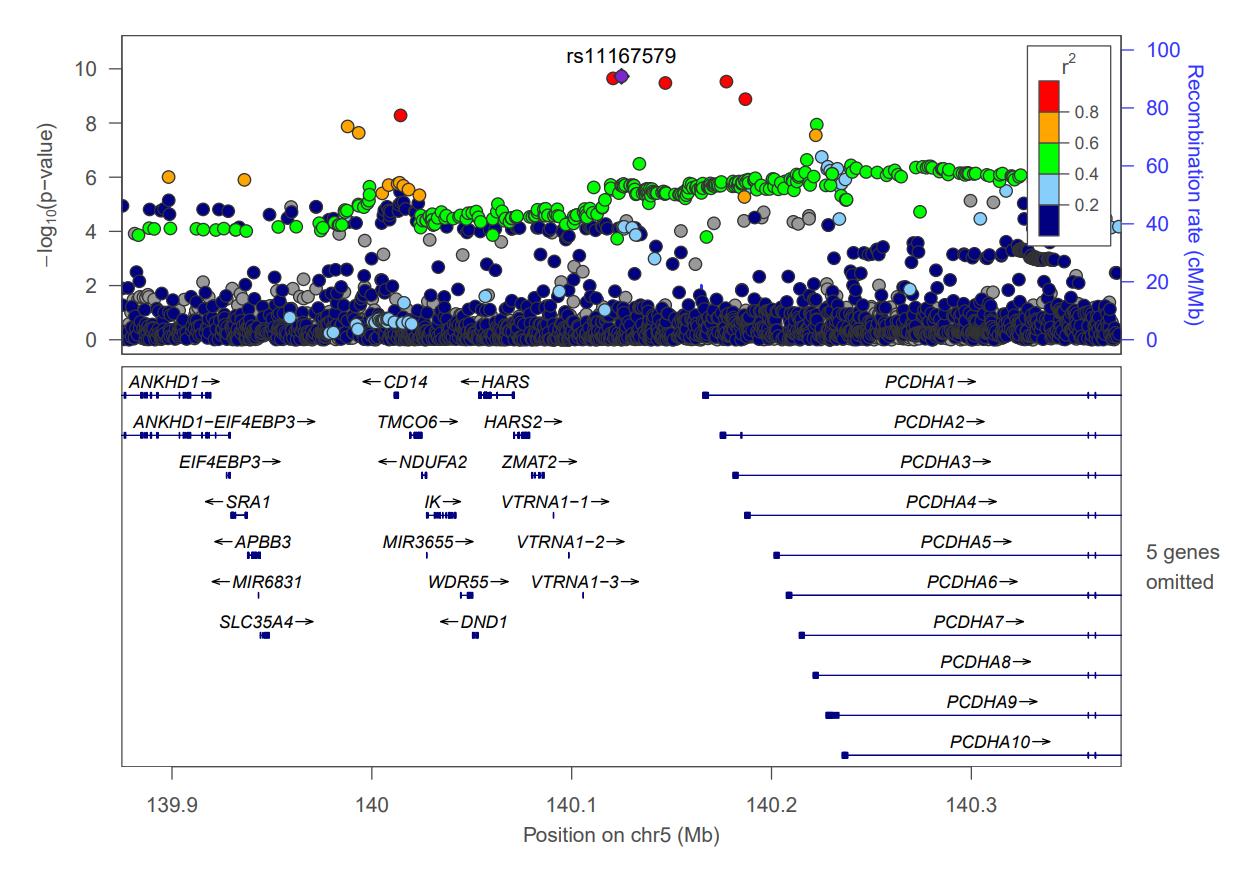

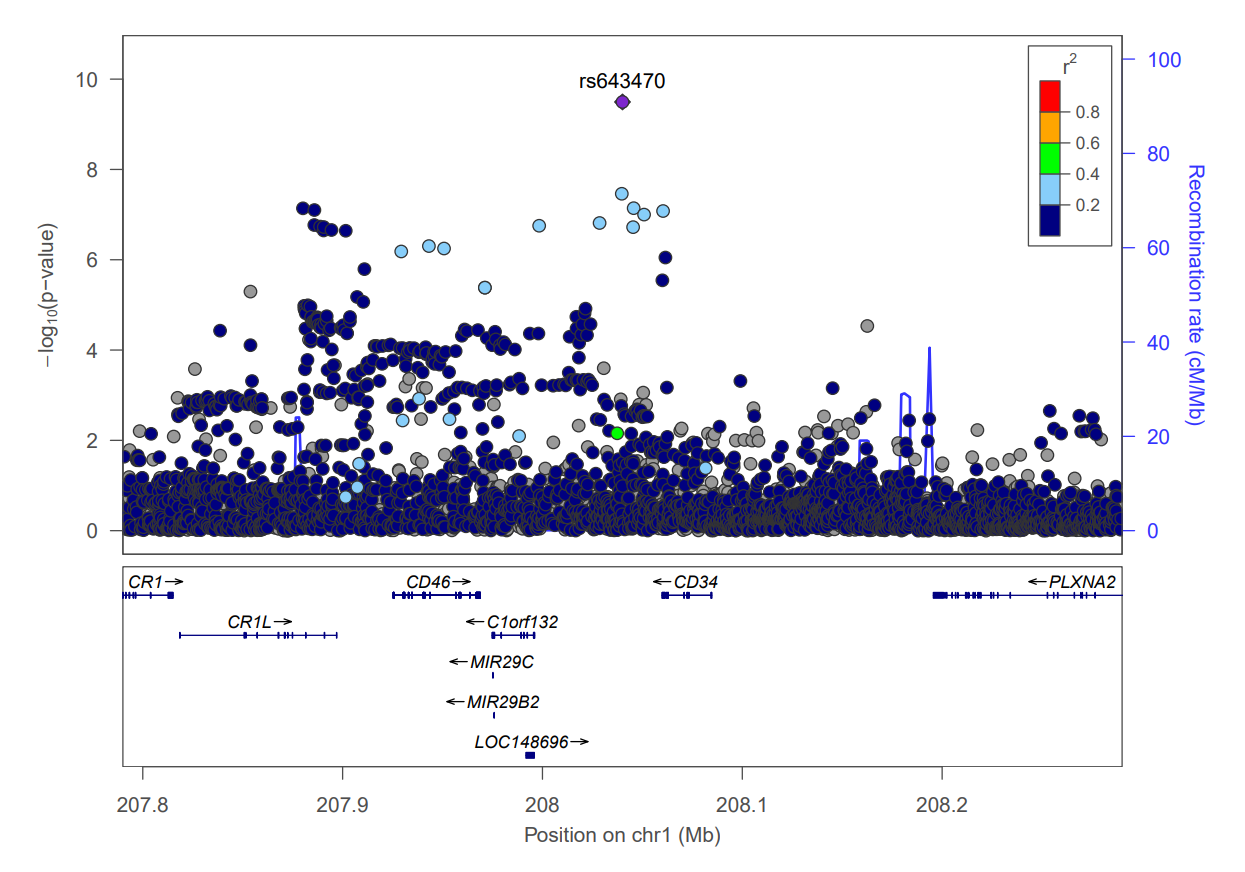

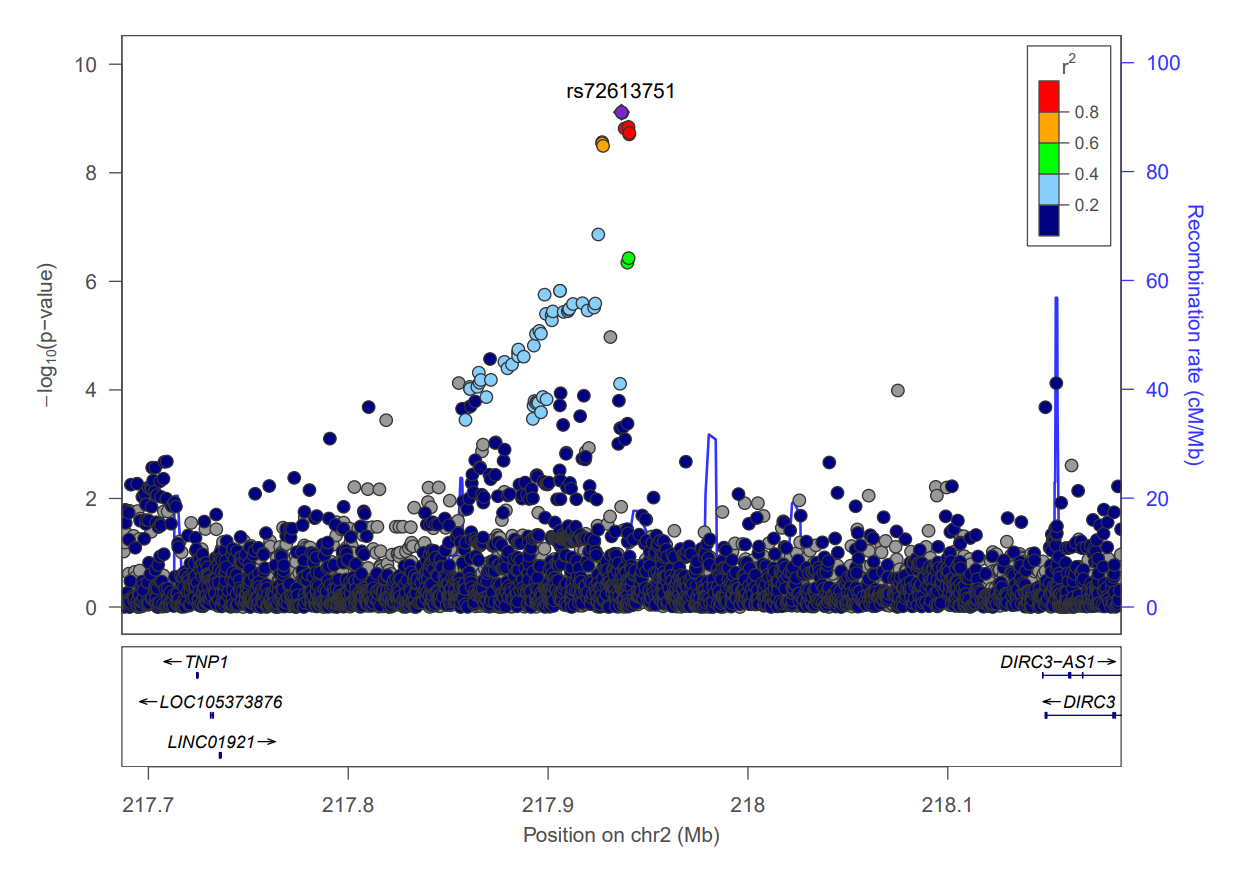

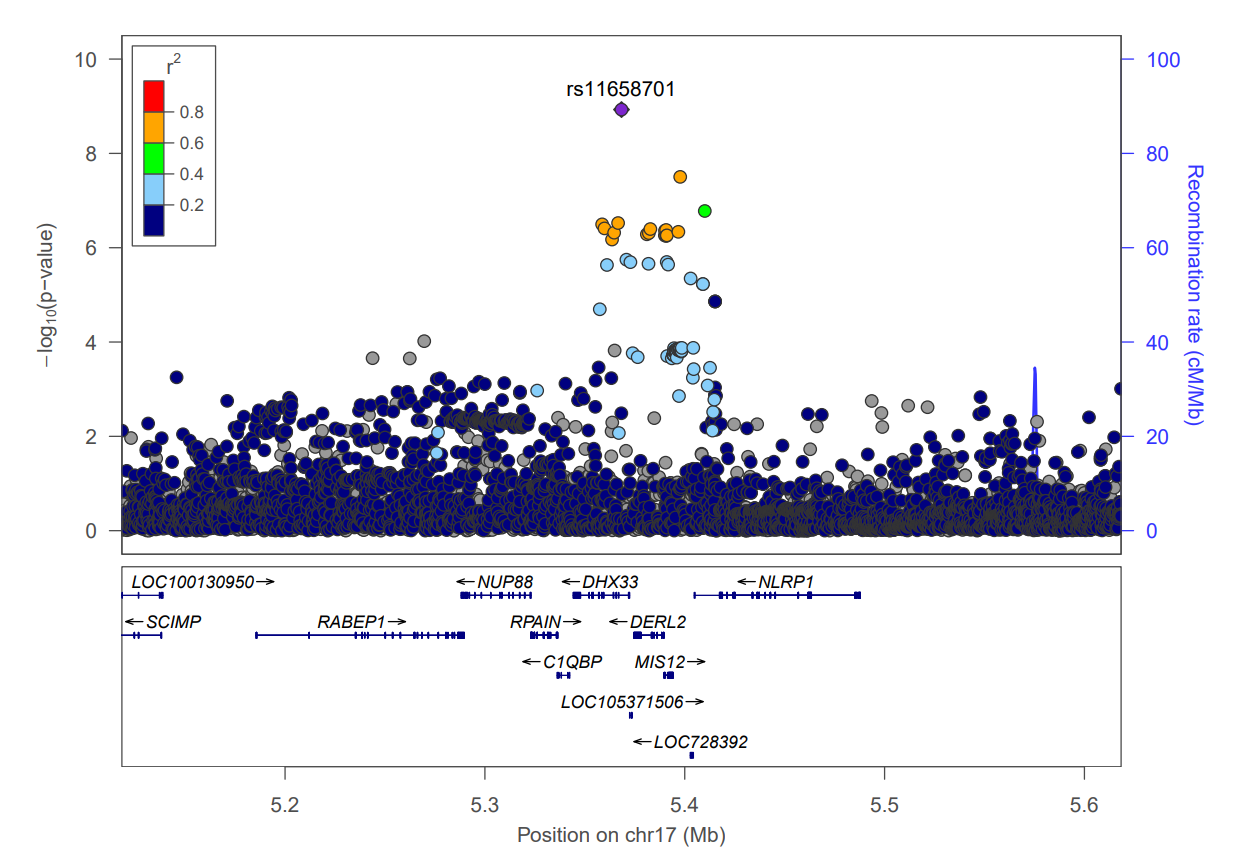

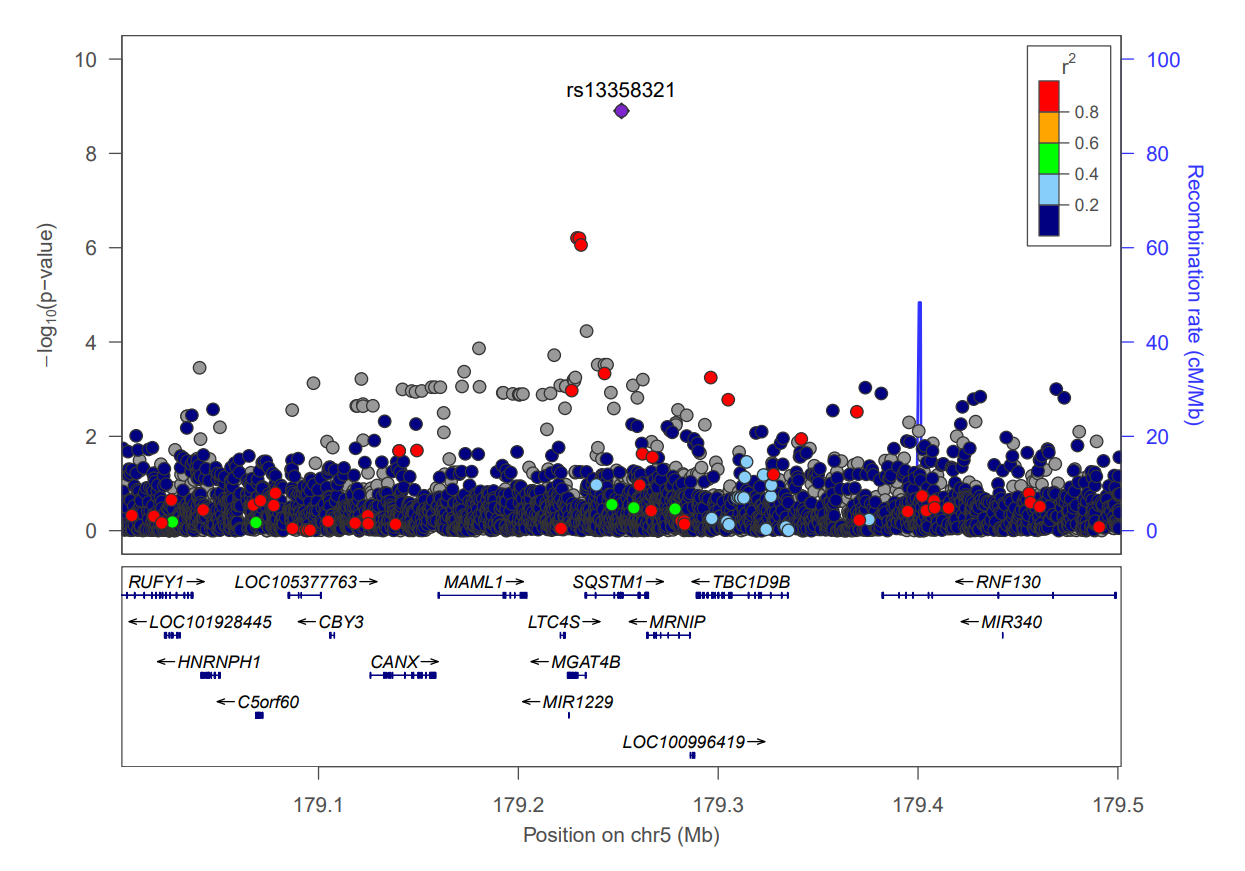

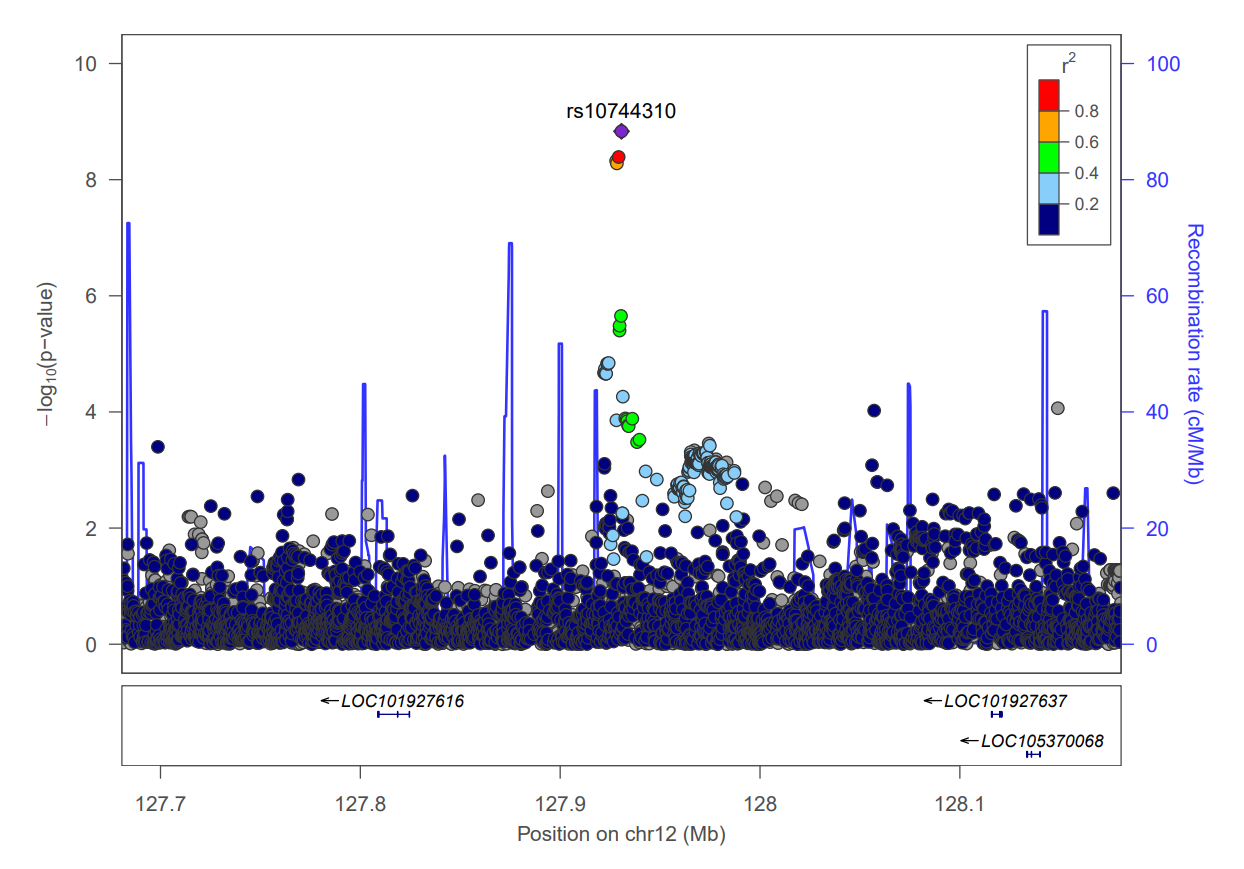

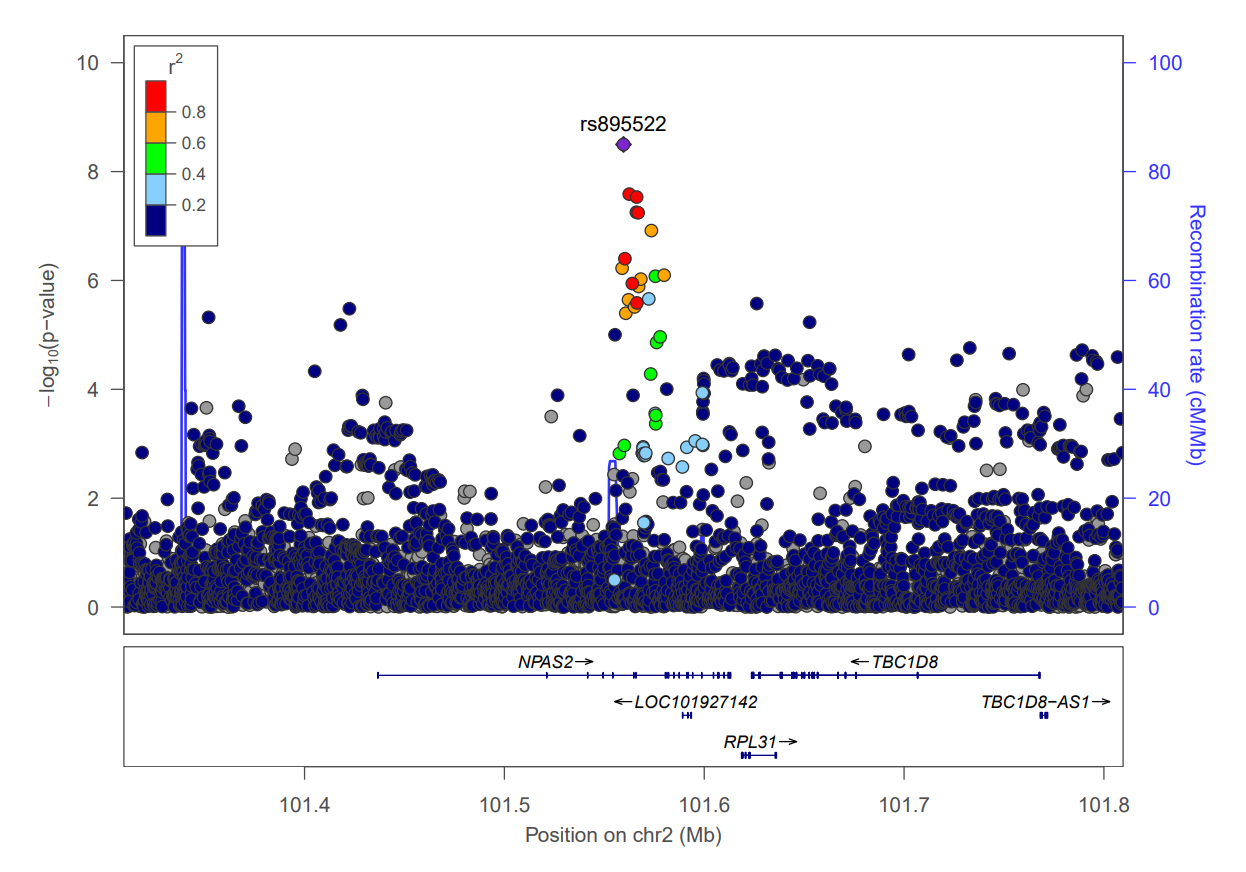

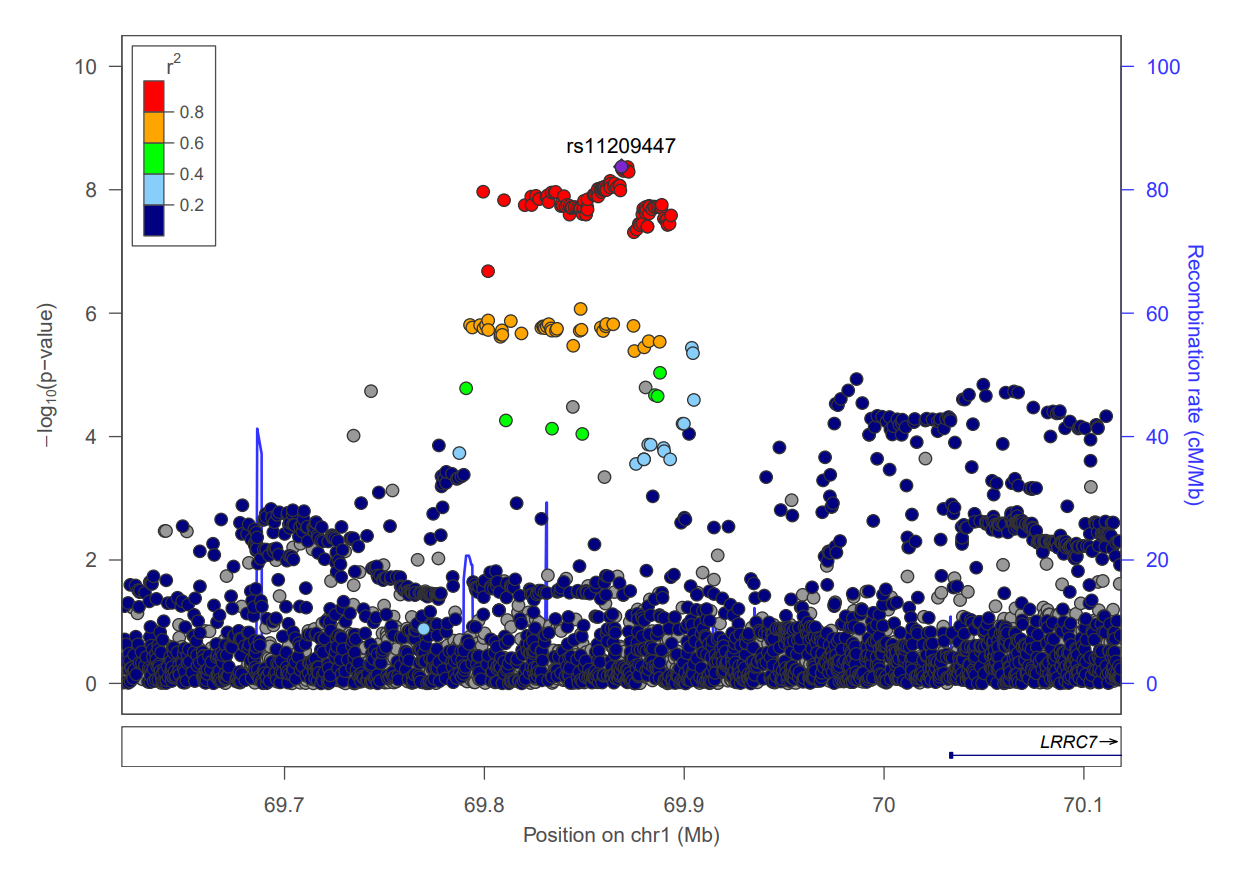

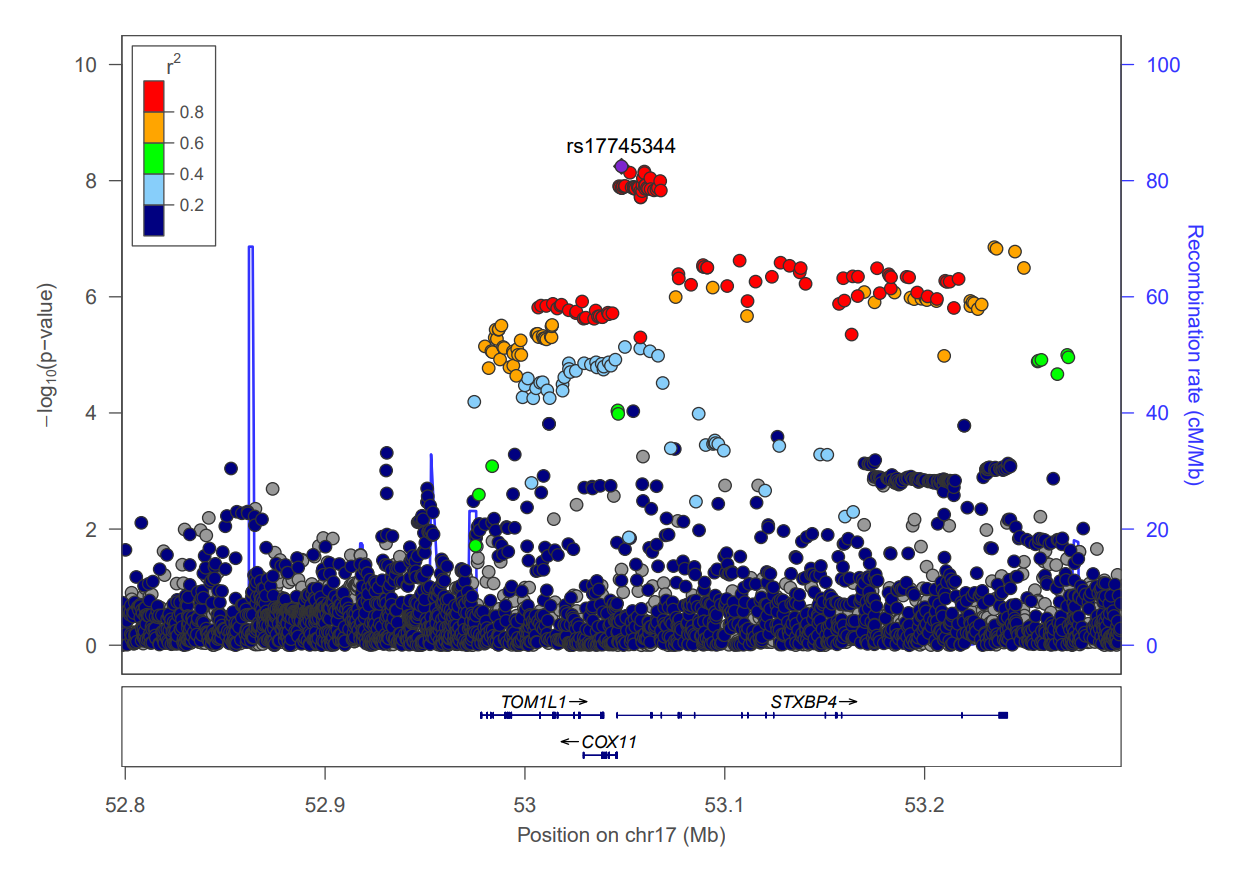

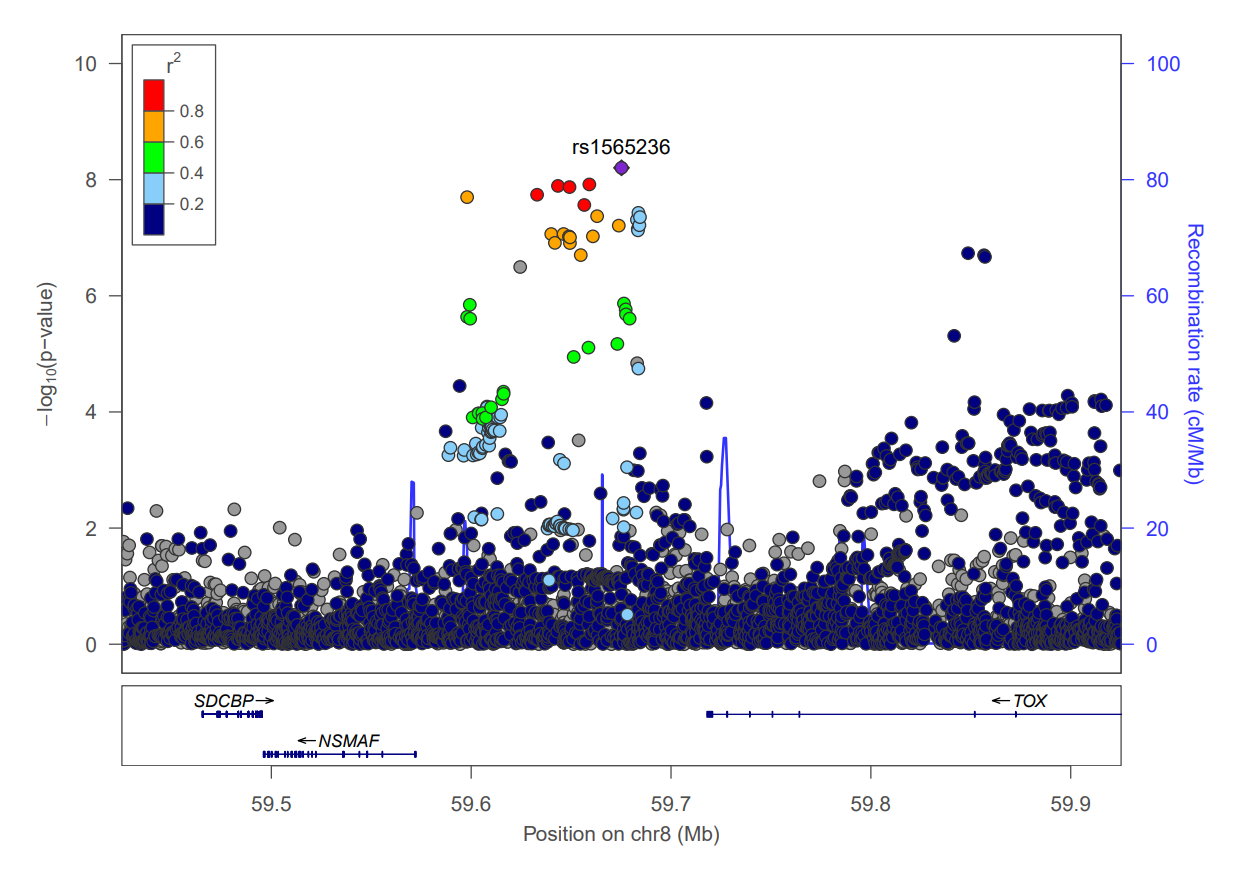

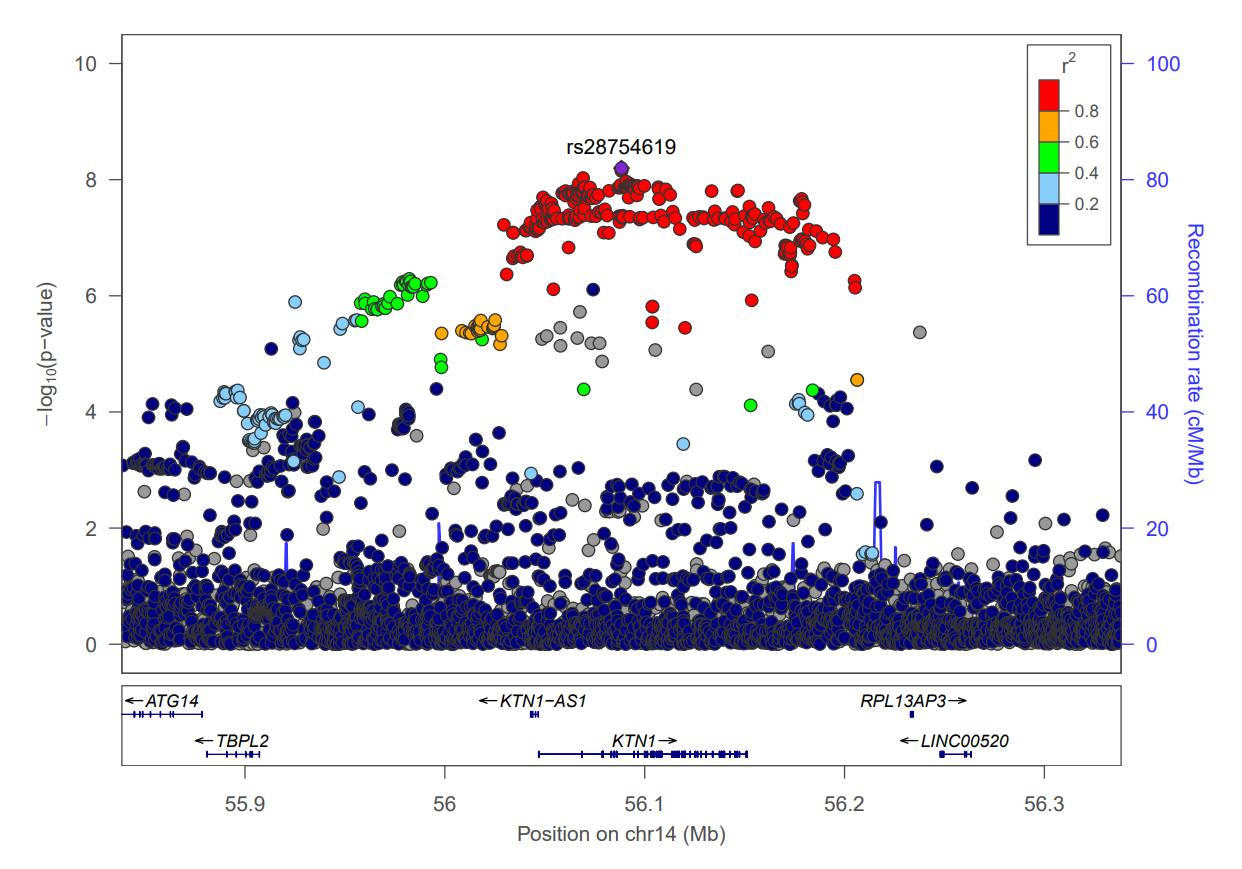

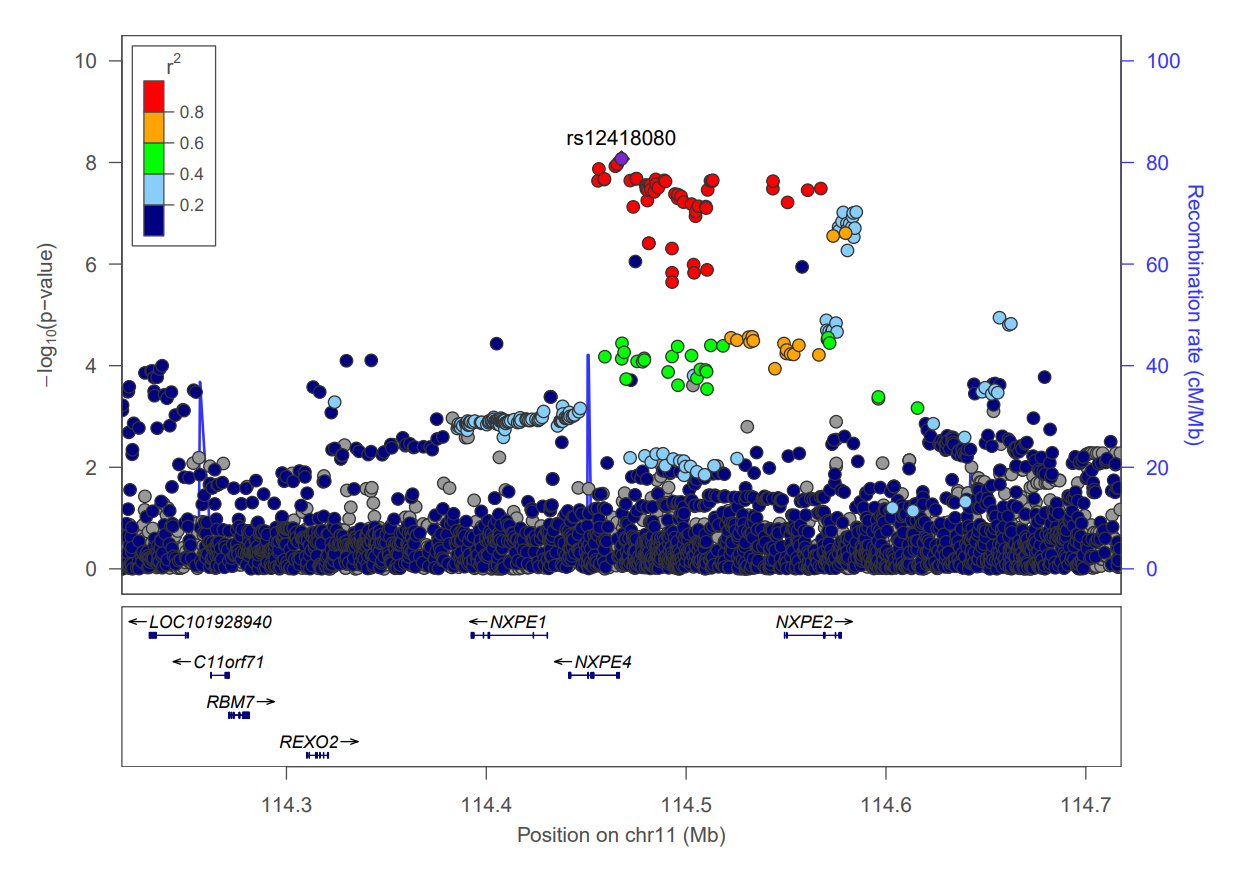

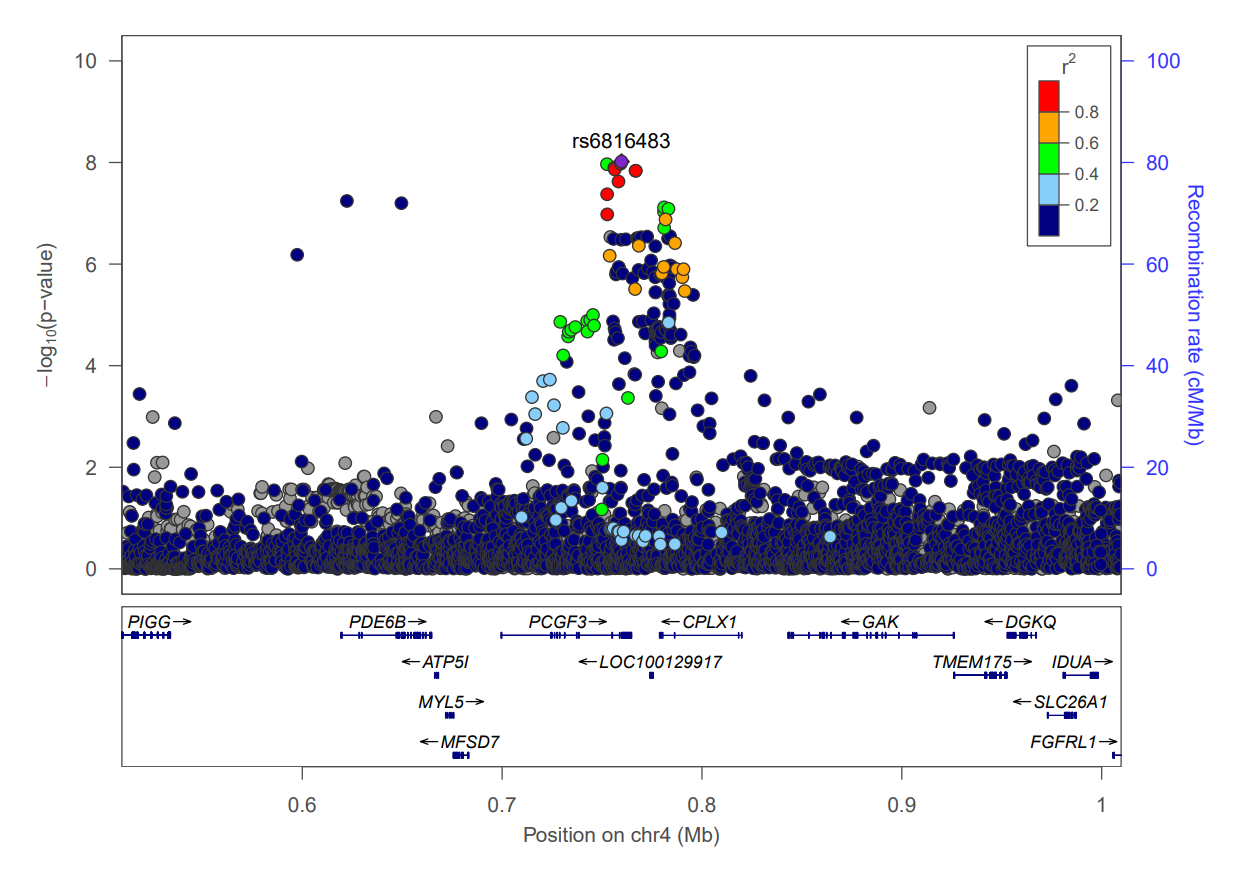

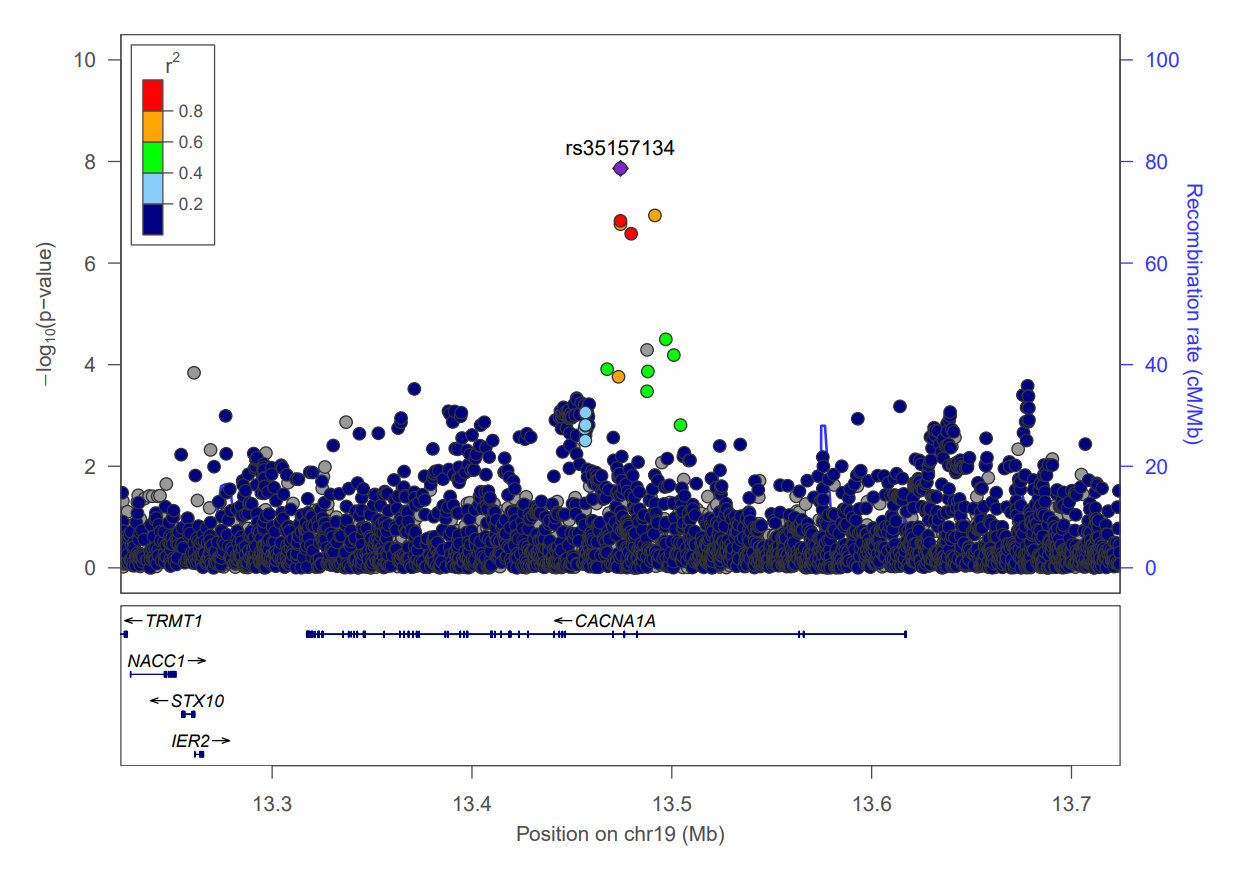

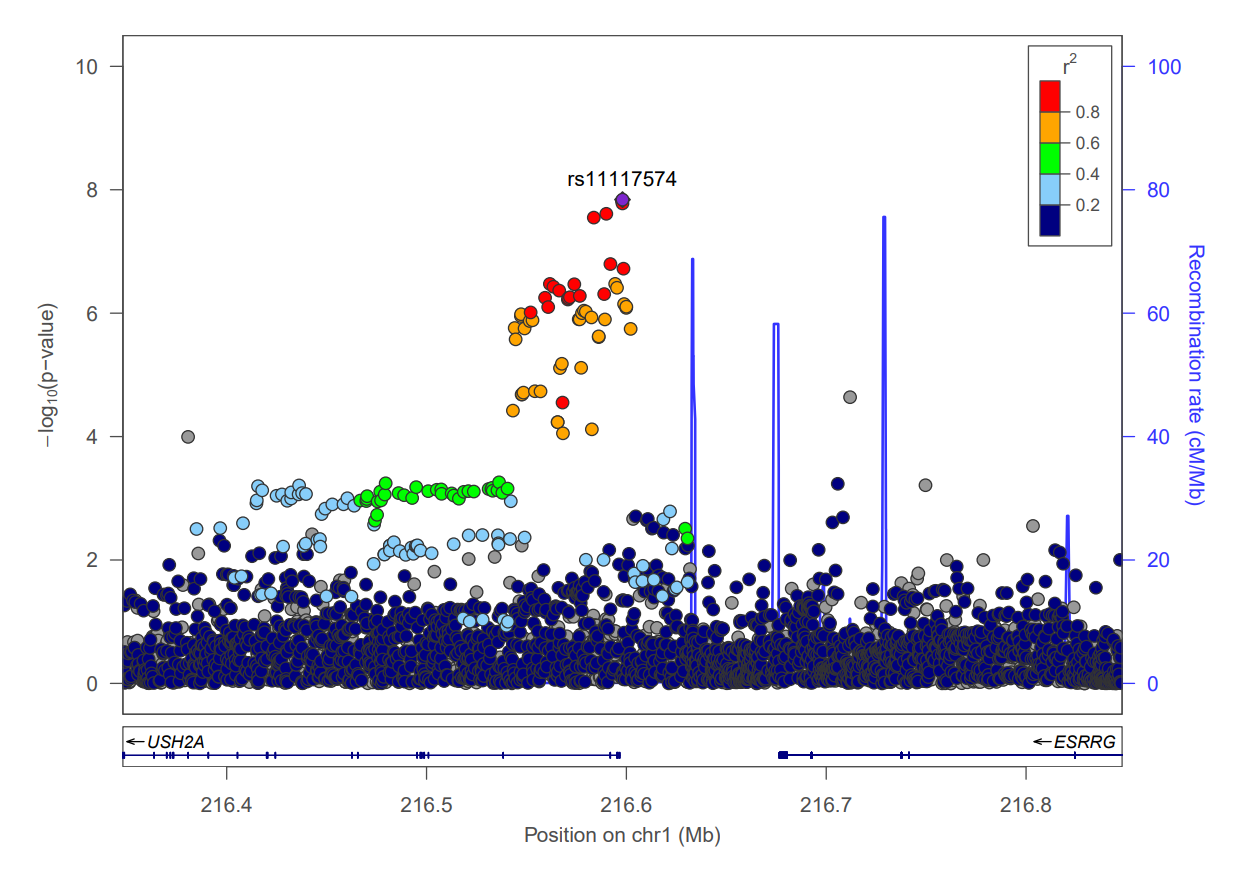

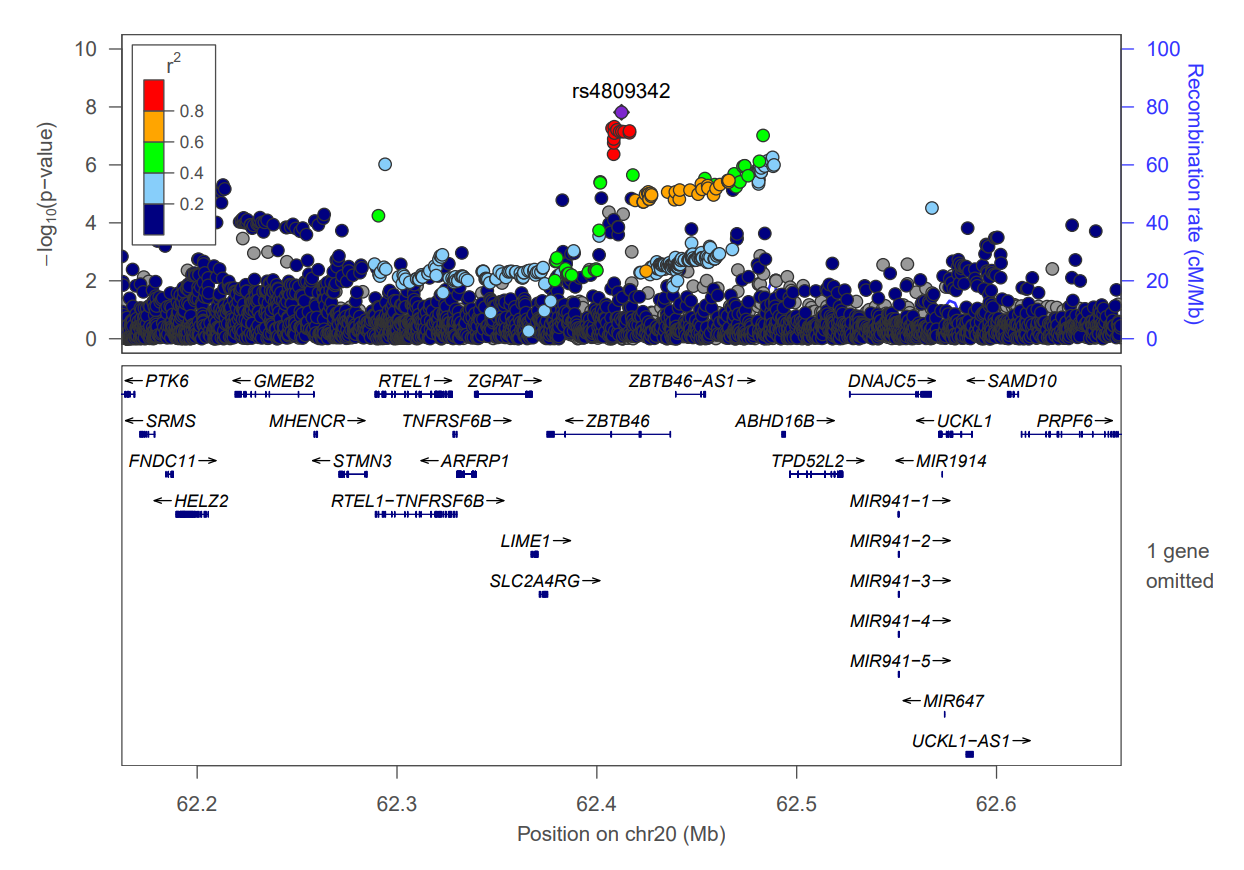

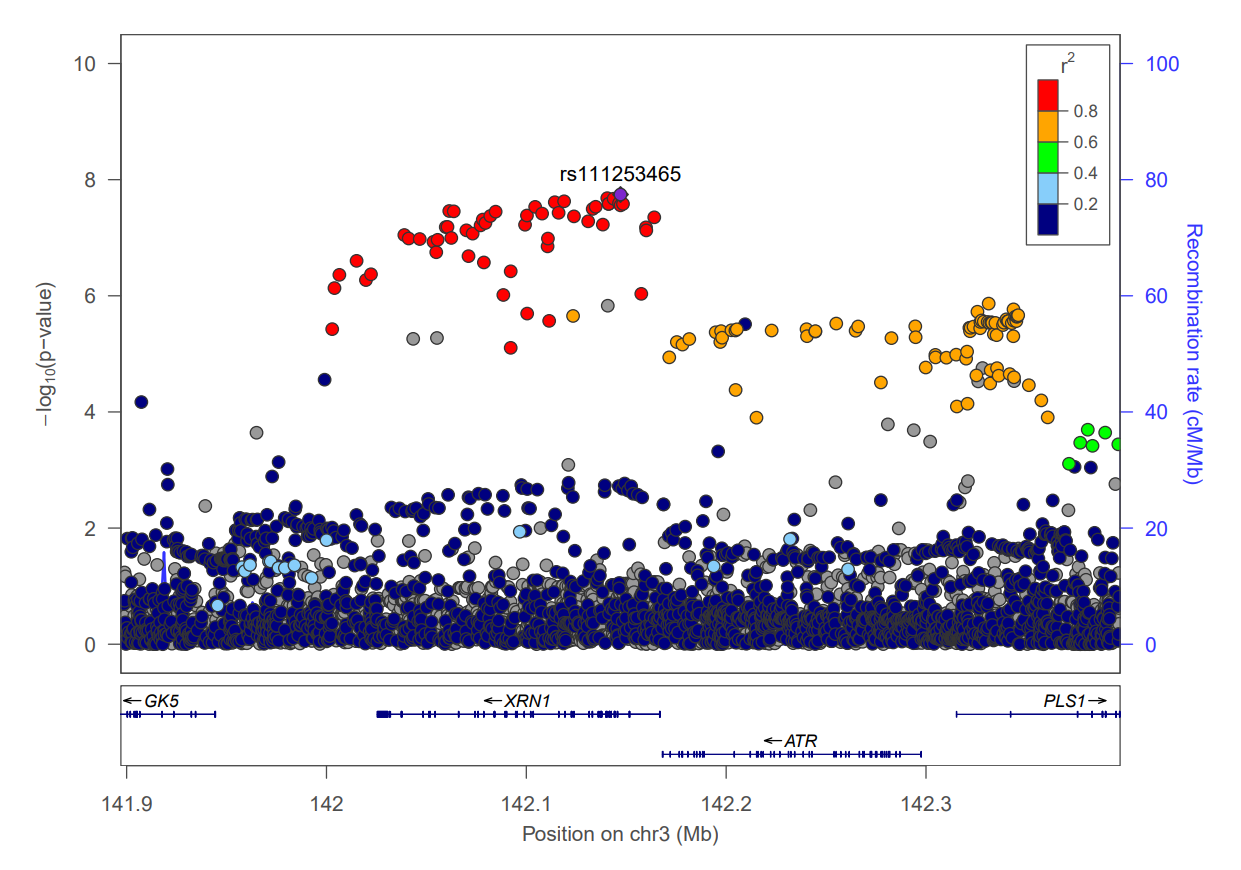

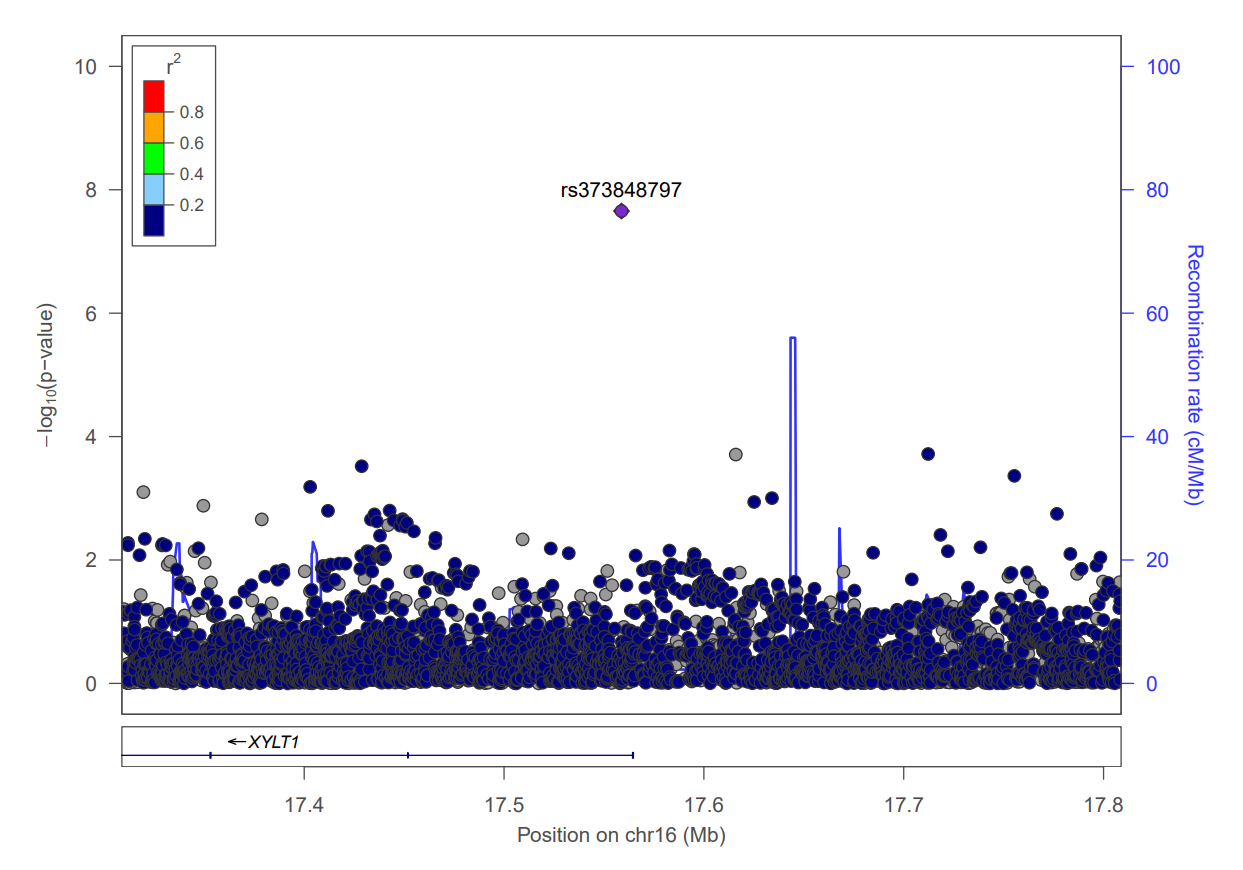

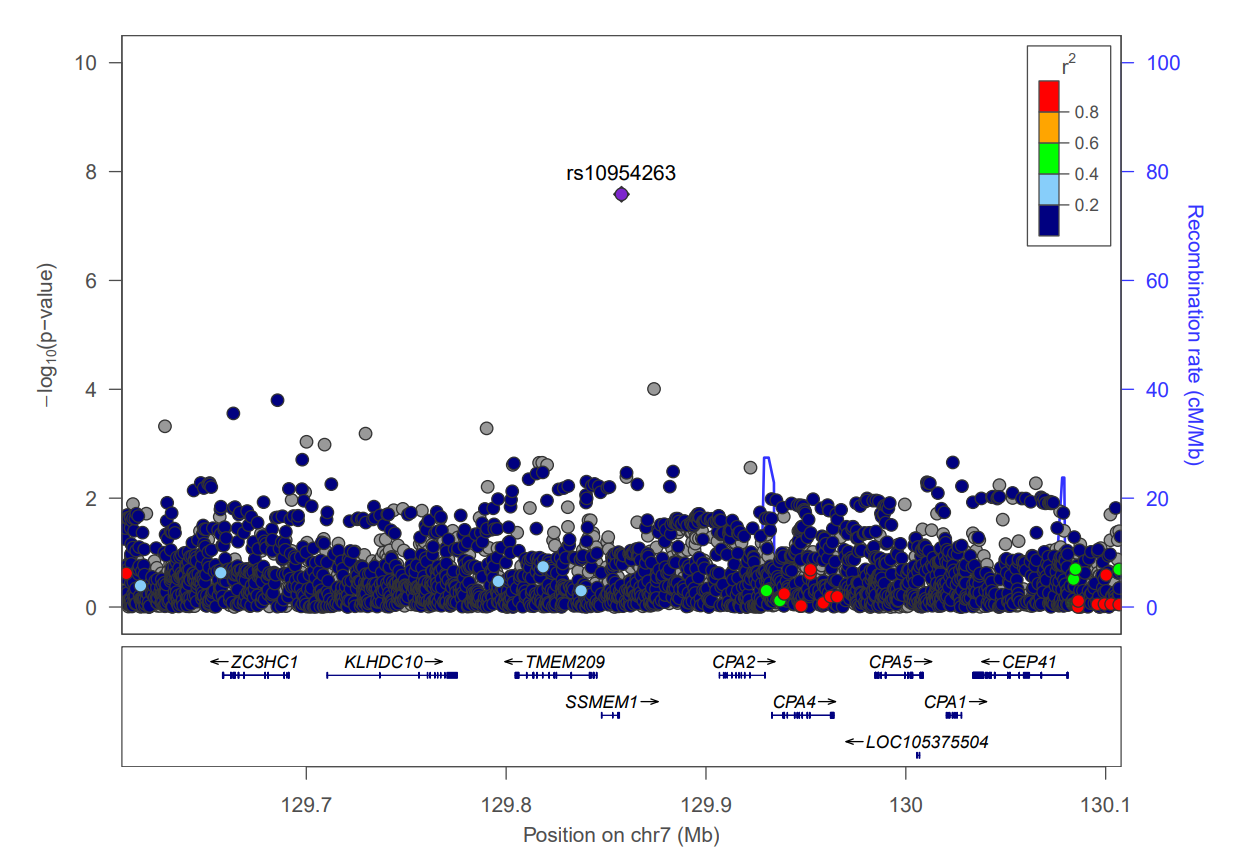

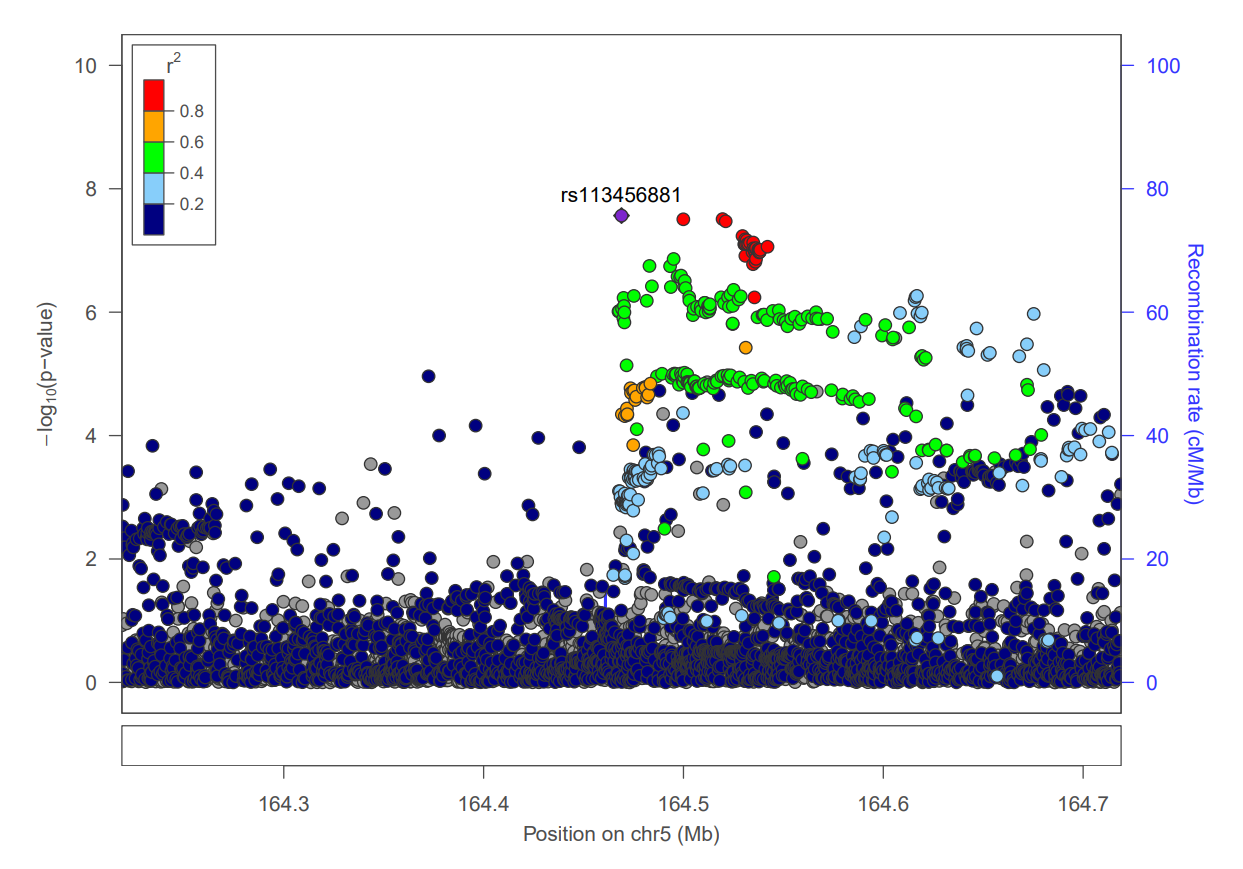

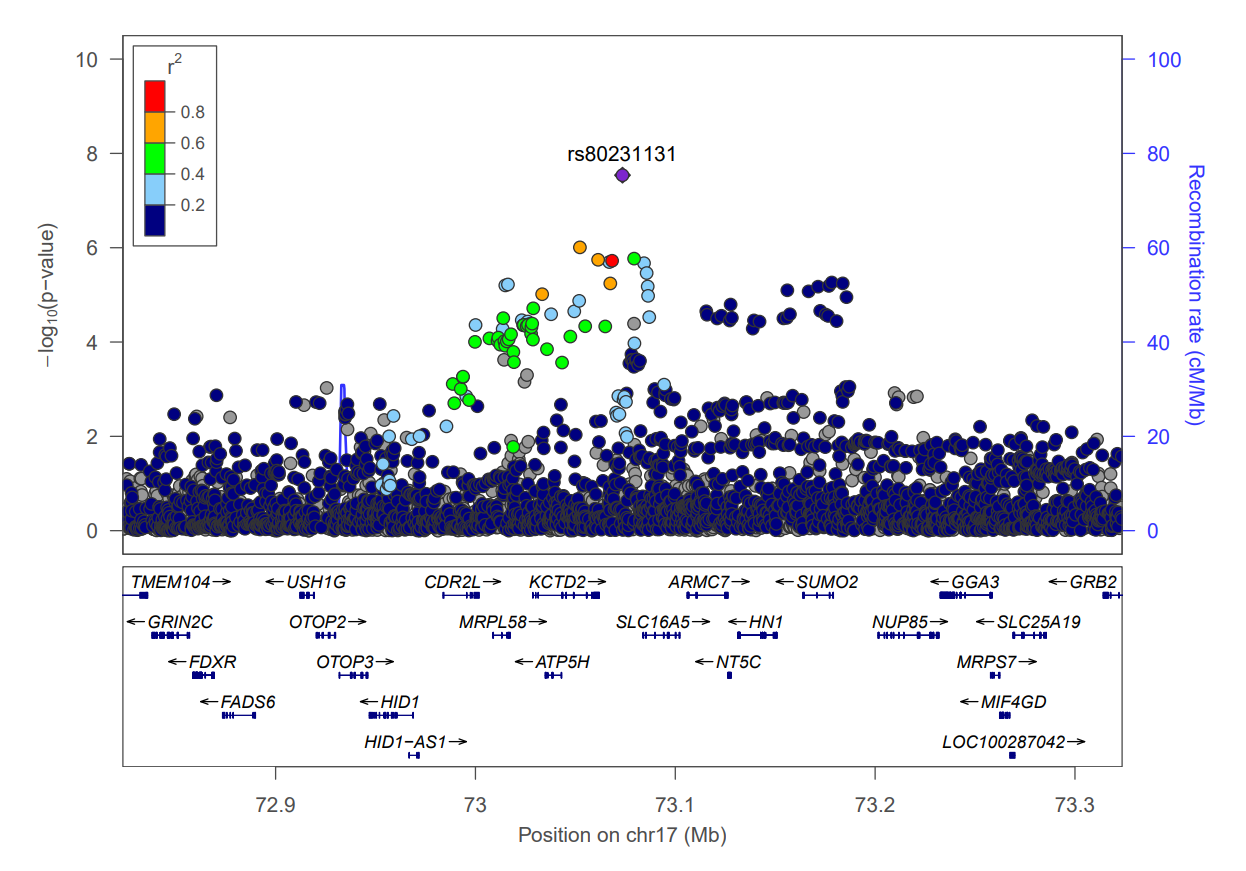

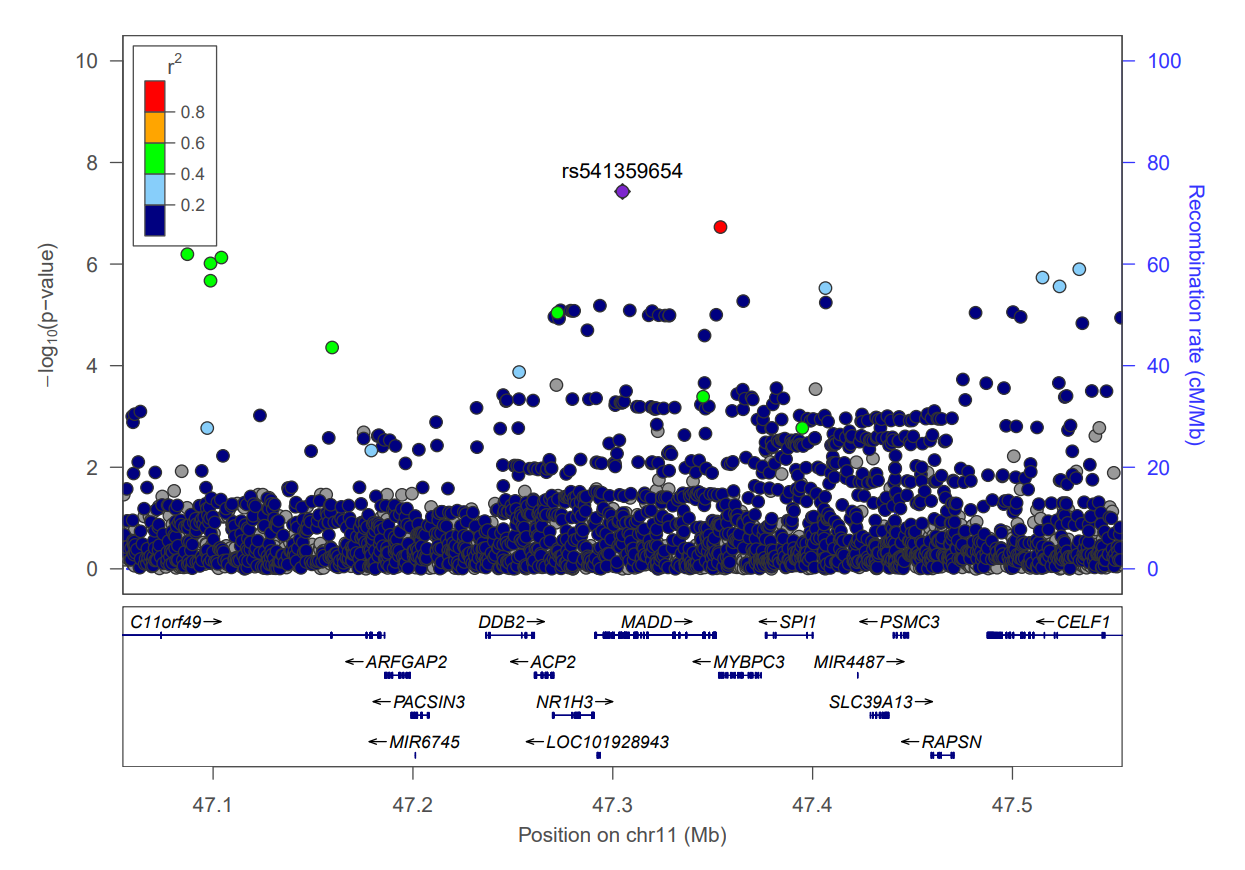

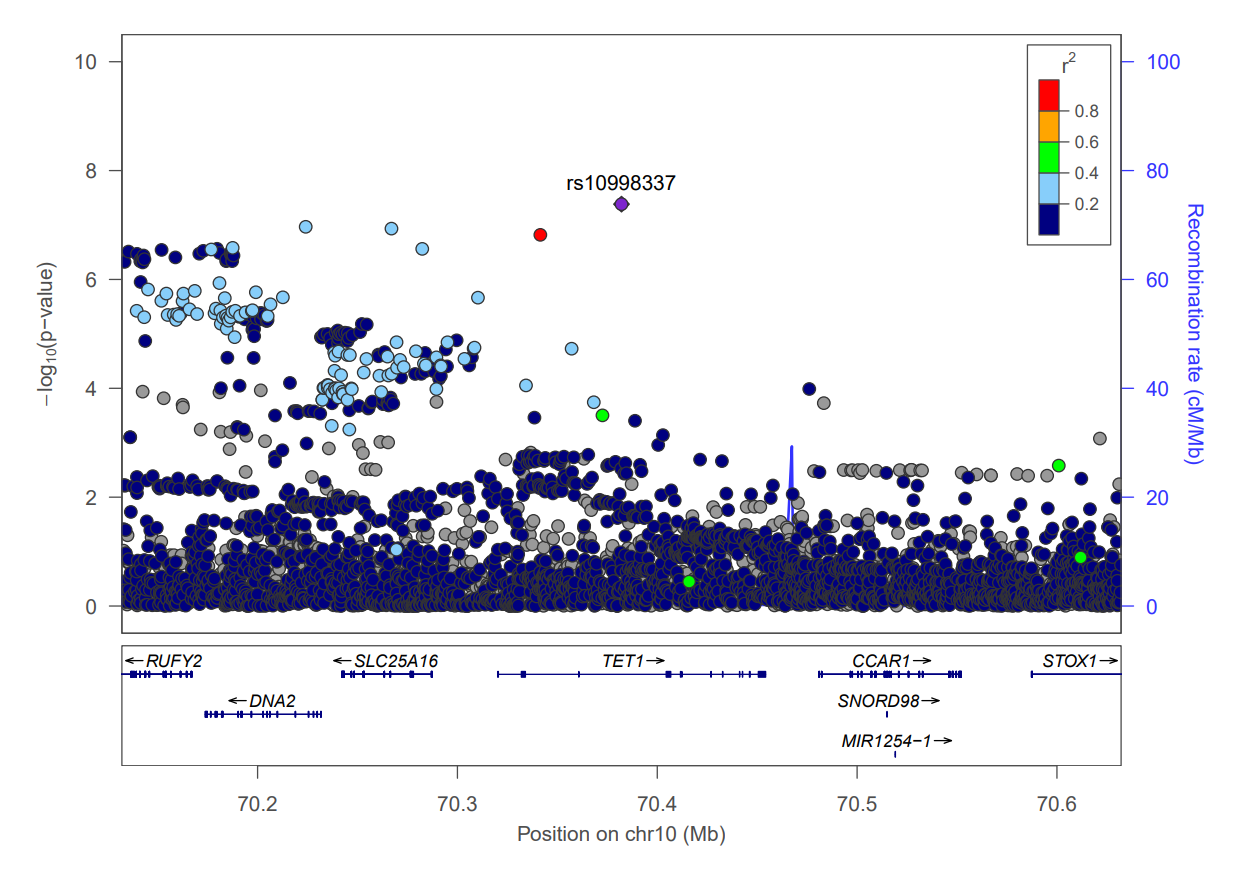

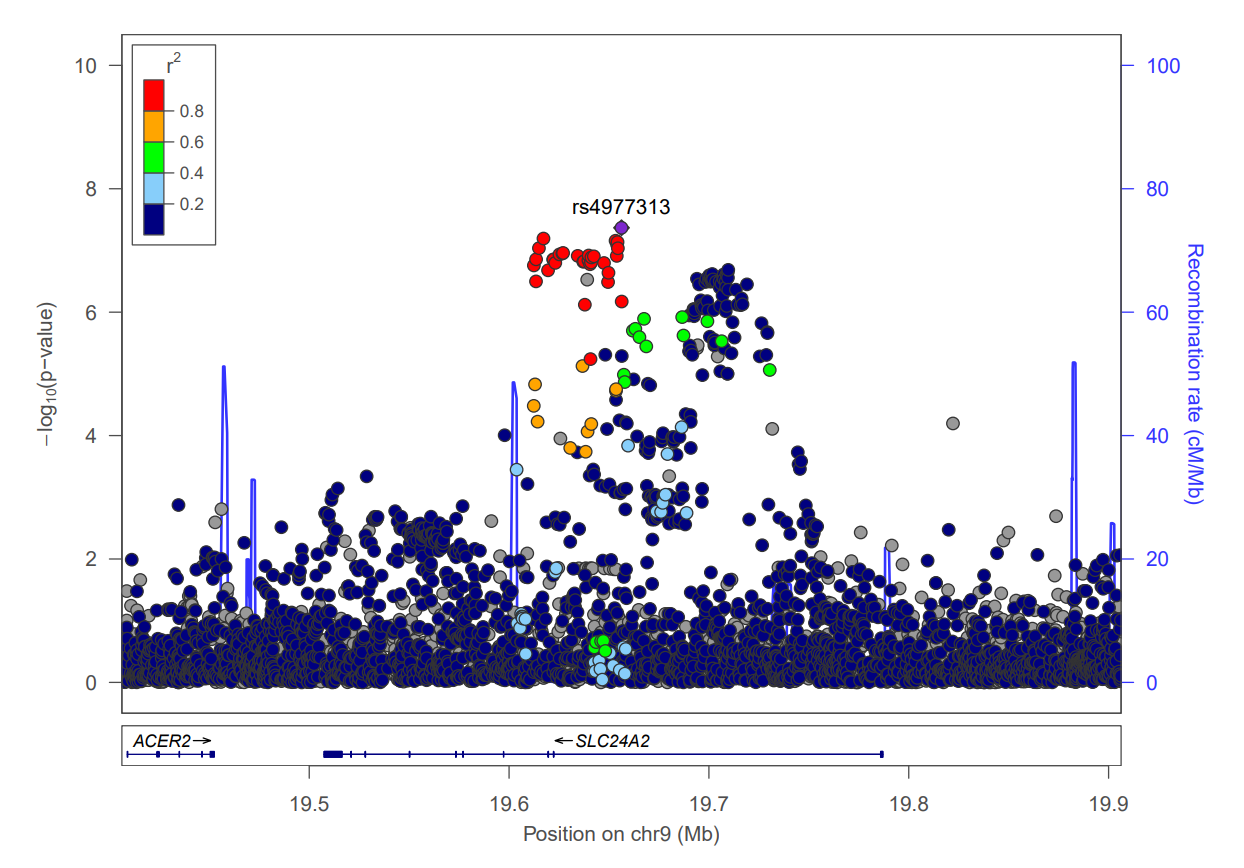

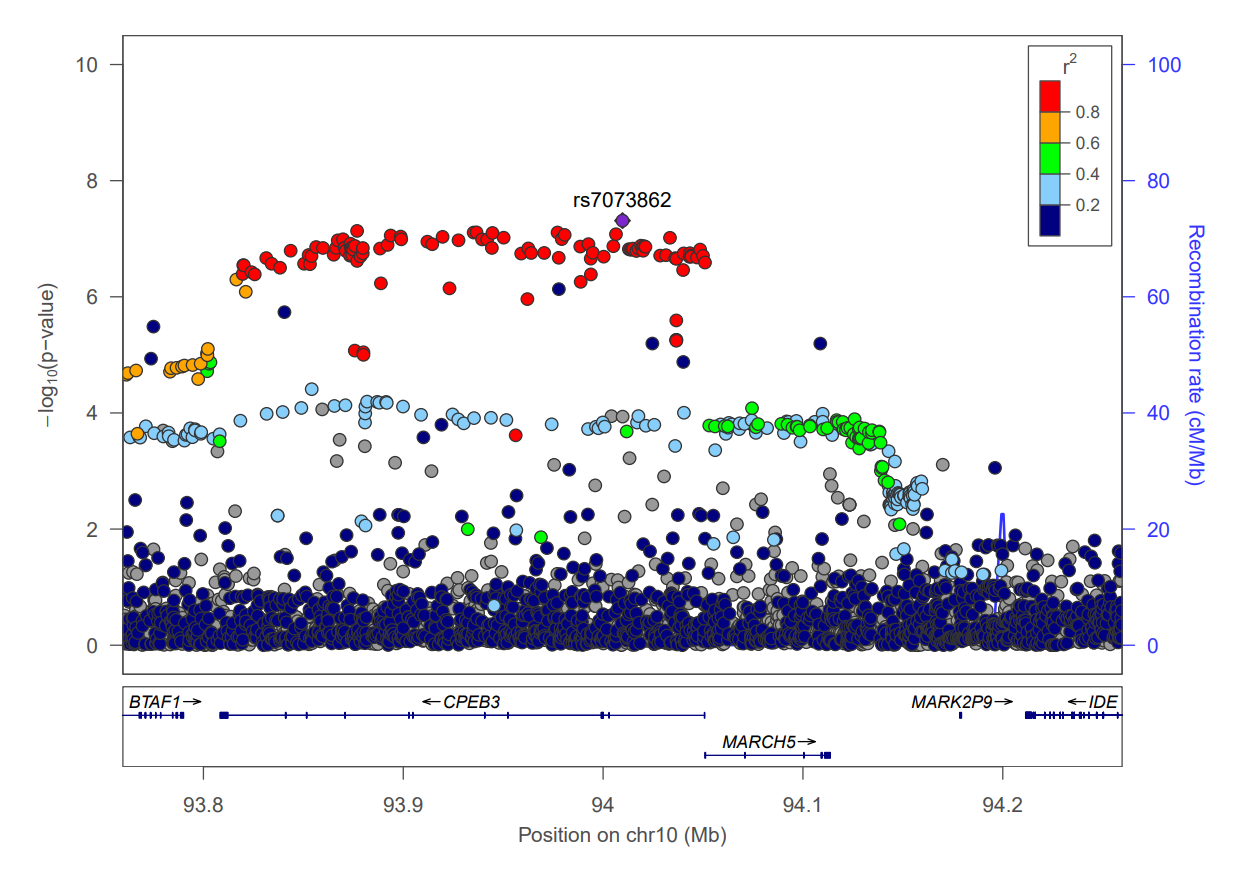

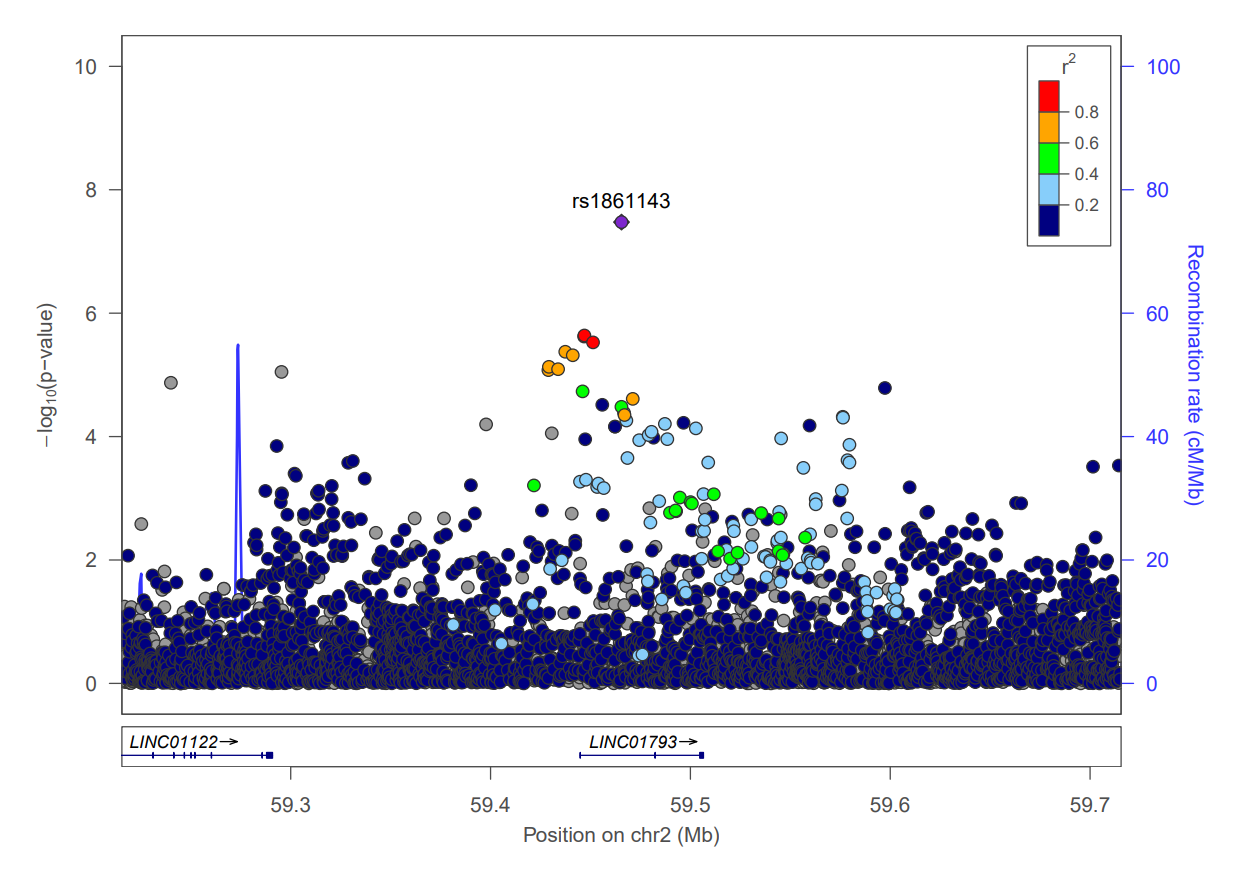

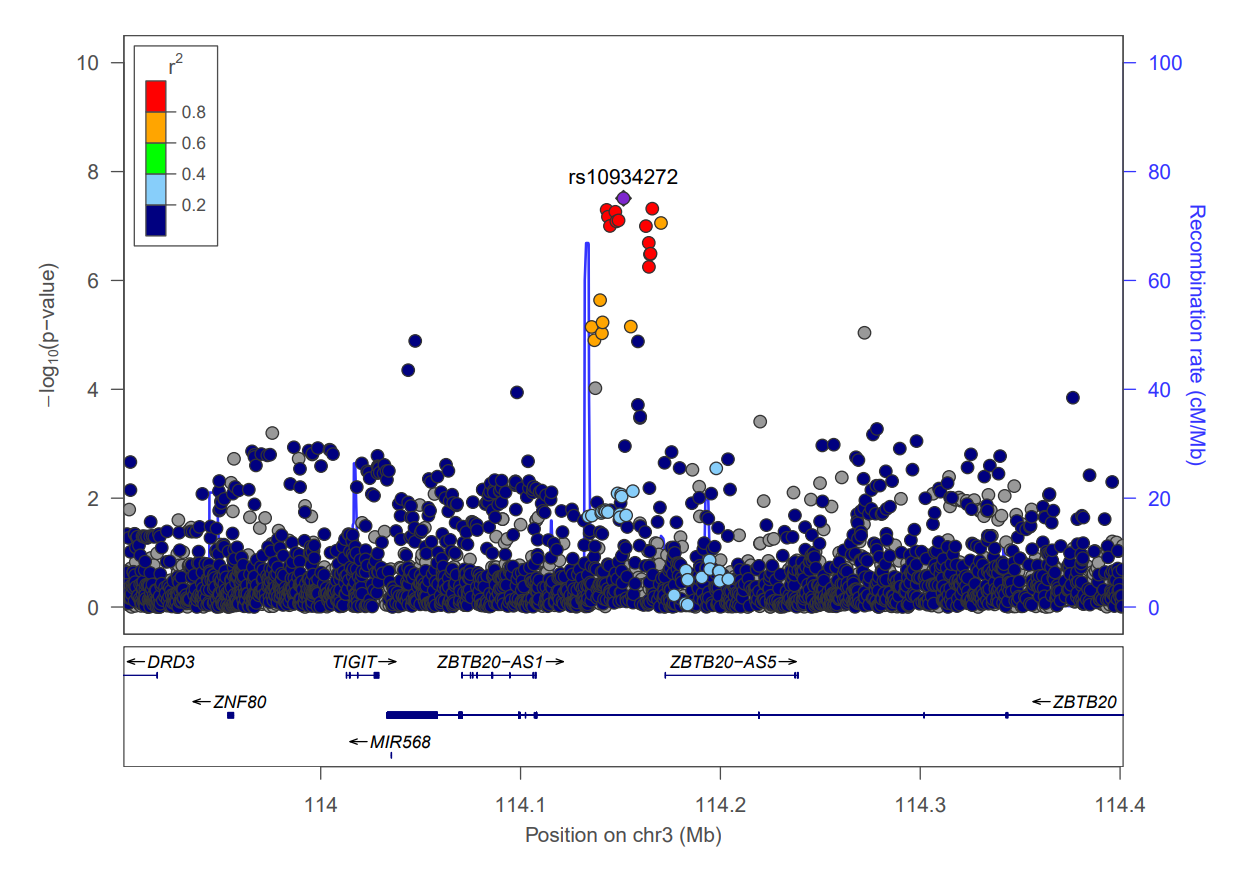

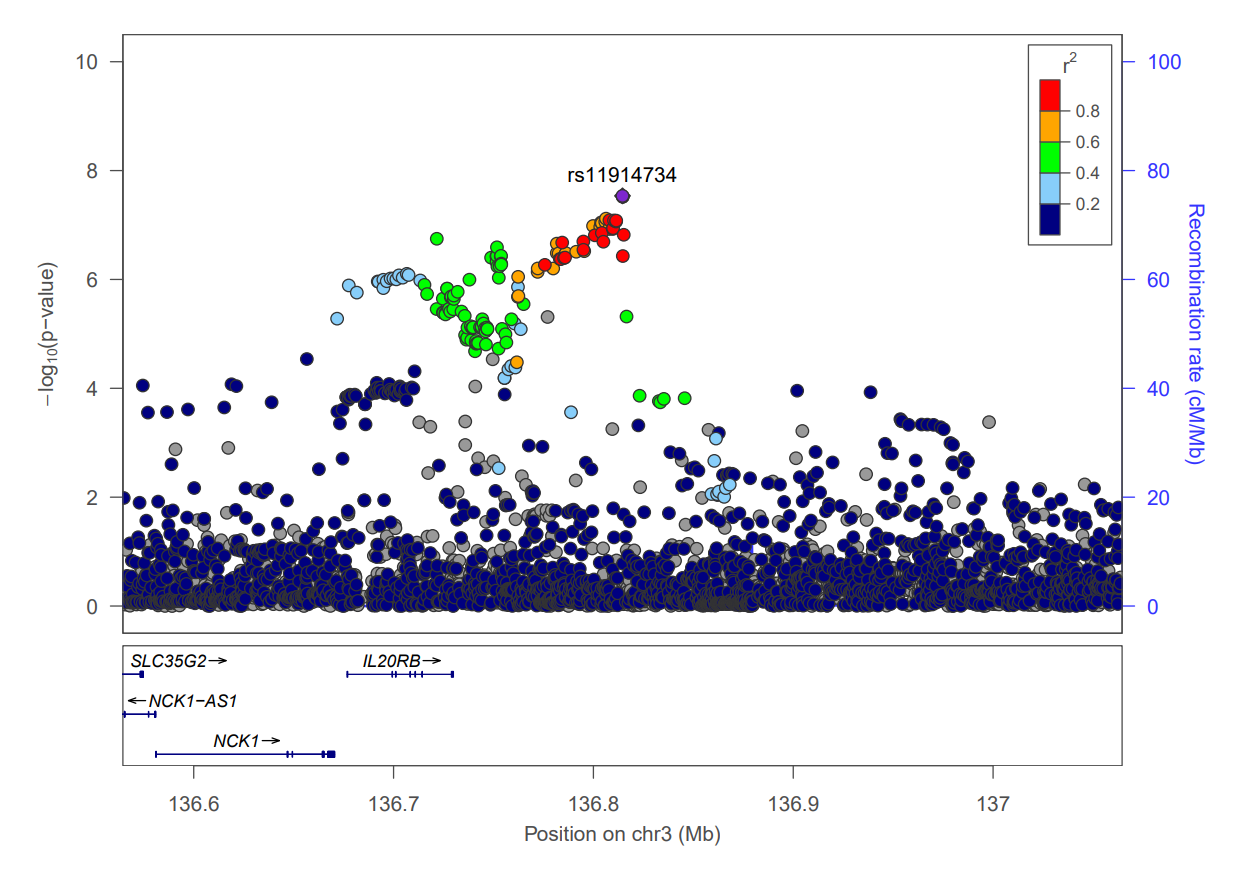

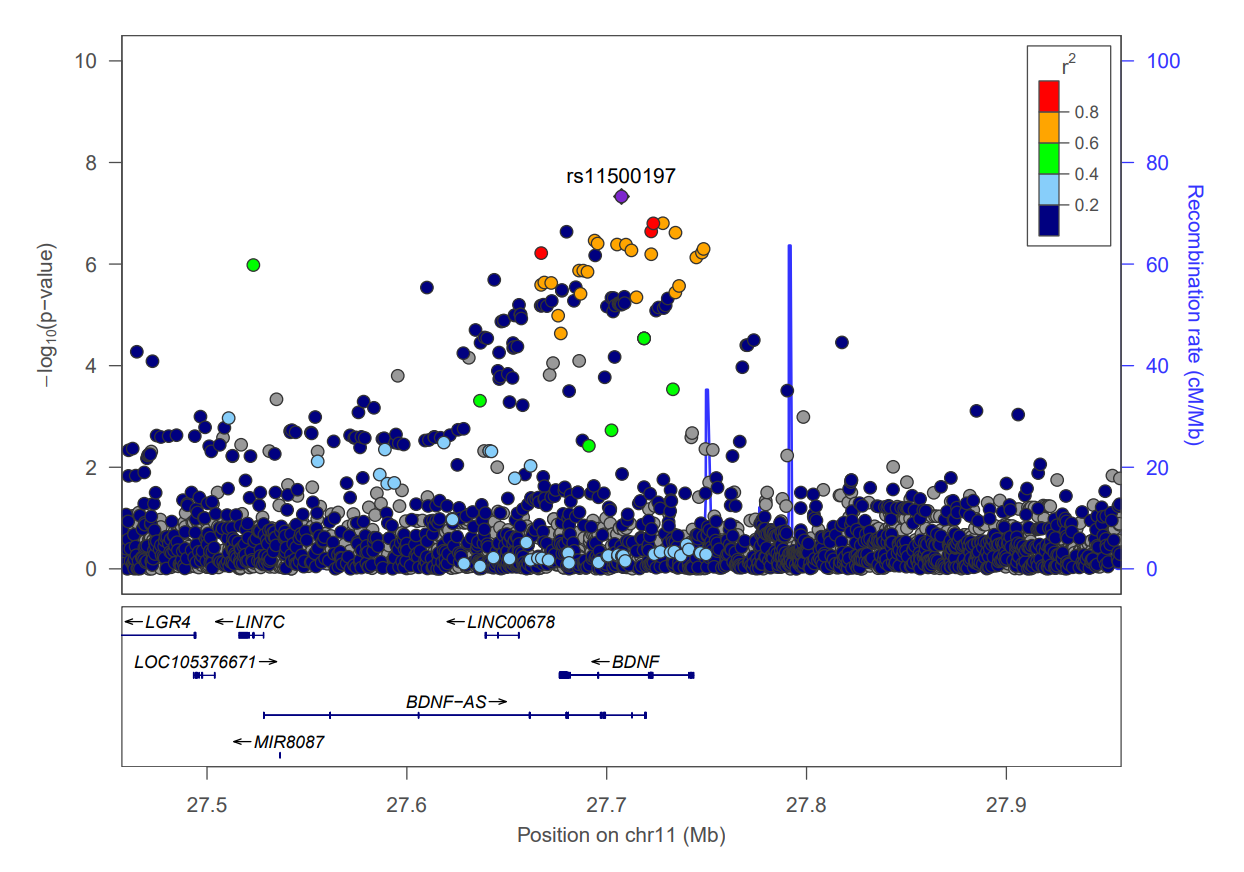

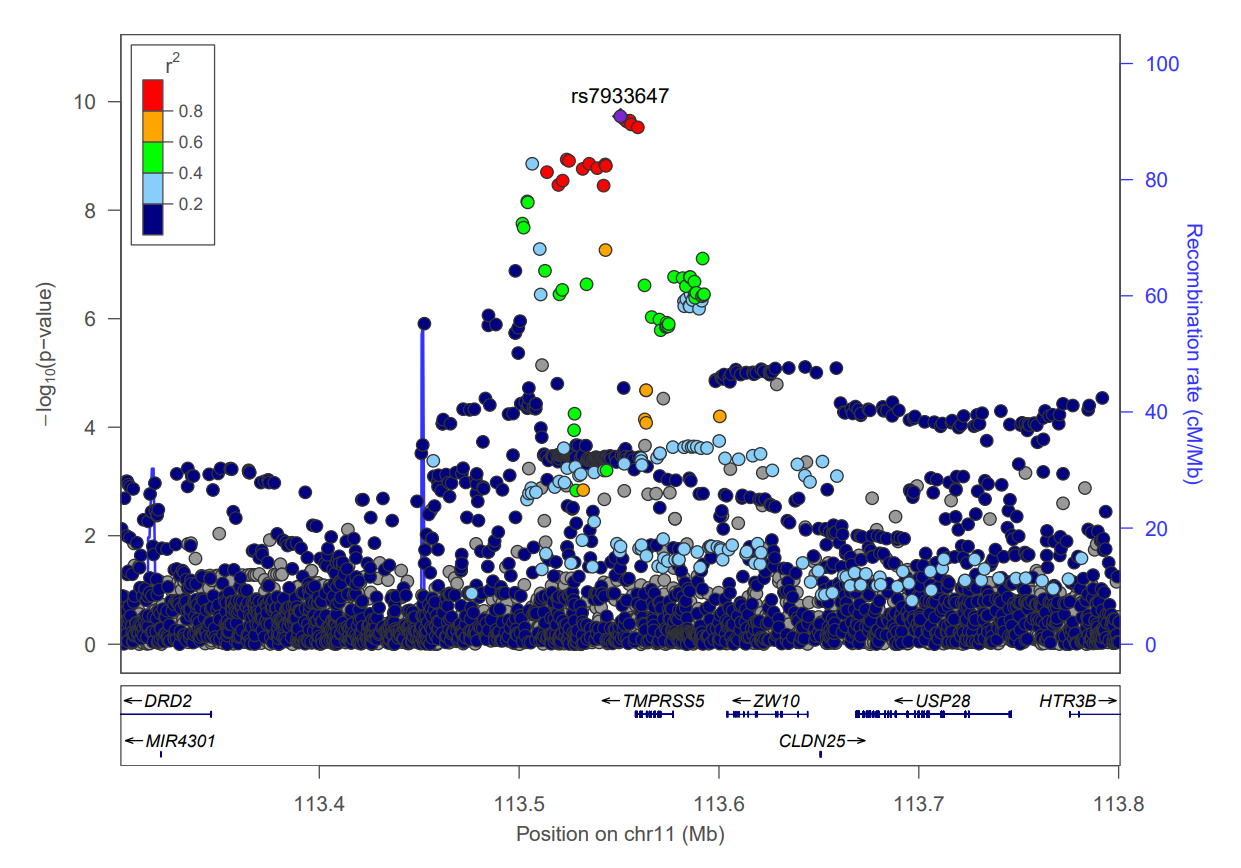

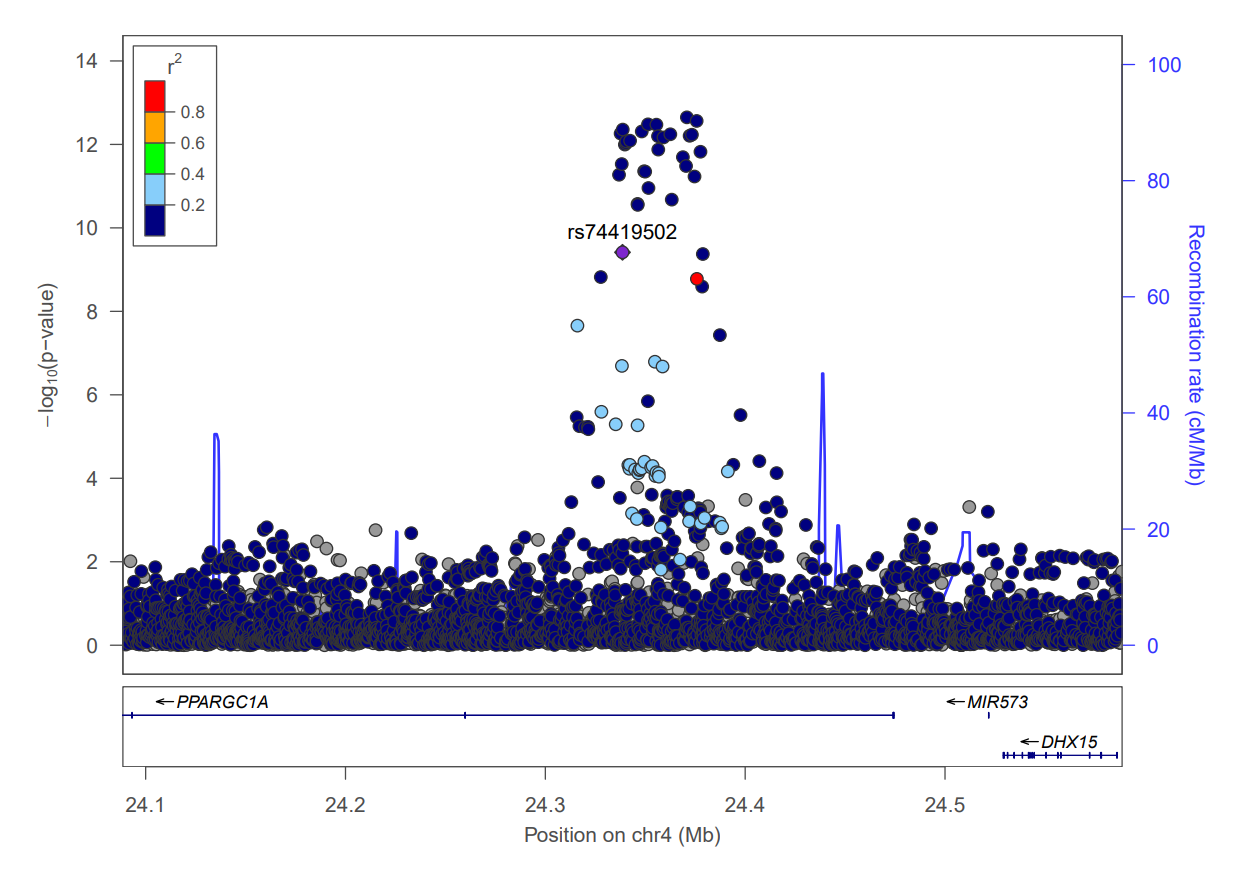

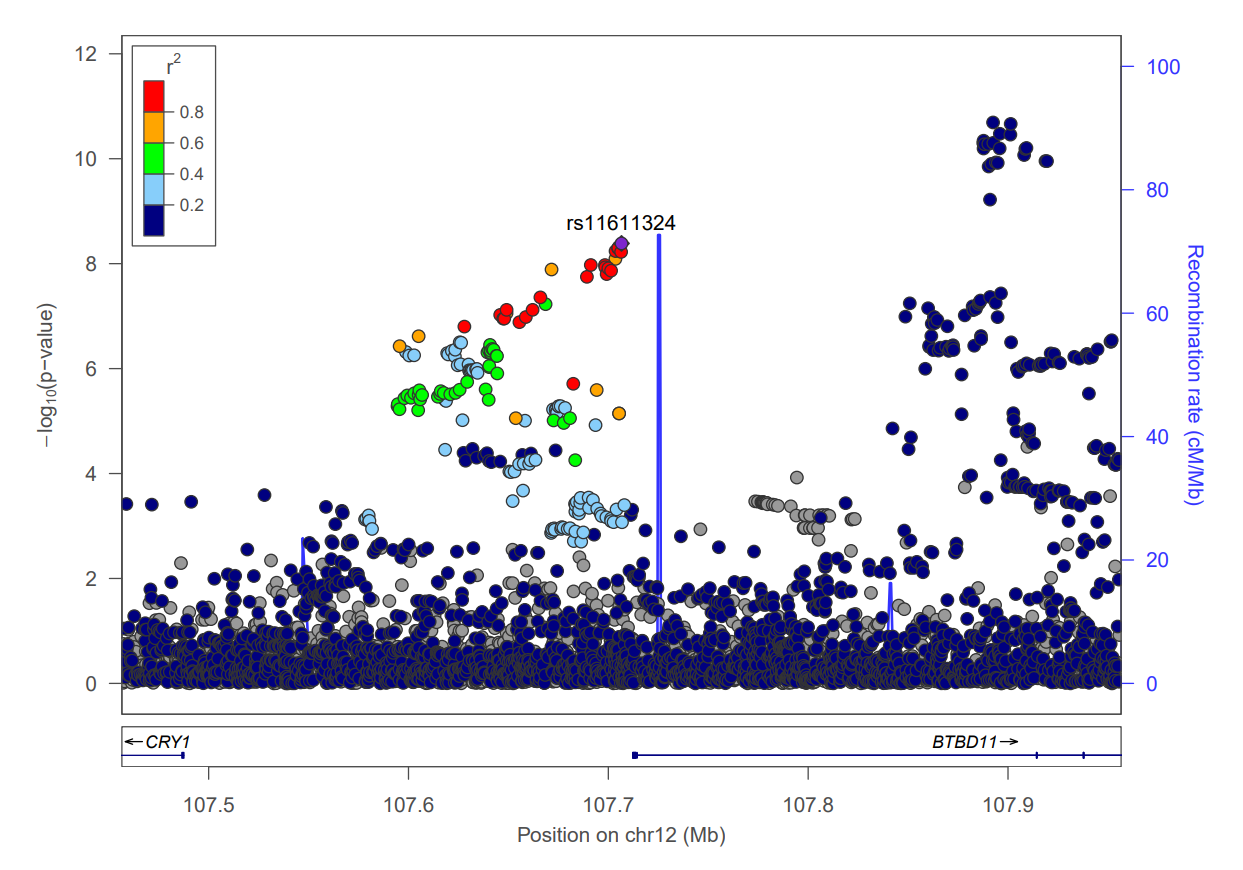

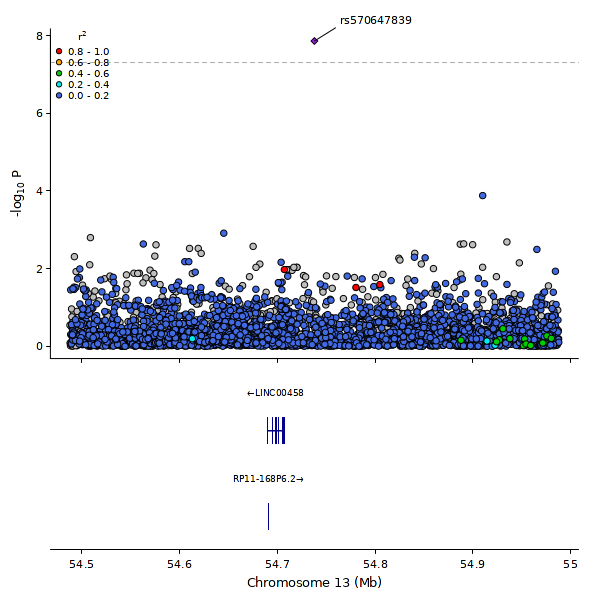

**Supplementary Figure 6. Regional plots of the loci associated with ET - Novel associations.** Each point represents a variant plotted by its chromosomal position (x-axis) and –log₁₀(P-value) (left y-axis), derived from a two-sided logistic regression likelihood-ratio test. Variants are colored by the degree of linkage disequilibrium (r²) with the lead variant (highlighted in red). Variants with predicted functional impact are indicated by their shape: squares represent moderate-impact variants, and diamonds represent high-impact variants. The recombination rate estimated from Icelandic data is shown as a solid blue line (right y-axis).

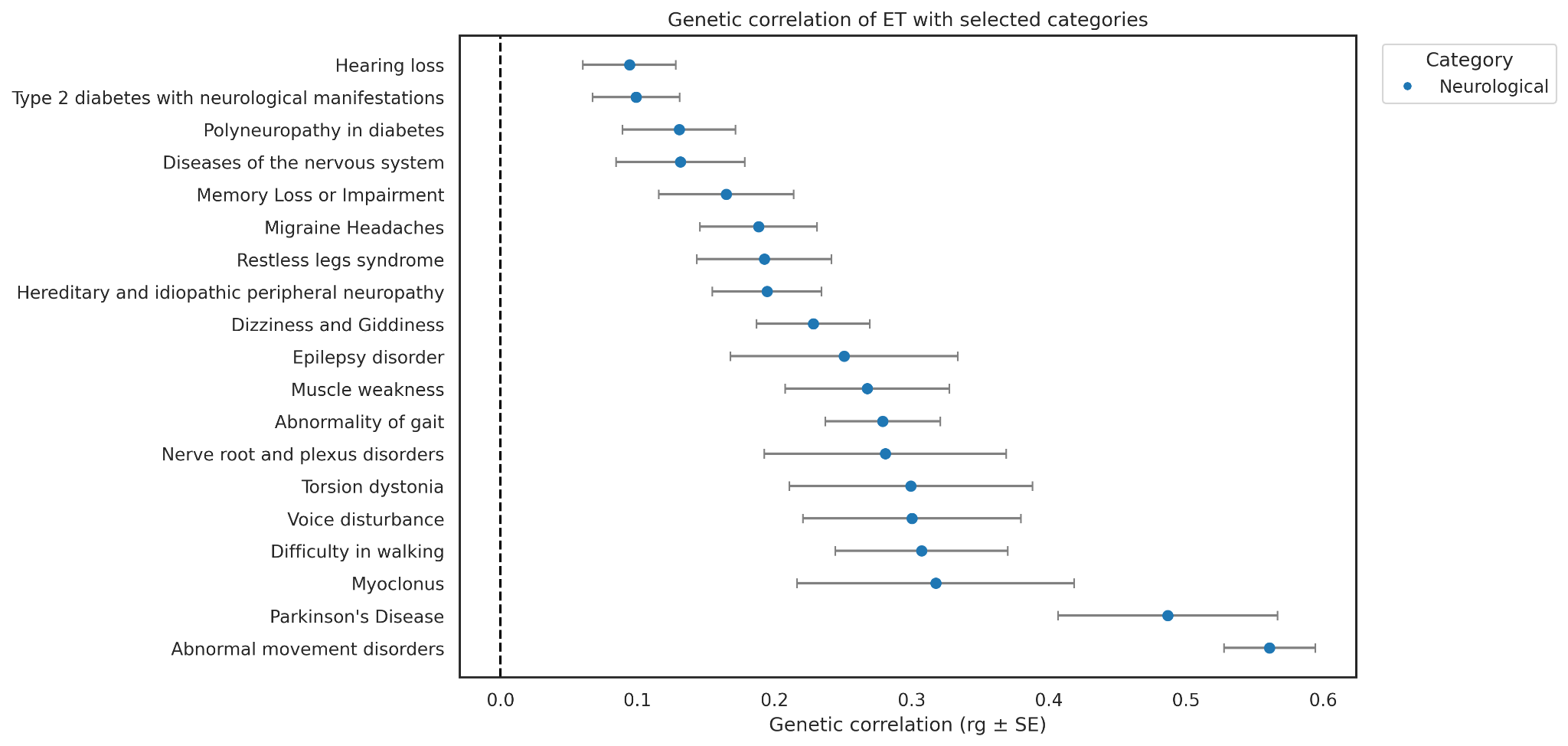

**Supplementary Figure 7. Genetic correlation between ET and Neurological traits**

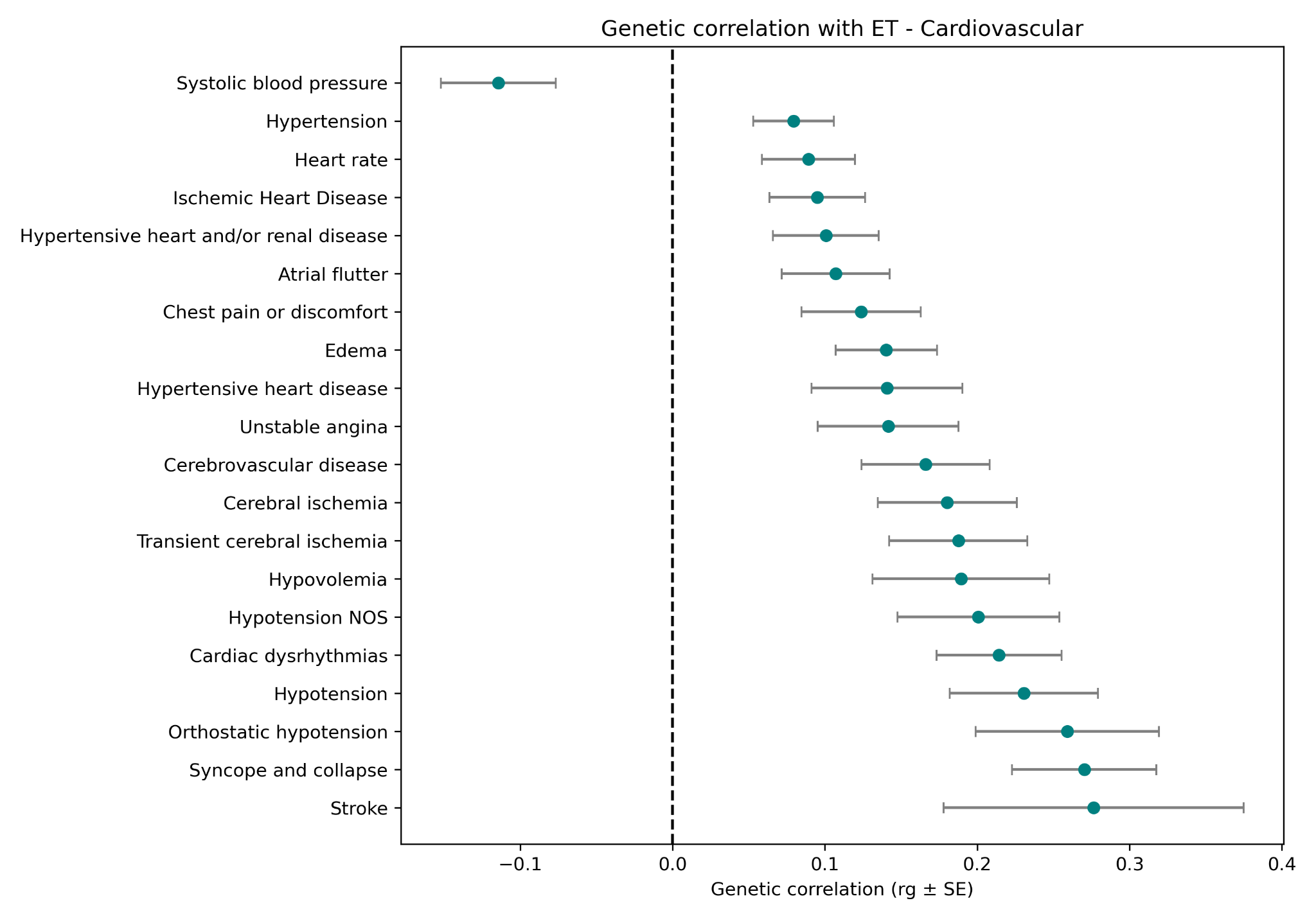

**Supplementary Figure 8. Genetic correlation between ET and Cardiovascular traits**

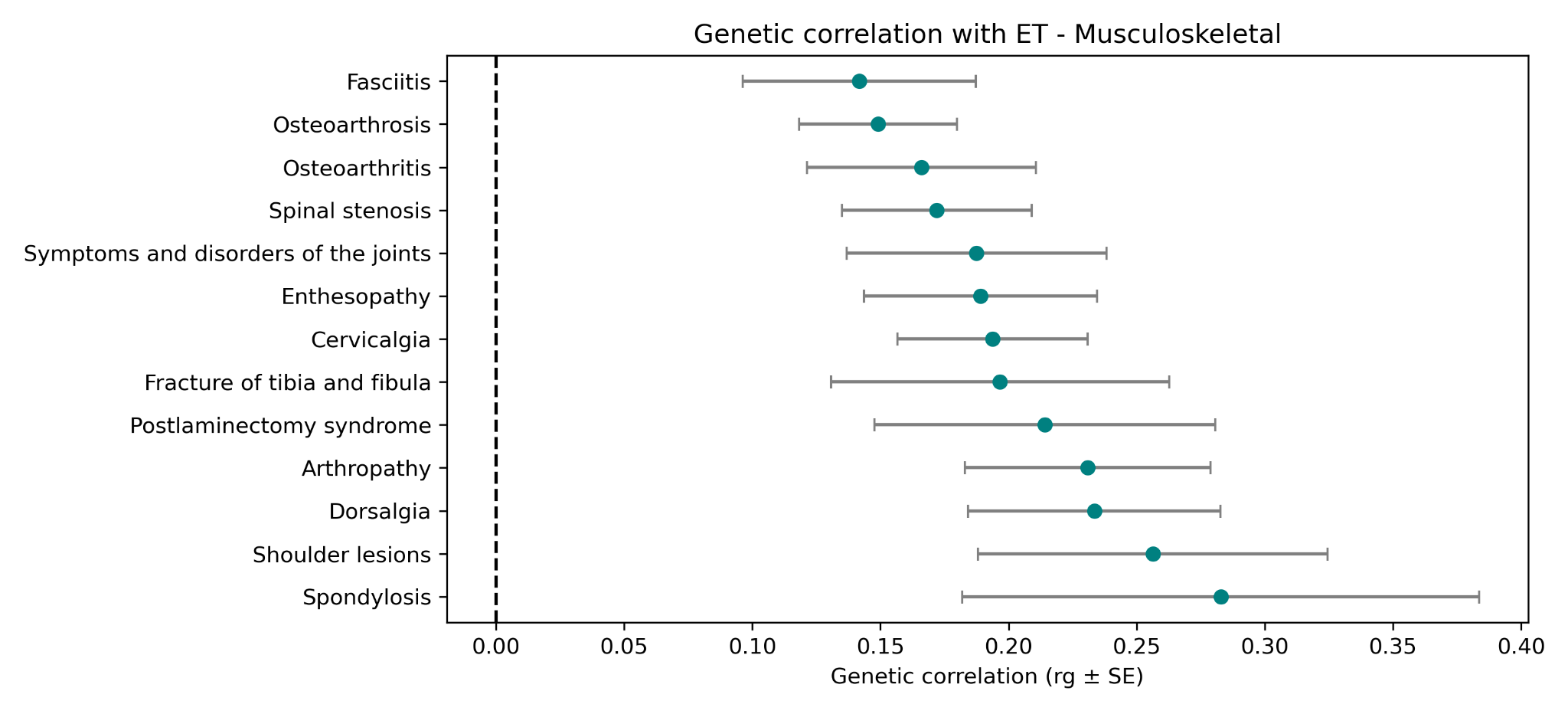

**Supplementary Figure 9. Genetic correlation between ET and Musculoskeletal traits**

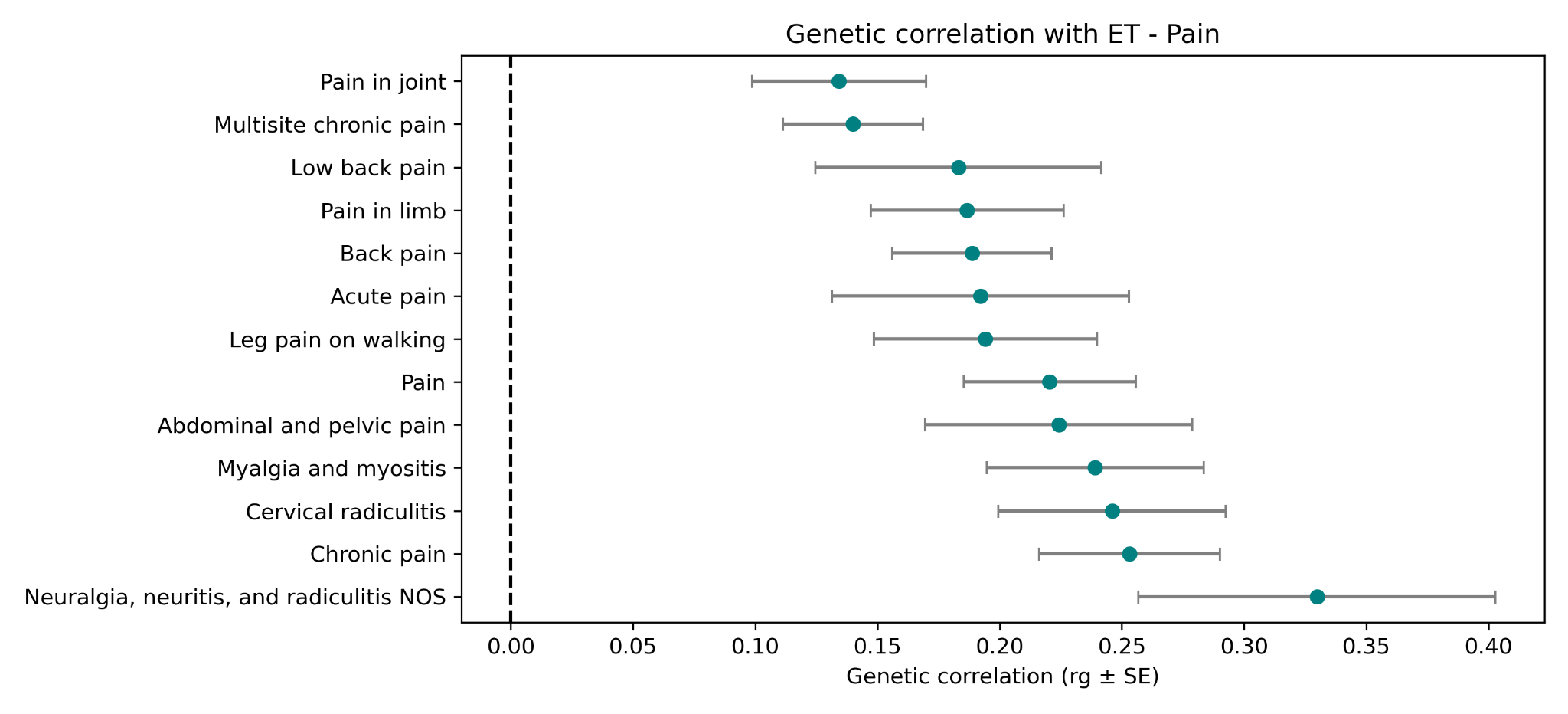

**Supplementary Figure 10. Genetic correlation between ET and Pain conditions**

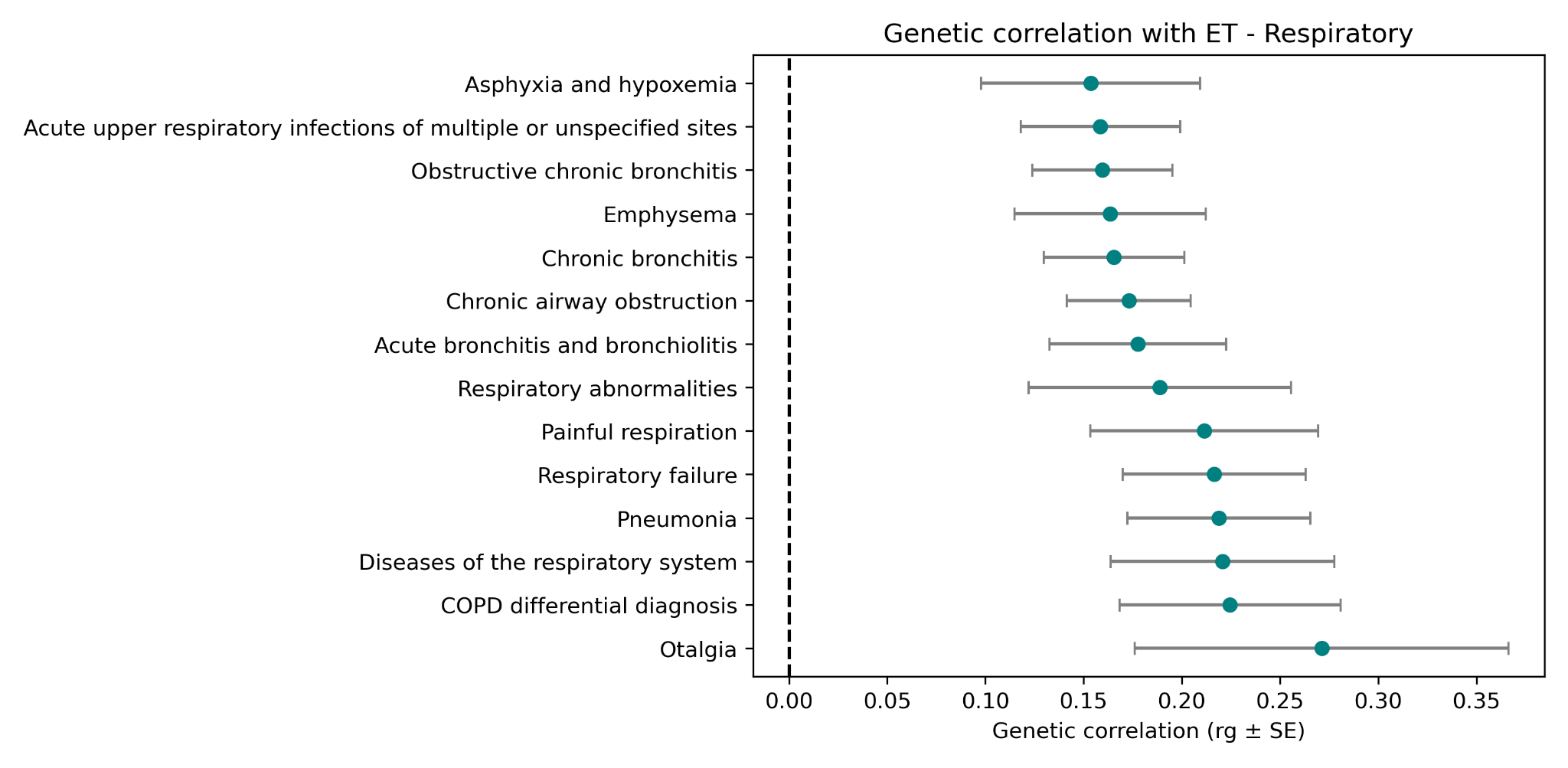

**Supplementary Figure 11. Genetic correlation between ET and Respiratory traits**

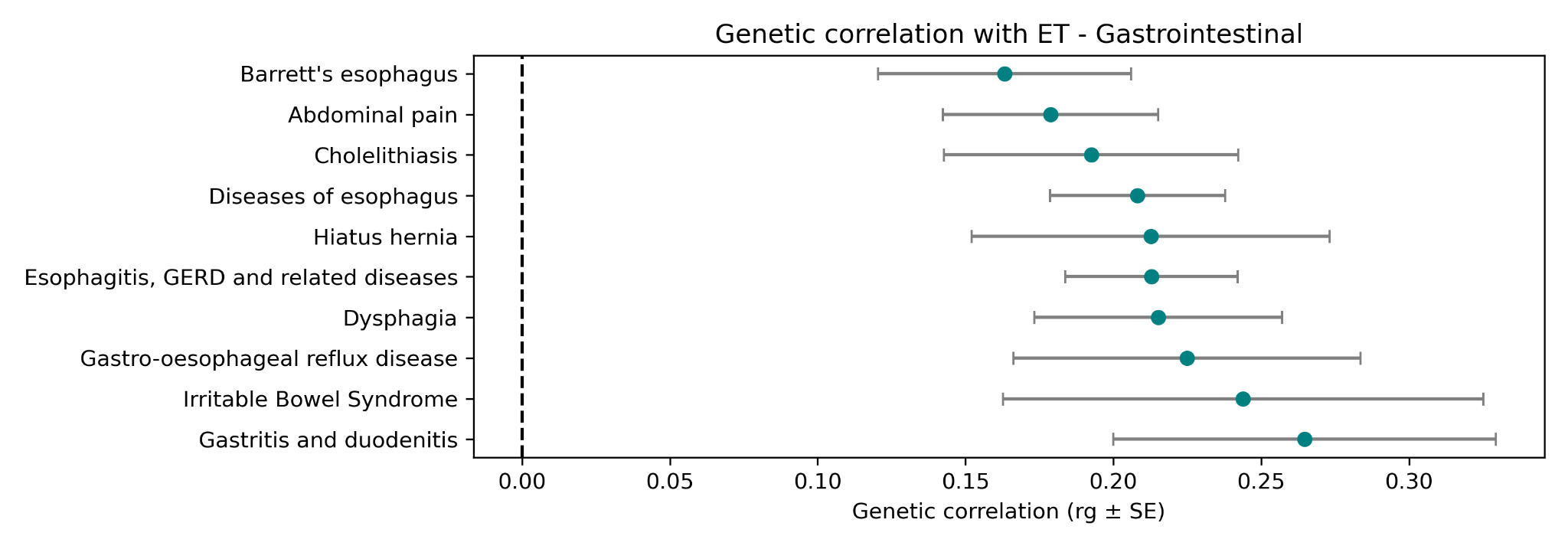

**Supplementary Figure 12. Genetic correlation between ET and Gastrointestinal traits**

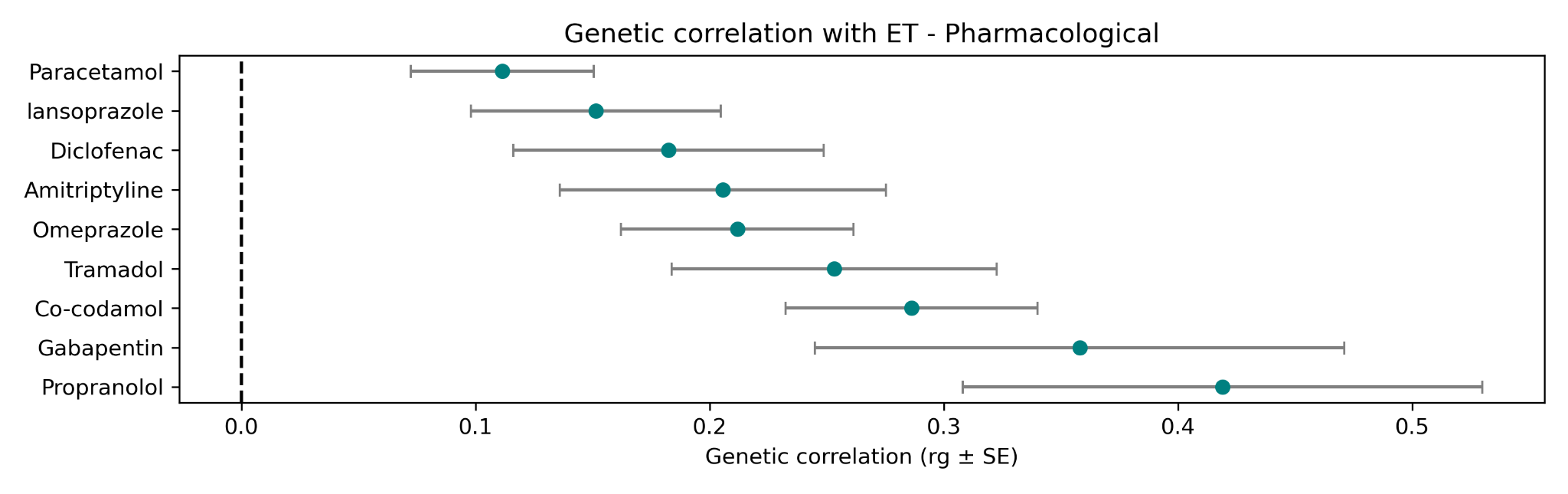

**Supplementary Figure 13. Genetic correlation between ET and pharmacological treatment**

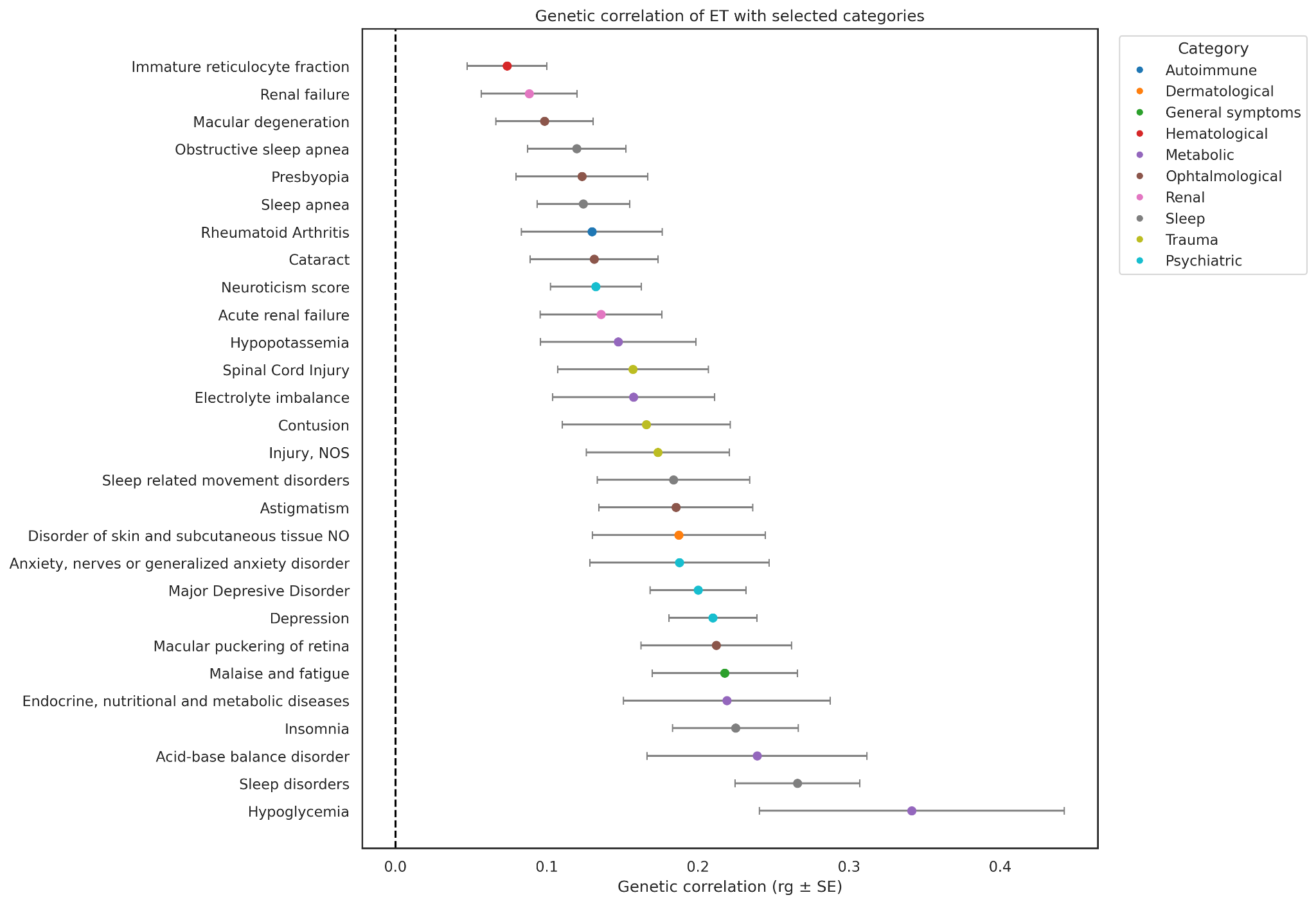

**Supplementary Figure 14. Genetic correlation between ET and other complex traits**

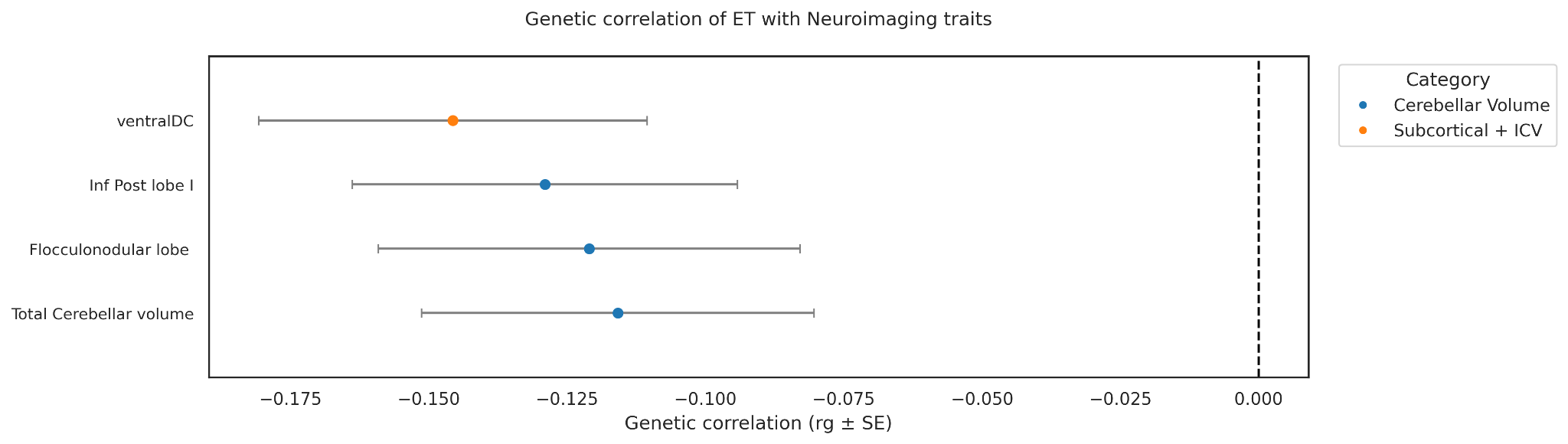

**Supplementary Figure 15. Genetic correlation between ET and brain volumes**

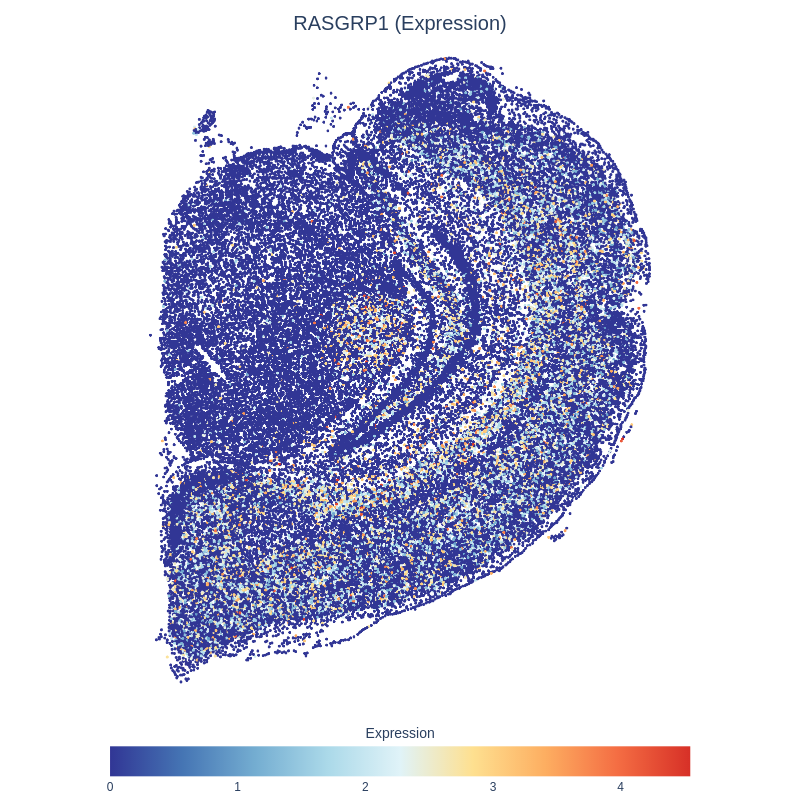

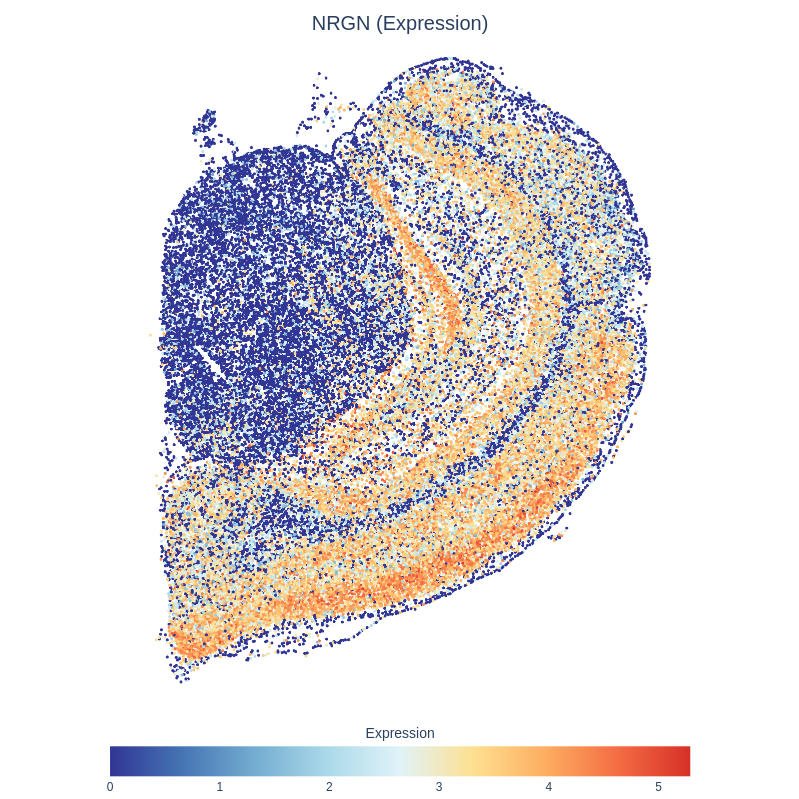

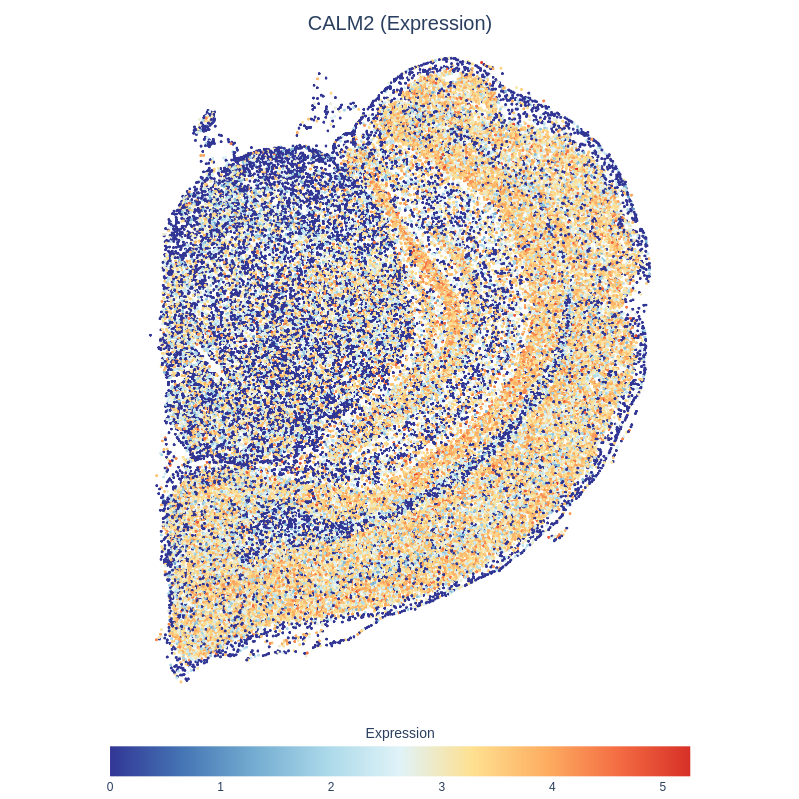

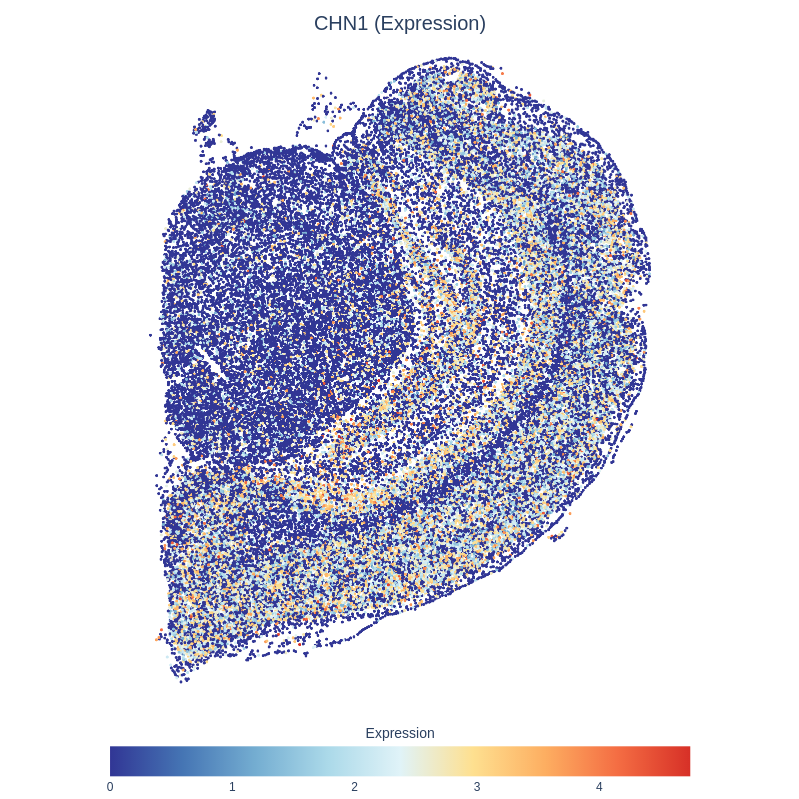

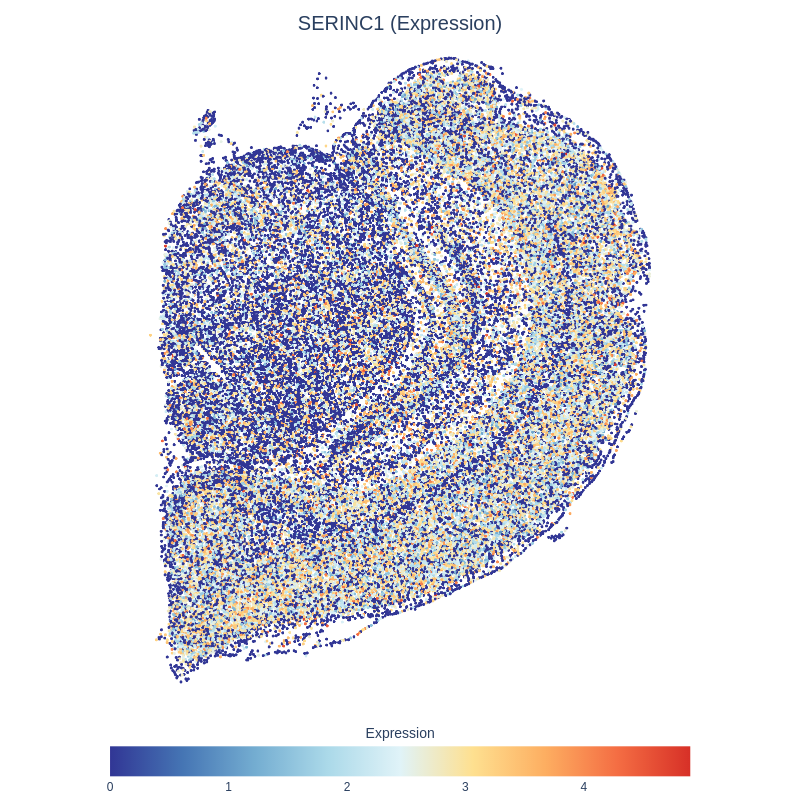

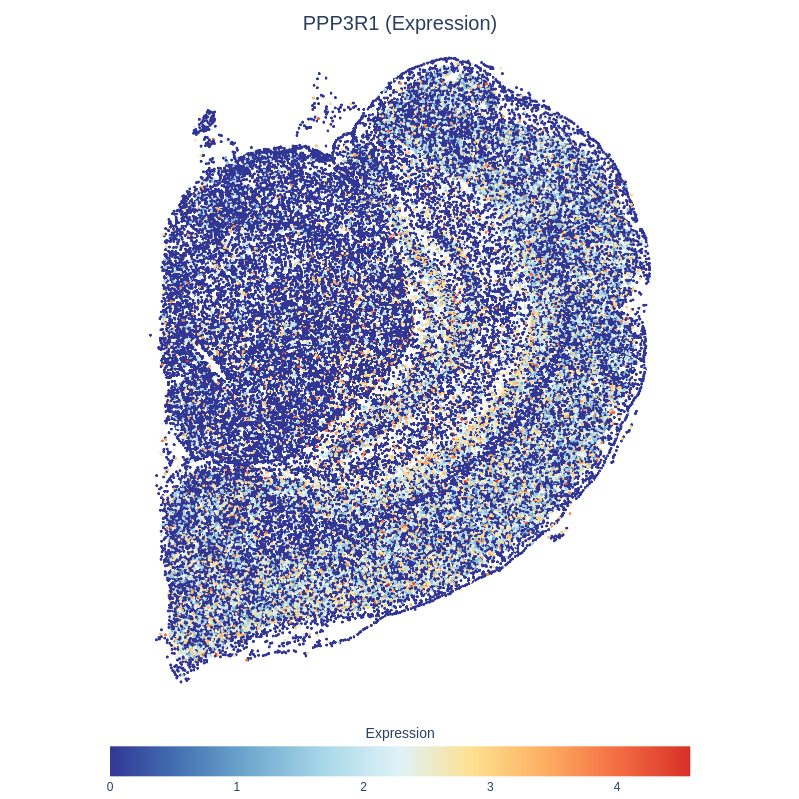

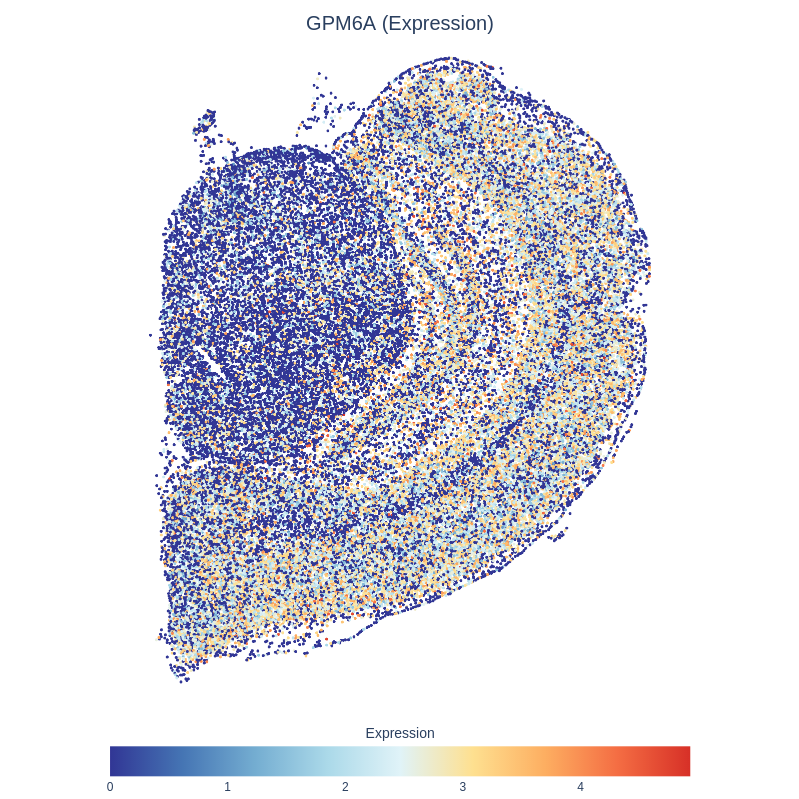

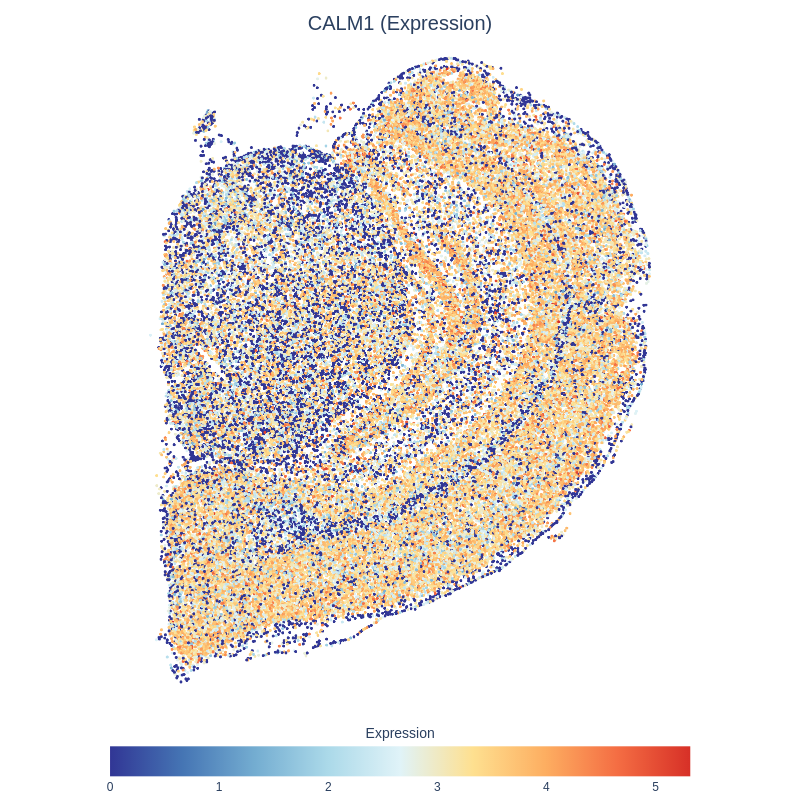

**Supplementary Figure 16. Spatial transcriptomic maps of *RASGRP1, NRGN, CALM2, CHN1, SEINC1, PPP3R1, GPM6A,* and *CALM1* genes involved in excitatory neuronal signaling within the hippocampal CA region.** The figure shows the normalised expression patterns in excitatory neurons, as identified by the gsMap method.

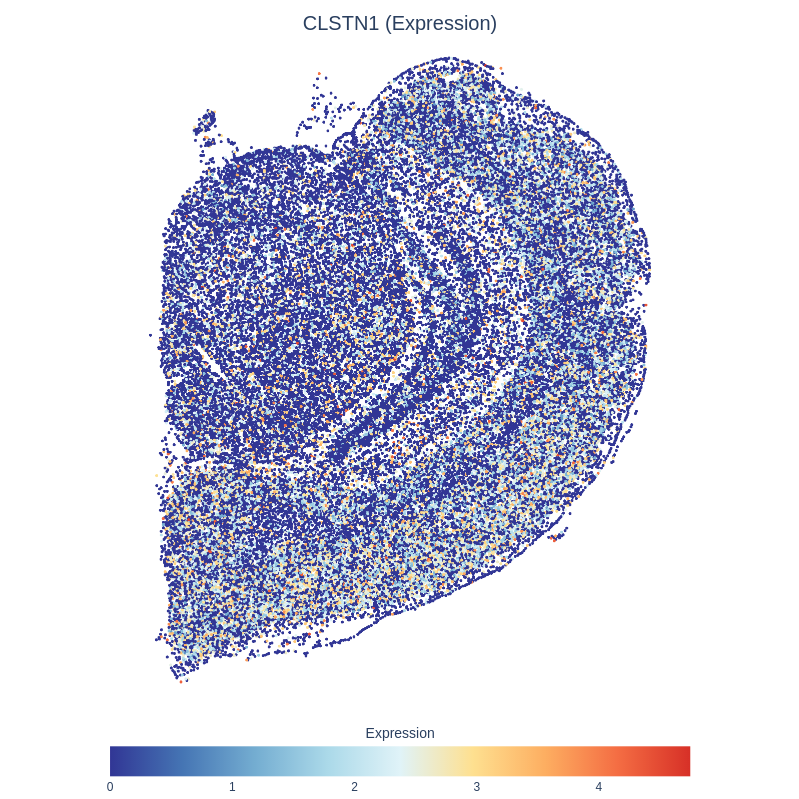

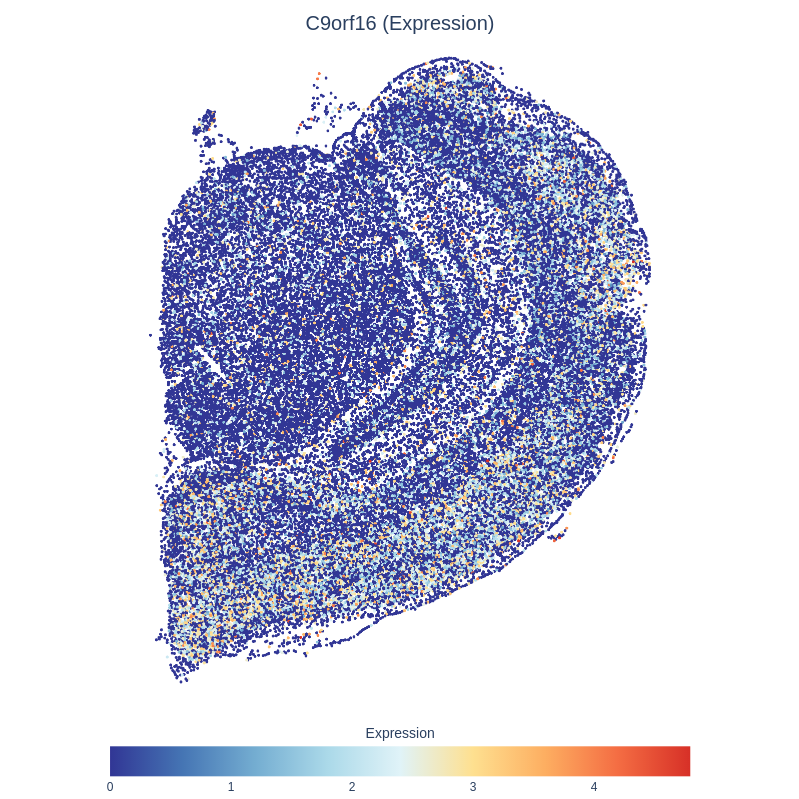

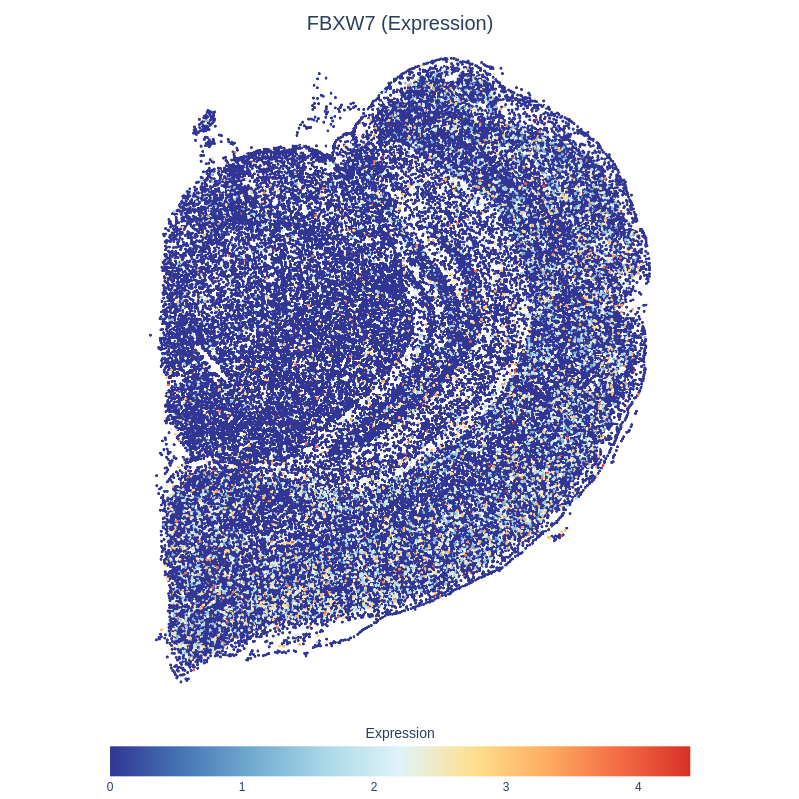

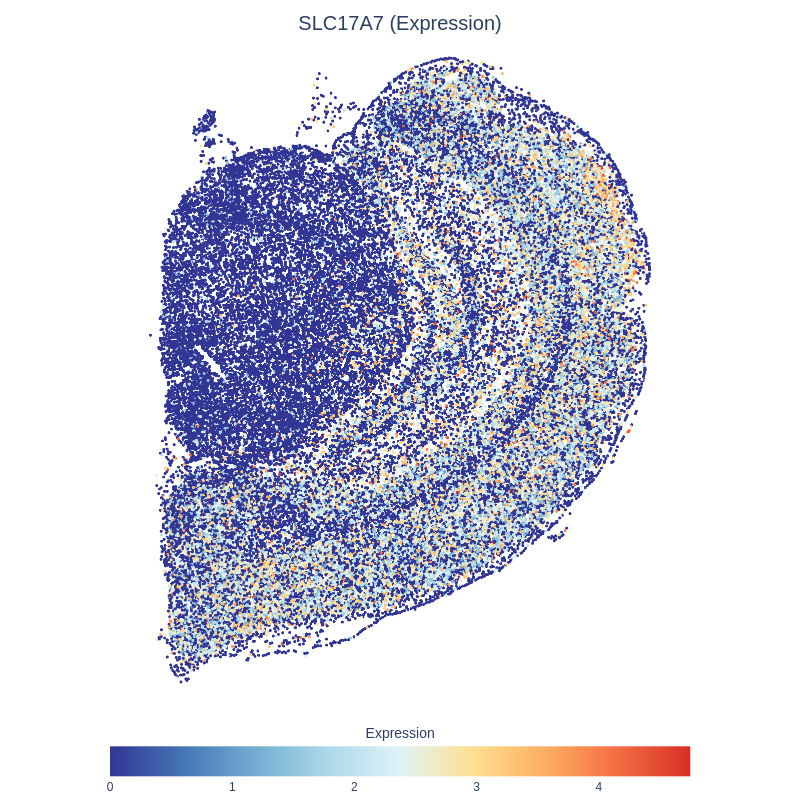

**Supplementary Figure 17. Spatial transcriptomic maps of *CLSTN1, C9orf16, FBXW7, CYFIP2* and *SLC17A7* genes involved in excitatory neuronal signaling within the LA region.** The figure shows the normalised expression patterns in excitatory neurons, as identified by the gsMap method.

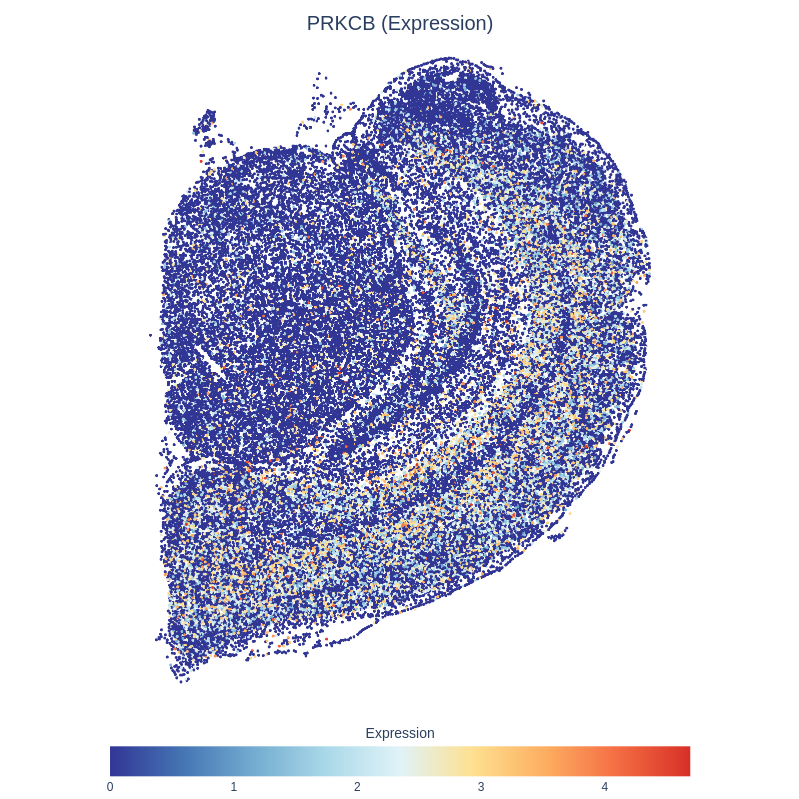

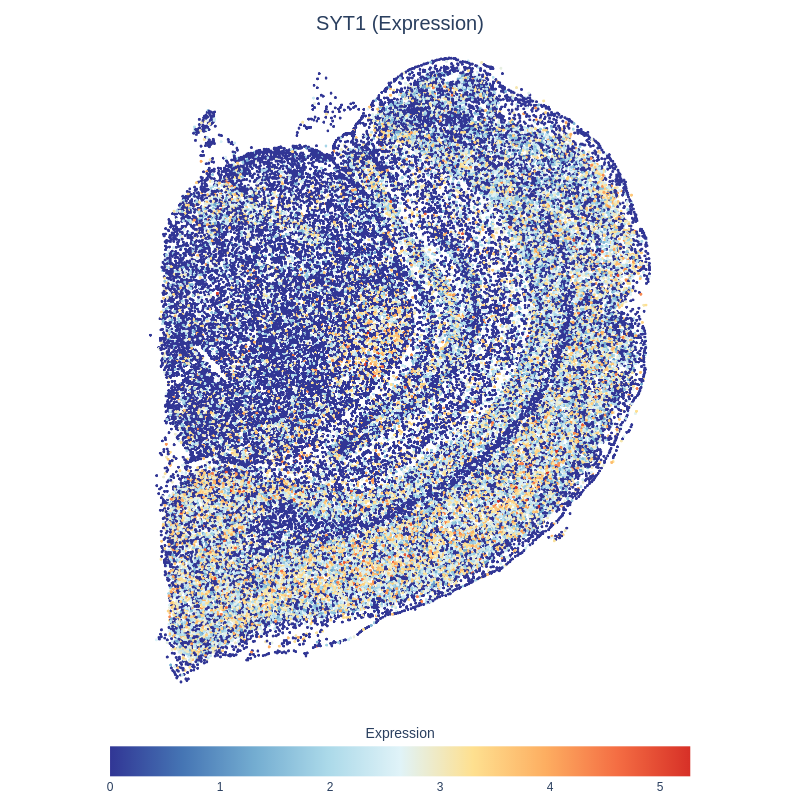

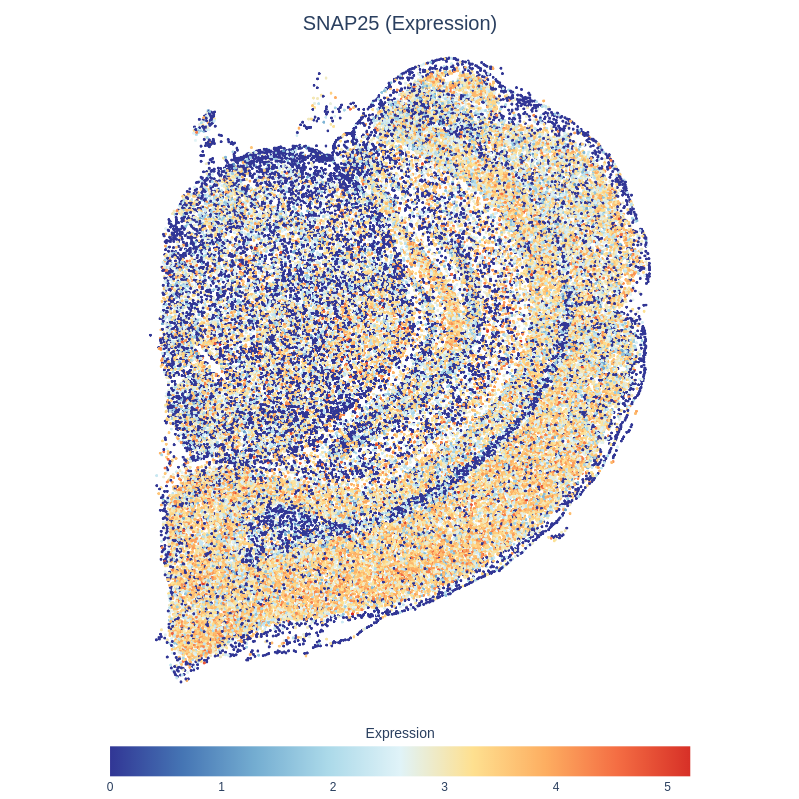

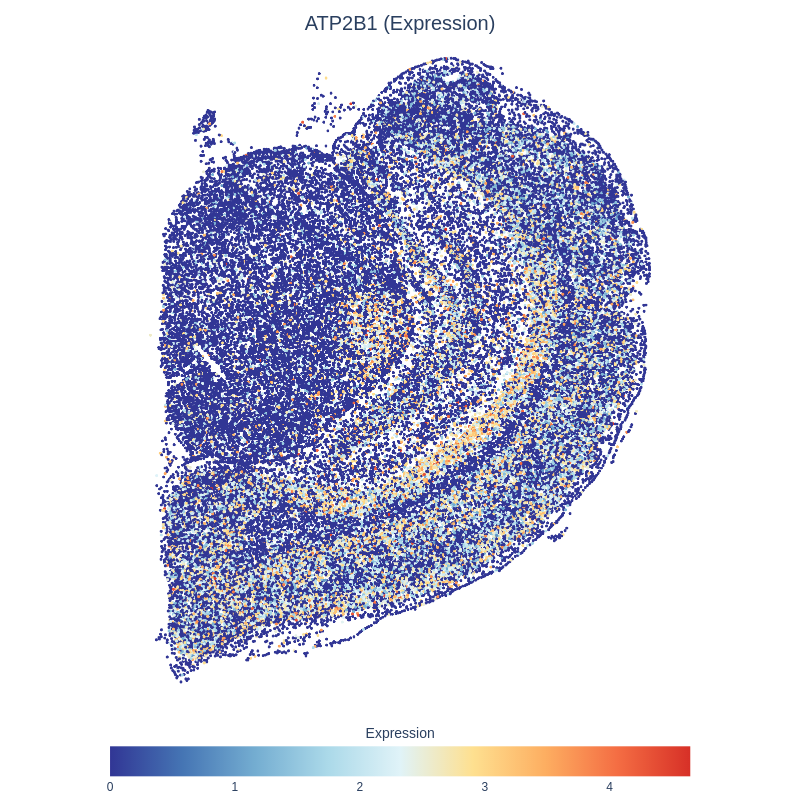

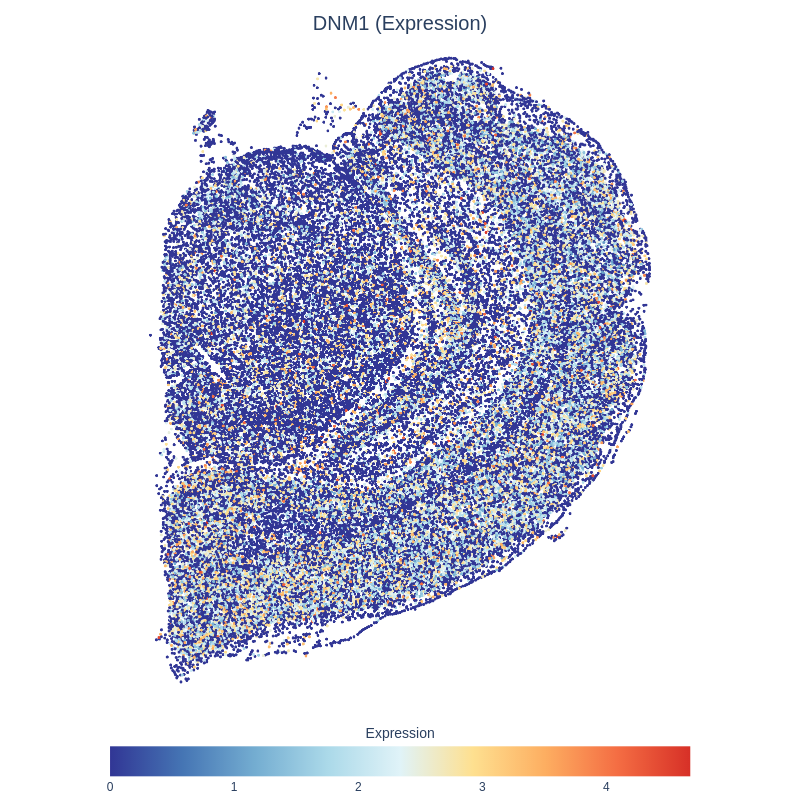

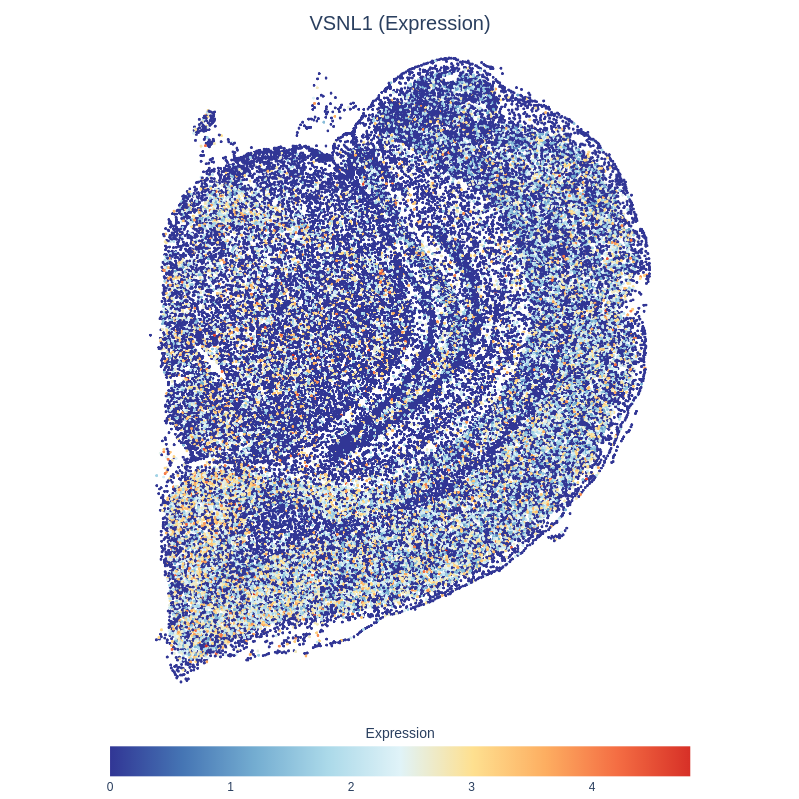

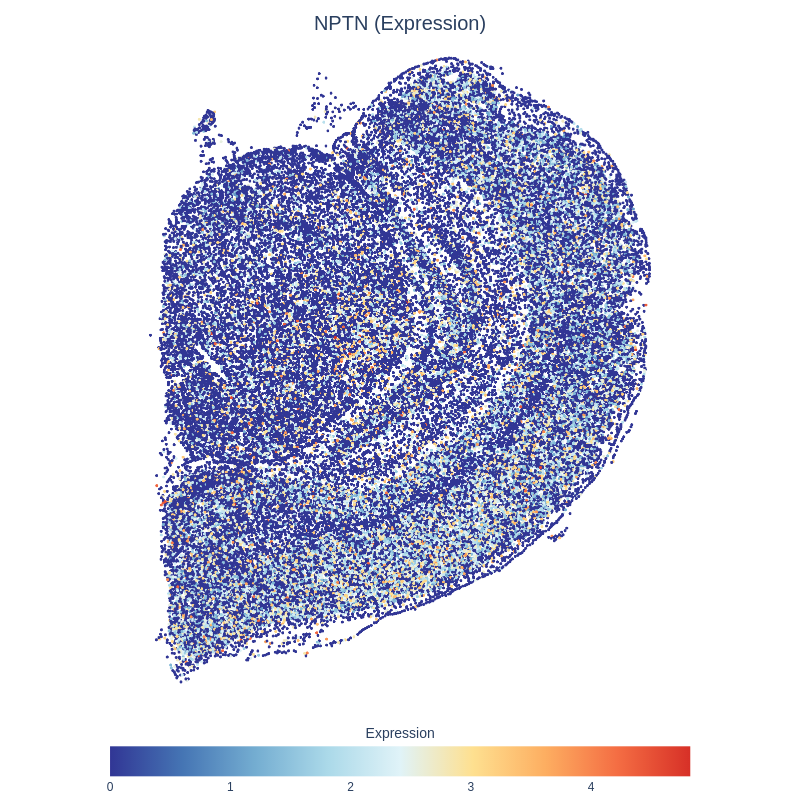

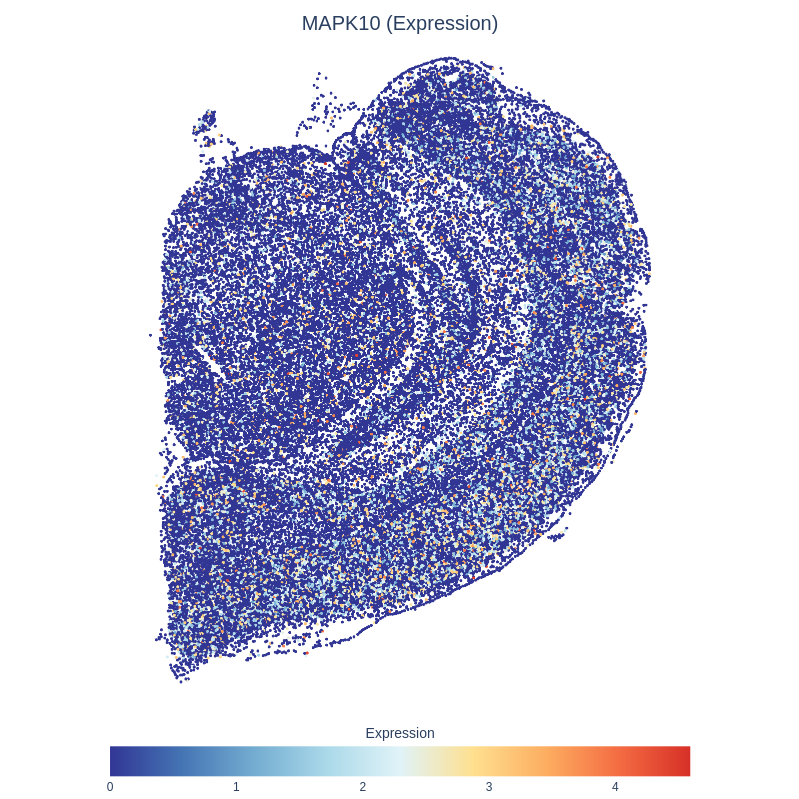

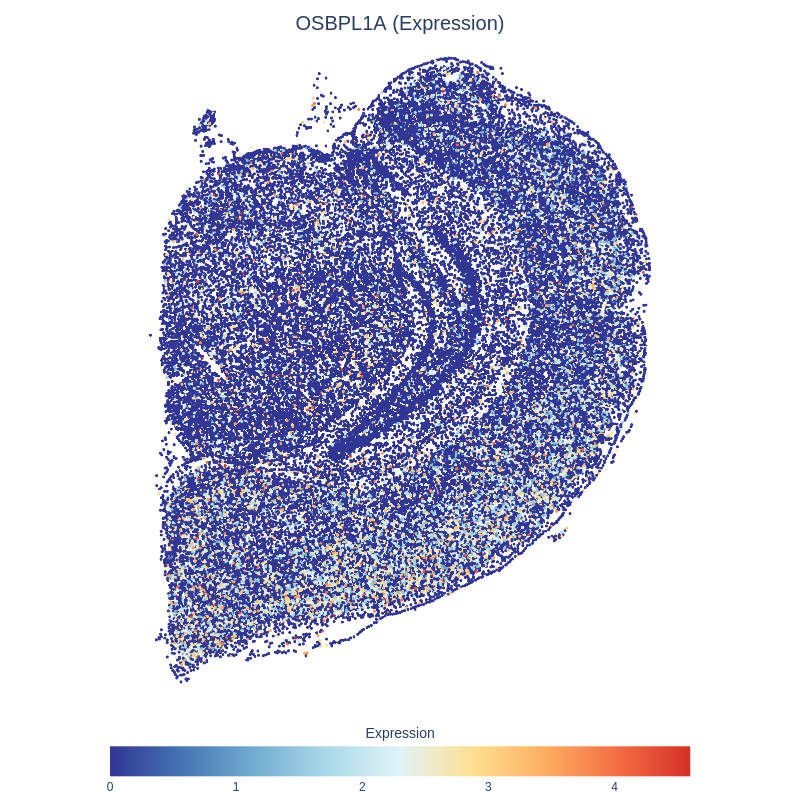

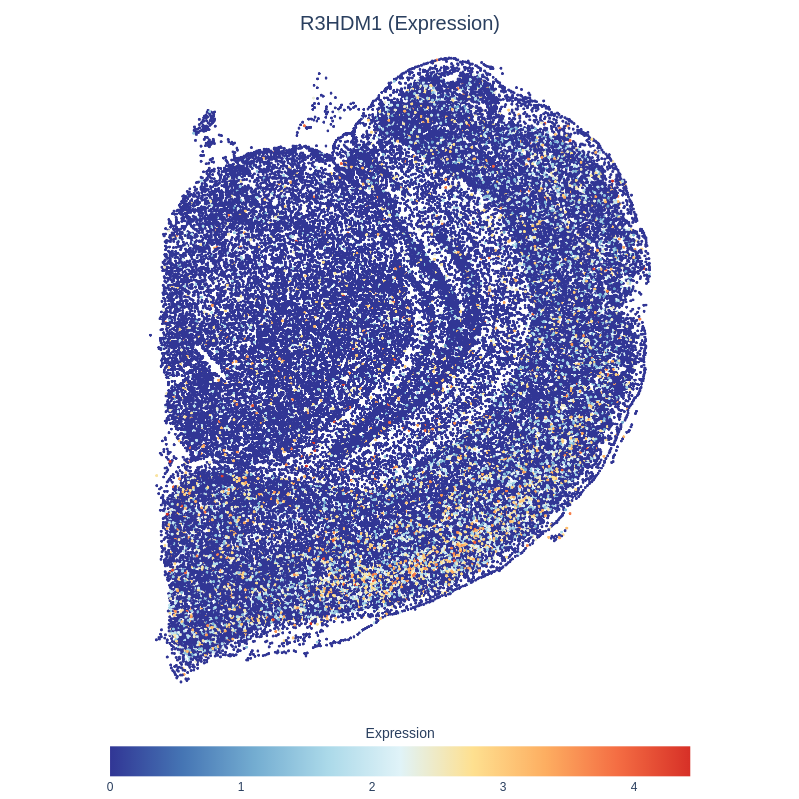

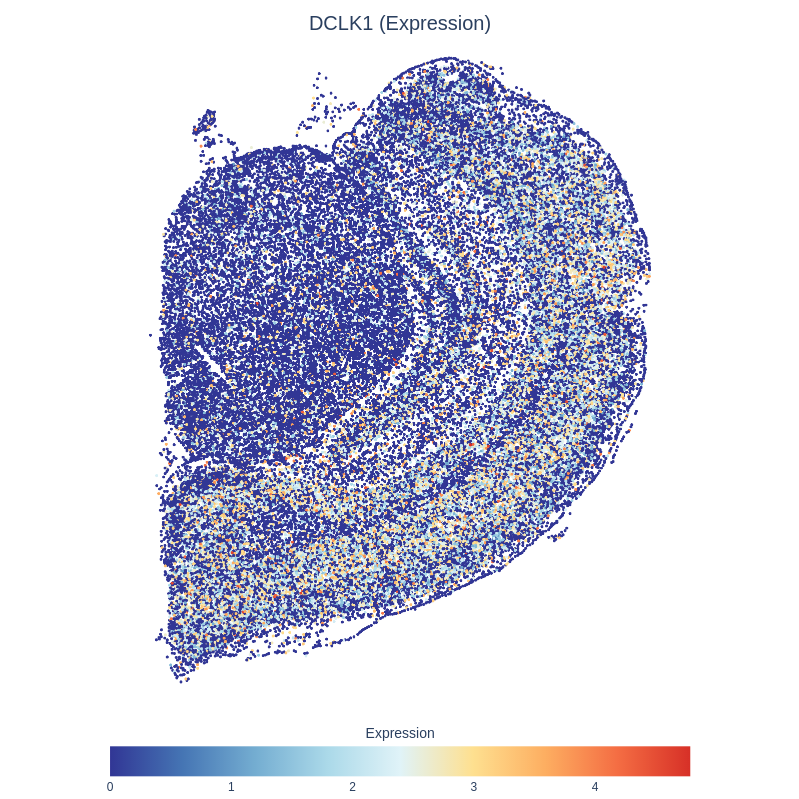

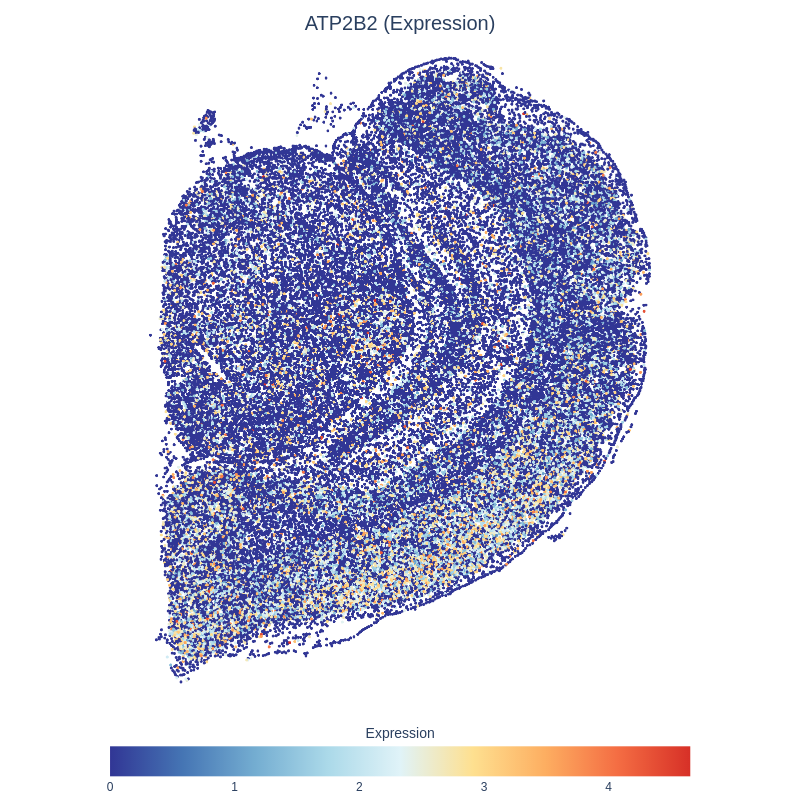

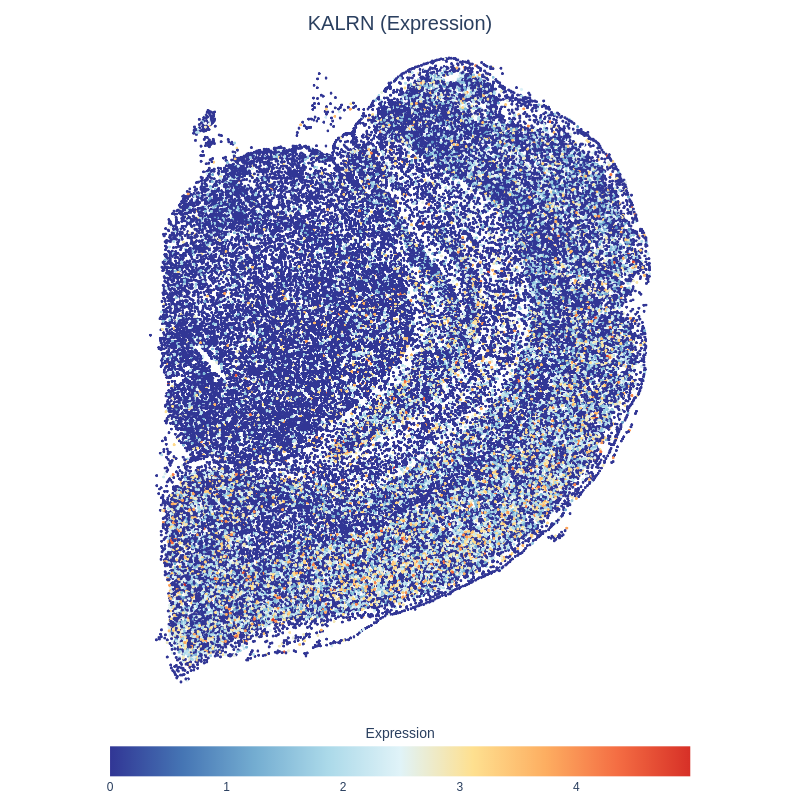

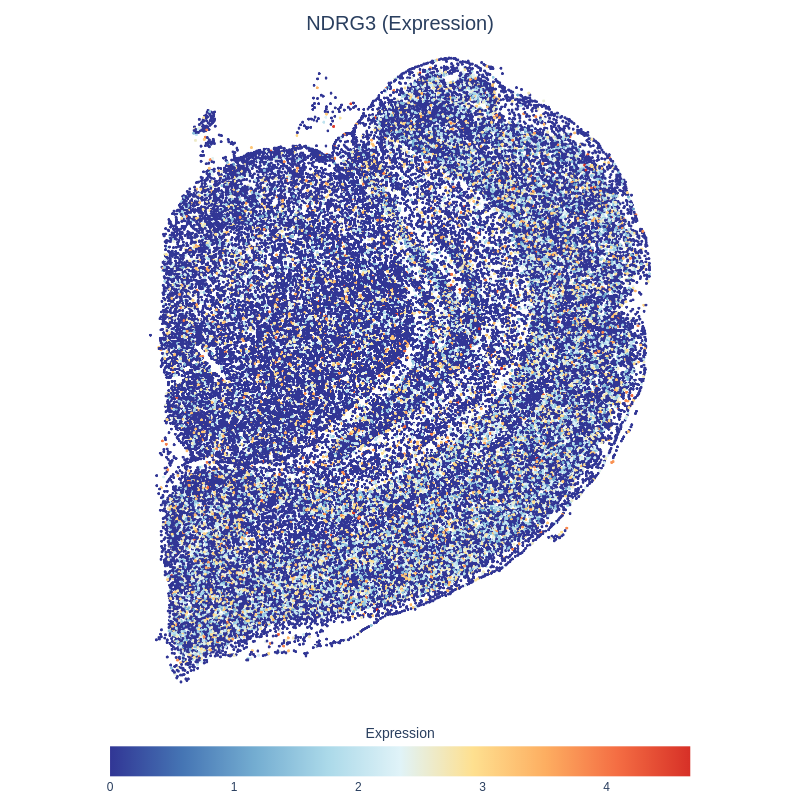

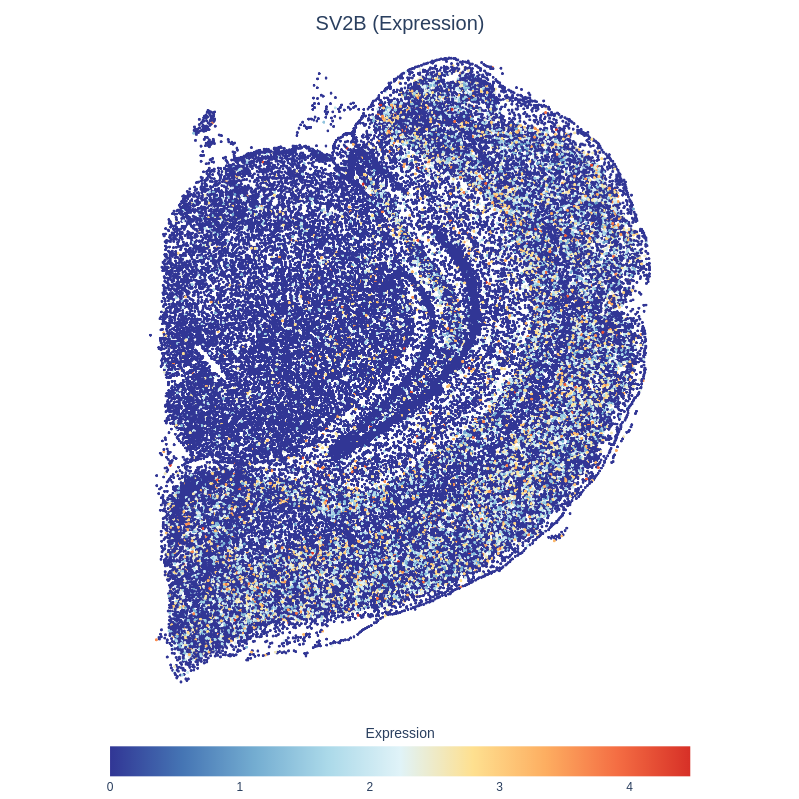

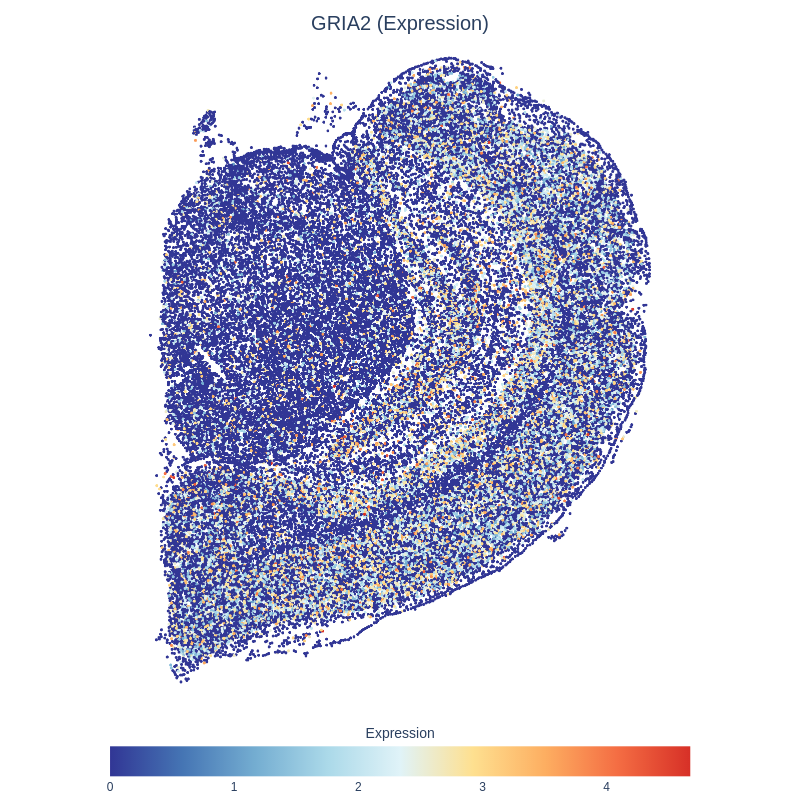

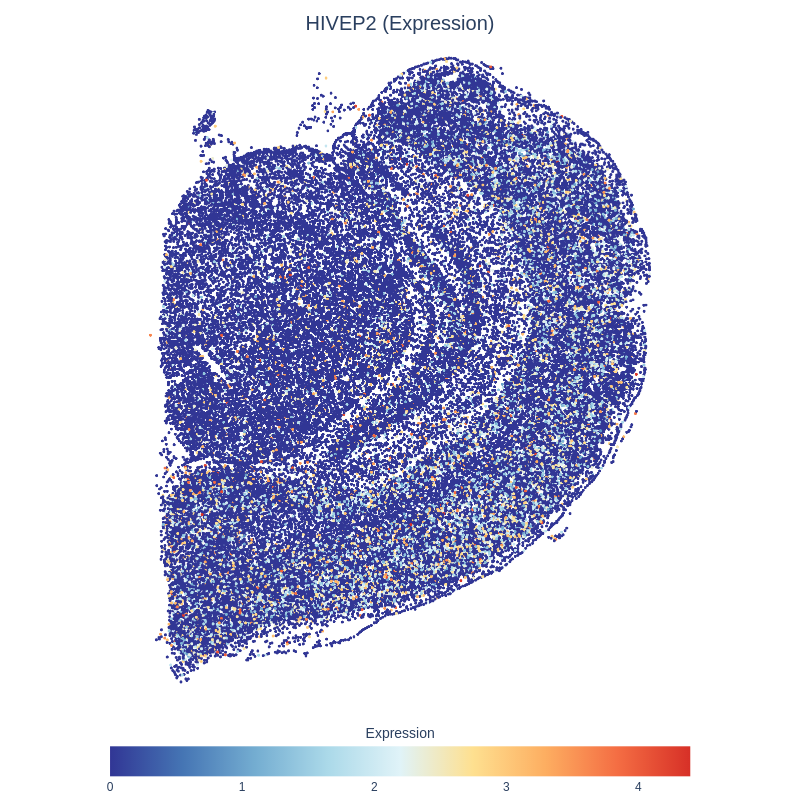

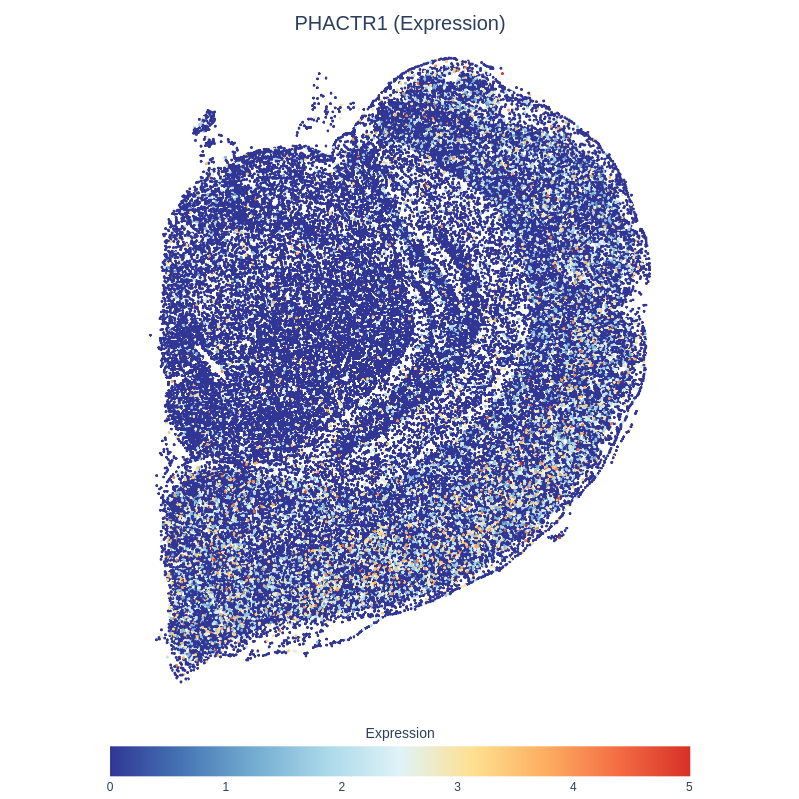

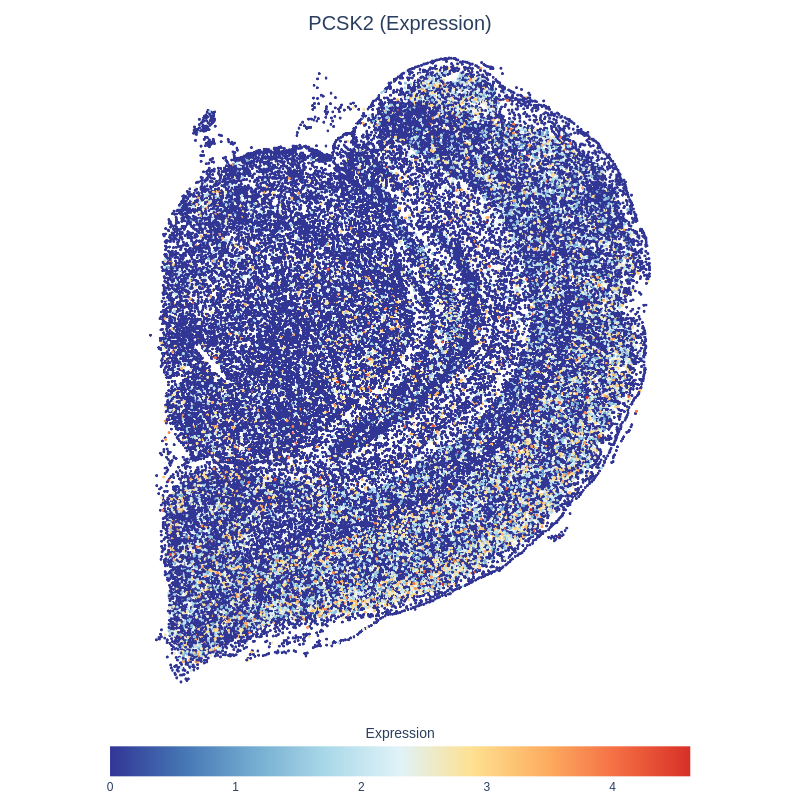

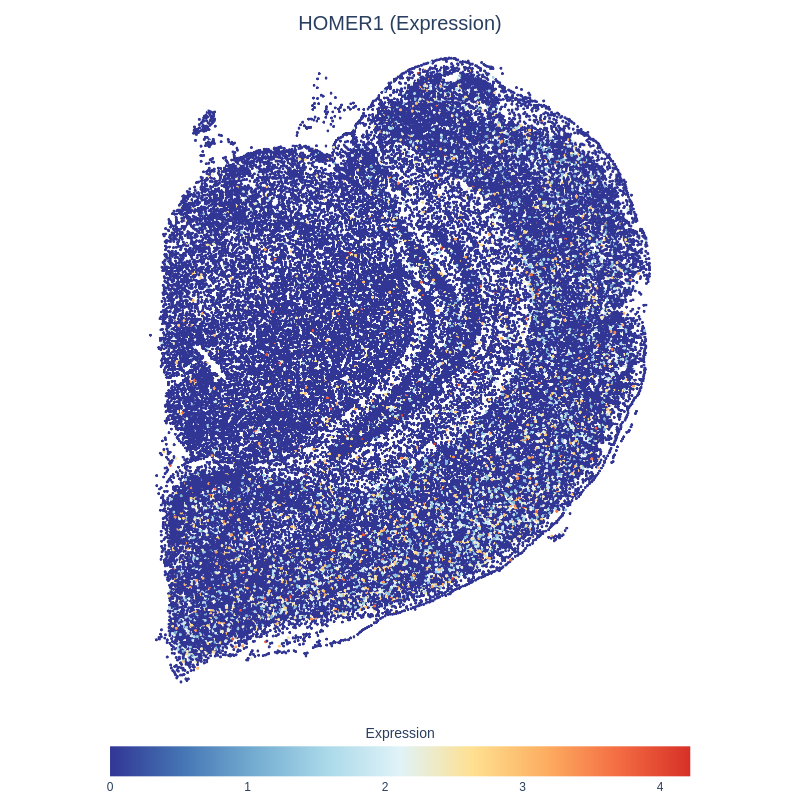

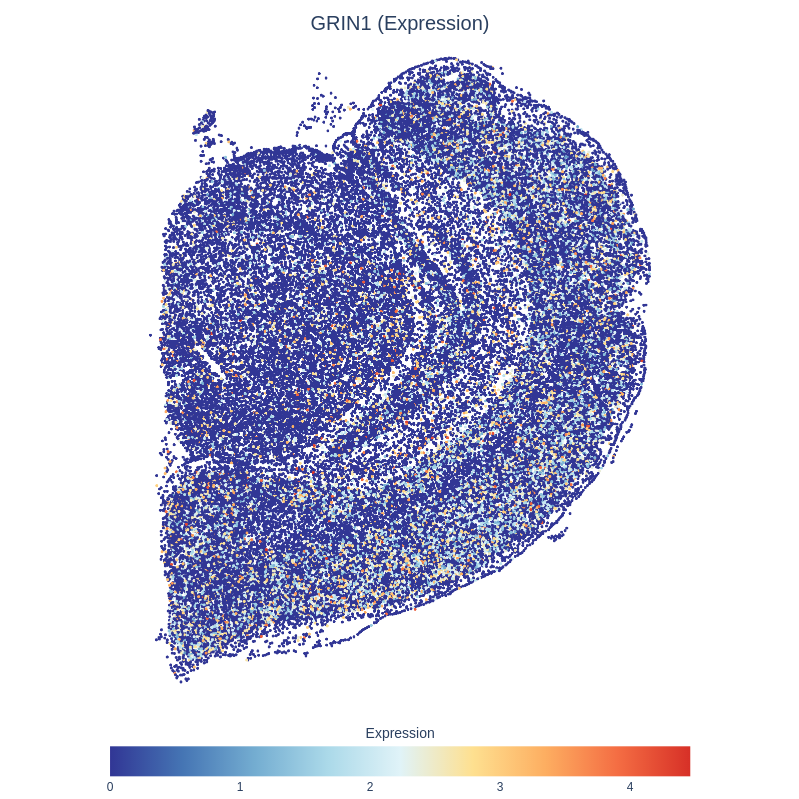

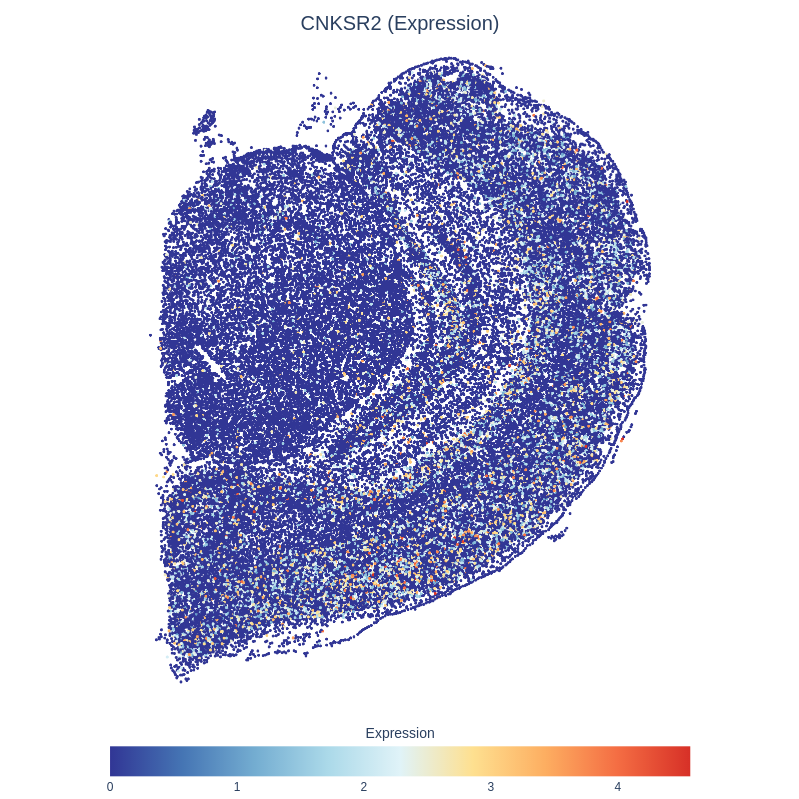

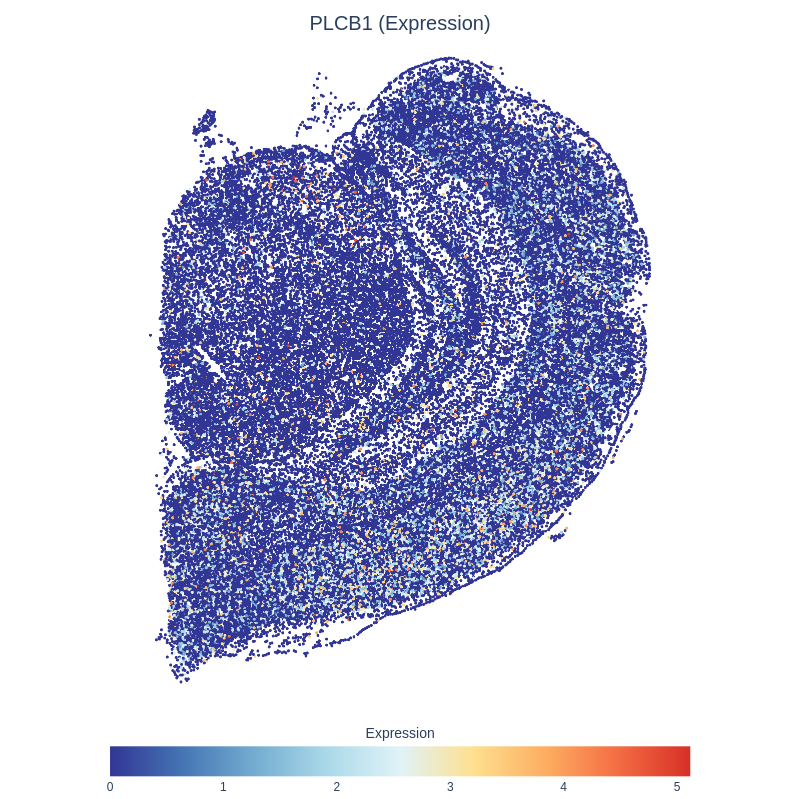

**Supplementary Figure 18. Spatial transcriptomic maps of *PRKCB, SYT1, SNAP25, SLC24A2, ATP2B1, DNM1, VSNL1, NPTN, MAPK10, OSBPL1A, R3HDM1, DCLK1, ATP2B2, KALRN, NDRG3, SV2B, GRIA2, HIVEP2, PHACTR1, PCSK2, HOMER1, GRIN1, CNKSR2, PLCB1,* and *BCL11A* genes involved in excitatory neuronal signaling within the L5/6 region.** The figure shows the normalised expression patterns in excitatory neurons, as identified by the gsMap method.

**Supplementary Figure 19. Spatial transcriptomic maps of *ATP2A2* and *VAMP2* genes involved in excitatory neuronal signaling within the L6 region.** The figure shows the normalised expression patterns in excitatory neurons, as identified by the gsMap method.

**Supplementary Figure 20. Spatial transcriptomic maps of *PPP3CA* genes involved in excitatory neuronal signaling within the GN DG region.** The figure shows the normalised expression patterns in excitatory neurons, as identified by the gsMap method.

**Supplementary Figure 21. Spatial transcriptomic maps of *SCN1B, RAB6A, NME7, NDRG4* and *ATP1B1* genes involved in excitatory neuronal signaling within the IN Pvalb+ region.** The figure shows the normalised expression patterns in excitatory neurons, as identified by the gsMap method.

**Supplementary Figure 22. Spatial transcriptomic maps of *MDH1, SNRPN* and *SCG5* genes involved in interneurons Pvalb+Gad1+.** The figure shows the normalised expression patterns in excitatory neurons, as identified by the gsMap method.

**Supplementary Figure 23. Spatial transcriptomic maps of *OPCML* gene involved in OPC region.** The figure shows the normalised expression patterns in excitatory neurons, as identified by the gsMap method.

**Supplementary Figure 24. Gene Spatial Specificity (GSS) scores of *RASGRP1, NRGN, CALM2, CHN1, SEINC1, PPP3R1, GPM6A,* and *CALM1* in excitatory neuronal signaling within the hippocampal CA region.** GSS quantifies the degree of spatially specific expression for each gene, with higher scores indicating greater localisation within subregions. These results highlight the spatial enrichment of genes involved in calcium signaling, synaptic function, and neuronal excitability across excitatory cells in the CA subfields.

**Supplementary Figure 25. Gene Spatial Specificity (GSS) scores of *CLSTN1, C9orf16, FBXW7, CYFIP2* and *SLC17A7* in excitatory neuronal signaling within LA region.** GSS quantifies the degree of spatially specific expression for each gene, with higher scores indicating greater localisation within subregions.

**Supplementary Figure 26. Gene Spatial Specificity (GSS) scores of *PRKCB, SYT1, SNAP25, SLC24A2, ATP2B1, DNM1, VSNL1, NPTN, MAPK10, OSBPL1A, R3HDM1, DCLK1, ATP2B2, KALRN, NDRG3, SV2B, GRIA2, HIVEP2, PHACTR1, PCSK2, HOMER1, GRIN1, CNKSR2, PLCB1,* and *BCL11A* in excitatory neuronal signaling within L5/6 region.** GSS quantifies the degree of spatially specific expression for each gene, with higher scores indicating greater localisation within subregions.

**Supplementary Figure 27. Gene Spatial Specificity (GSS) scores of *ATP2A2* and *VAMP2* in excitatory neuronal signaling within L6 region.** GSS quantifies the degree of spatially specific expression for each gene, with higher scores indicating greater localisation within subregions.

**Supplementary Figure 28. Gene Spatial Specificity (GSS) scores of *PPP3CA* in excitatory neuronal signaling within GN DG region.** GSS quantifies the degree of spatially specific expression for each gene, with higher scores indicating greater localisation within subregions.

**Supplementary Figure 29. Gene Spatial Specificity (GSS) scores of *PSCN1B, RAB6A, NME7, NDRG4 and ATP1B1* in excitatory neuronal signaling within IN Pvalb+ region.** GSS quantifies the degree of spatially specific expression for each gene, with higher scores indicating greater localisation within subregions.

**Supplementary Figure 30. Gene Spatial Specificity (GSS) scores of *MDH1, SNRPN* and *SCG5*** **in interneurons Pvalb+Gad1+.** GSS quantifies the degree of spatially specific expression for each gene, with higher scores indicating greater localisation within subregions.

**Supplementary Figure 31. Gene Spatial Specificity (GSS) scores of the *OPCML* gene** **in OPC region.** GSS quantifies the degree of spatially specific expression for each gene, with higher scores indicating greater localisation within subregions.

**Supplementary Figure 32.** **Difference in mean age at diagnosis for essential tremor across PGS deciles.** A higher PRS was associated with an earlier age at diagnosis; however, this association did not reach statistical significance (p > 0.05).
